# Baseline malaria infection status and RTS,S/AS01E malaria vaccine efficacy

**DOI:** 10.1101/2023.11.22.23298907

**Authors:** Michal Juraska, Angela M. Early, Li Li, Stephen F. Schaffner, Marc Lievens, Akanksha Khorgade, Brian Simpkins, Nima S. Hejazi, David A. Benkeser, Qi Wang, Laina D. Mercer, Samuel Adjei, Tsiri Agbenyega, Scott Anderson, Daniel Ansong, Dennis K. Bii, Patrick B.Y. Buabeng, Sean English, Nicholas Fitzgerald, Jonna Grimsby, Simon K. Kariuki, Kephas Otieno, François Roman, Aaron M. Samuels, Nelli Westercamp, Christian F. Ockenhouse, Opokua Ofori-Anyinam, Cynthia K. Lee, Bronwyn L. MacInnis, Dyann F. Wirth, Peter B. Gilbert, Daniel E. Neafsey

**Affiliations:** Fred Hutchinson Cancer Center, Vaccine and Infectious Disease Division, Seattle, WA, USA; Broad Institute, Infectious Disease and Microbiome Program, Cambridge, MA, USA; GSK, Wavre, Belgium; Harvard T.H. Chan School of Public Health, Department of Biostatistics, Boston, MA, USA; Rollins School of Public Health, Department of Biostatistics and Bioinformatics, Atlanta, GA, USA; Department of Statistics, University of Washington, Seattle, WA, USA; PATH, Seattle, WA, USA; Kwame Nkrumah University of Science and Technology/Agogo Presbyterian Hospital, Agogo, Asante Akyem, Ghana; Centre for Global Health Research, Kenya Medical Research Institute, Kisumu, Kenya; Malaria Branch, Division of Parasitic Diseases and Malaria, Center for Global Health, Centers for Disease Control and Prevention, Kisumu, Kenya; Malaria Branch, Division of Parasitic Diseases and Malaria, Center for Global Health, Centers for Disease Control and Prevention, Atlanta, GA, USA; Harvard T.H. Chan School of Public Health, Department of Immunology and Infectious Diseases, Boston, MA, USA; Department of Biostatistics, University of Washington, Hans Rosling Center for Population Health, Seattle, WA, USA

## Abstract

**Background:** The only licensed malaria vaccine, RTS,S/AS01_E_, confers moderate protection against symptomatic disease. Because many malaria infections are asymptomatic, we conducted a large-scale longitudinal parasite genotyping study of samples from a clinical trial exploring how vaccine dosing regimen affects vaccine efficacy (VE).

**Methods:** 1,500 children aged 5–17 months were randomized to receive four different RTS,S/AS01_E_ regimens or a rabies control vaccine in a phase 2b clinical trial in Ghana and Kenya. We evaluated the time to the first new genotypically detected infection and the total number of new infections during two follow-up periods in over 36K participant specimens. We performed a post hoc analysis of VE based on malaria infection status at first vaccination and force of infection.

**Results:** We observed significant and comparable VE (25–43%, 95% CI union 9–53%) against first new infection for all four RTS,S/AS01_E_ regimens across both follow-up periods (12 and 20 months). Each RTS,S/AS01_E_ regimen significantly reduced the number of new infections in the 20-month follow-up period (control mean 4.1 vs. RTS,S/AS01_E_ mean 2.6–3.0). VE against first new infection was significantly higher in participants who were malaria-infected (68%; 95% CI, 50 to 80%) versus uninfected (37%; 95% CI, 23 to 48%) at the first vaccination (P=0.0053) and in participants experiencing greater force of infection between dose 1 and 3 (P=0.059).

**Conclusions:** All tested dosing regimens blocked some infections to a similar degree. Improved VE in participants infected during vaccination could suggest new strategies for highly efficacious malaria vaccine development and implementation. (ClinicalTrials.gov number, NCT03276962)

## Introduction

Malaria infection by the *Plasmodium falciparum* parasite causes over 230 million cases and 600,000 deaths per year, and progress in reducing morbidity and mortality through vector control and drug treatment has stalled.^1^ RTS,S/AS01_E_ (hereafter referred to as RTS,S; GSK, Wavre, Belgium) is the first vaccine recommended for *P. falciparum* malaria by the World Health Organization and it provides moderate protective efficacy against clinical malaria. Improving protective efficacy is a major goal of ongoing work, including testing alternative dosing schedules and gaining a greater understanding of the mechanism of protection.

Most malaria vaccine trials evaluate vaccine efficacy (VE) using clinical disease as an endpoint, but enhanced understanding of the mechanism and magnitude of protection may be gained from molecular detection of new infections, given that a large proportion of malaria infections are asymptomatic.^2^ Here we report on the MAL-095 study, a genotyping investigation of infection endpoints employing samples from the MAL-094 phase 2b randomized controlled trial of RTS,S. The MAL-094 trial enrolled 5-17 month-old children in Ghana and Kenya and used clinical disease endpoints to investigate the effect of dosing regimen on VE, ultimately finding no significant differences in VE against clinical disease between a delayed third dose regimen (R017), a fractional third dose regimen (Fx012), and the standard full dose regimen (R012).^3^

To explore protection in that study using a molecular infection endpoint, we genotyped > 36,000 blood samples taken both at symptomatic clinic visits and at monthly cross-sectional timepoints. We used a genotyping assay that detects infections at a sub-microscopic level and distinguishes newly incident superinfections from persistent asymptomatic infections, yielding the capacity to measure both the time to first new infection and the cumulative number of new parasite infections post-vaccination. We additionally assessed VE according to genotype of the infecting parasites given the previous observation of allele-specific VE in the phase 3 RTS,S trial.^4^ Because our genotyping assay detects newly incident superinfections in individuals with pre-existing infections, we performed a post hoc analysis of VE based on infection status at first vaccination to test the common hypothesis cited in other recent studies that an erythrocytic malaria infection during vaccination impairs development of a protective immune response.^5,6^

## Methods

### Study Design and Sequence-Data Generation

As described fully in the primary analysis of the parent study evaluating protection against clinical disease,^3^ participants 5–17 months in age were enrolled at study sites in Agogo, Ghana, and Siaya, Kenya (750 per site), and randomly assigned into one of five vaccination groups (1:1:1:1:1). The control rabies vaccination group was vaccinated at months 0, 1, and 2. Participants in the RTS,S groups received two full doses at months 0 and 1 and either full doses at months 2 and 20 (group R012-20), full doses at months 2, 14, 26, and 38 (group R012-14), fractional doses at months 2, 14, 26, and 38 (group Fx012-14; early fourth dose), or fractional doses at months 7, 20, and 32 (group Fx017-20; delayed third dose; **Fig. S1–S3** in the Supplementary Appendix).

Participant samples were collected as dried blood spots (DBS) on Whatman FTA sample cards at the baseline enrollment visit (M0), cross-sectionally at monthly intervals until study month 20, and during febrile clinic visits. Blood smears were collected for microscopy-based detection of infection. For asymptomatic individuals, blood smears were evaluated at a later date and did not trigger treatment to clear infection. Participants meeting the primary or secondary case definitions of malaria were treated according to the national guidelines of each country, with the primary case definition being >5000 asexual parasites per μL and fever (axillary temperature ≥ 37.5°C), and the secondary case definition being any parasitemia > 0 and fever or history of fever within 24 h of presentation.

We analyzed all DBS samples by extracting DNA and performing Illumina-based amplicon sequencing of the *cs* C-terminus coding region (*cs*) and a comparably polymorphic coding region for the antigen *sera2*. We defined distinct haplotypes as the combined genotype of all nucleotide variants in a given amplicon sequence. Complexity of Infection (COI) was defined as the maximum number of distinct haplotypes detected in a sample at either amplicon. We declared a new parasite infection on a specific sampling date if at least one haplotype was observed for either amplicon that had not been previously detected in the preceding three samples from that individual. Haplotype diversity was high at both study sites for both the *cs* and *sera2* amplicons (**Fig. S4** in the Supplementary Appendix). A full description of molecular methods, data filtration, and sequence analysis is in the Supplementary Appendix. All MiSeq data were submitted to the NCBI Sequence Read Archive (BioProject PRJNA983279).

### Study Oversight

The trial (MAL-095; NCT03281291) and its parent study (MAL-094; NCT03276962) were sponsored by GlaxoSmithKline Biologicals SA, the vaccine developer and manufacturer, and funded by both GlaxoSmithKline Biologicals SA and the PATH Malaria Vaccine Initiative, which received a grant from the Bill and Melinda Gates Foundation. The trial protocol was approved by all relevant ethical review boards and signed or witnessed thumbprint informed consent was obtained from the children’s parents or guardians prior to participation.

### Statistical Analysis

All analyses planned prior to the execution of this study are described in the Statistical Analysis Plan (SAP) included in the Supplementary Appendix. In short, we first assessed VE of each RTS,S regimen vs. rabies control and relative VE comparing RTS,S regimens head-to-head to prevent the first new genotypic infection and to reduce the number of new genotypic infections. These analyses were performed in parallel for the Exposed Set (ES) of participants who received the first vaccine dose (**Fig. S2** in the Supplementary Appendix) and the Per-Protocol Set (PP) of participants who received the first 3 doses per protocol and were in follow-up at 14 days after the third dose (**Fig. S3** in the Supplementary Appendix). We analyzed the follow-up period from the first dose to the month 20 visit in the ES and from 14 days after the third dose to a visit scheduled 12 months after the third dose in the PP.

To study vaccine effects on time to the first new infection, we defined VE as one minus the hazard ratio (RTS,S vs. control) of the first new infection estimated using the Cox proportional hazards model with 95% Wald confidence intervals (CIs) and two-sided Wald tests of zero VE. For RTS,S head-to-head comparisons, relative VE was defined analogously replacing the control with an active comparator regimen. Further, we estimated instantaneous VE over time with 95% pointwise and simultaneous CIs using nonparametric kernel-smoothing^7^ and tested for variation in VE across time^8^. Cumulative incidence of the first new infection was estimated using the transformed Nelson-Aalen estimator for the cumulative hazard function.

We measured vaccine effects on the number of new infections by the additive difference (RTS,S vs. comparator) in the mean number of new infections. The infection count was defined as unobserved if the number of missed visits or samples exceeded a specified threshold (Supplementary Appendix). We assessed the mean difference by targeted maximum likelihood estimation (TMLE)^9^ accounting for unobserved infection counts. Additionally, we employed TMLE to estimate reverse cumulative distribution functions of the number of new infections in each study group.

Besides overall VE, we assessed in the PP whether and how VE against the first new infection depended on genotypic characteristics of infecting parasites using augmented inverse probability weighting^10,11^ and their complete-case analogs^12^ (details in Supplementary Appendix).

Finally, we assessed whether baseline parasite positivity and/or infection risk modified the effect of RTS,S on the time to the first new genotypic infection and the time to the first new clinical malaria episode. Covariate-adjusted Cox proportional hazards models with separate baseline hazards for each study site, employing 95% Wald CIs and Wald interaction tests, and Nelson-Aalen-based cumulative incidence curves were used. We performed a sensitivity ‘matching’ Cox analysis with stratified sampling, wherein baseline negative participants were randomly sampled from the same randomization group and study site by matching baseline positive participants on the date of the third vaccination to address potential confounding by low- vs. high-transmission season. Additionally, a sensitivity analysis with E-values quantified the robustness of evidence for baseline parasite positivity and M2-FOI causally modifying vaccine efficacy (details in Supplementary Appendix).

All aforementioned analyses were performed on pooled data from both study sites as well as separately within each site. Tests for VE departing from 0 were adjusted for multiplicity separately within each analysis cohort, study site-pooled vs. -specific analysis, and each of the three sets of treatment comparison types defined as follows: comparisons vs. the control regimen other than the primary comparisons of each of Fx012-14 and Fx017-20 vs. control, comparisons vs. the “standard” RTS,S regimen R012-20, and head-to-head comparisons of novel RTS,S regimens (details in the Supplementary Appendix). For each multiplicity set, P-value adjustments were implemented to control the familywise error rate (FWER) (Holm-Bonferroni^13^) and the false discovery rate (FDR) (Q-values; Benjamini-Hochberg^14^). We defined FWER statistical significance as an FWER-adjusted P-value ≤ 0.05 and FDR statistical significance as a Q-value ≤ 0.2 together with an unadjusted P-value ≤ 0.05. All P-values are two-sided except P-values testing differential VE by 3D7 Hamming distances and by COI, which are double one-sided.

## Results

### Participants and Genotypic Endpoints

The ES comprised 1,500 children (750 per study site), with 36,080 specimens collected between the first dose and the month 20 visit and 35,456 (98.3%) with genotyping completed (**Fig. S2** in the Supplementary Appendix). Of those, 5,078 (14.3%) were confirmed parasite-positive, among which 3,937 (77.5%) were associated with a new infection. The PP comprised 1,332 children (687 in Agogo and 645 in Siaya), with 34,147 specimens collected during the PP follow-up period (14 days to 12 months after the third dose) and 33,547 (98.2%) with genotyping completed (**Fig. S3** in the Supplementary Appendix). Of those, 4,746 (14.1%) were confirmed parasite-positive, among which 3,690 (77.7%) were associated with a new infection. In the ES and PP, 1,030 (68.7%) and 763 (57.3%) participants, respectively, experienced the first new genotypic infection during the respective follow-up. The median time from the first (third) dose to the first new infection in the ES (PP) was 40.9 (37.0) weeks. The mean number of new genotypic infections per individual was 2.9 and 1.5 in the ES and PP, respectively.

### Vaccine Efficacy against First New Infection

VE of each RTS,S regimen vs. the control regimen was 25–31% (95% CI union, 9 to 43) in the ES and 37–43% (95% CI union, 21 to 53) in the PP, each significantly different from zero (all P<0.0033 in the ES and <0.001 in the PP) (**Fig. 1** and **Fig. S5** in the Supplementary Appendix). No significant differences in the hazard rate of the first new infection were found in head-to-head comparisons of RTS,S regimens (all P>0.32) (**Fig. S6** in the Supplementary Appendix). Instantaneous VE over time suggests that the full dose at month 2 may have provided a more sustained protection than a fractional dose at month 2, as VE of Fx012-14 waned to zero by 7 months after the third dose (**Fig. S7** in the Supplementary Appendix).

**Figure 1.**
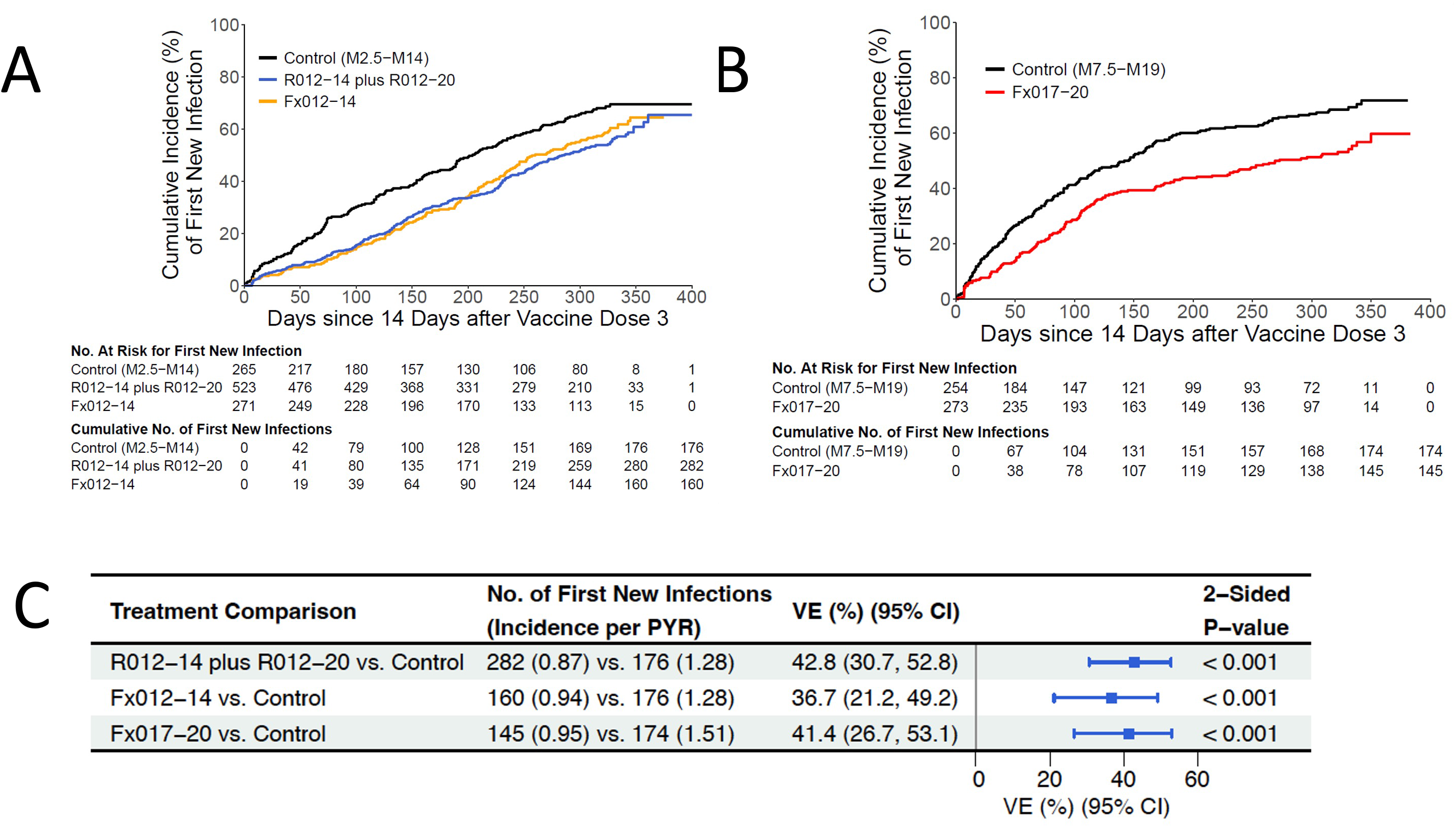
Cumulative incidence and vaccine efficacy against the first new genotypic infection for the PP cohort. First new genotypic infection (A) between 14 days after month 2 through month 14 for R012-14 plus R012-20 and Fx012-14 vs. the control regimen and (B) between 14 days after month 7 through month 19 for Fx017-20 vs. the control regimen. (C) Forest plot of vaccine efficacies against the first new infection vs. the control regimen. PYR: person-year at risk; VE: vaccine efficacy; CI: confidence interval; PP: Per Protocol cohort; No: number.

### Vaccine Efficacy to Reduce Number of New Infections

The mean number of new infections in RTS,S recipients was significantly lower than that in control recipients in both the ES and PP (all P<0.001) (**Fig. 2** and **Fig. S8** in the Supplementary Appendix). In the ES, the mean new infection count during 20 months ranged between 2.6–3.0 among RTS,S recipients and was 4.1 among control recipients, with the mean difference ranging –1.6 to –1.1 (95% CI union, –2.1 to –0.6). In the PP, the mean new infection count between 14 days and 12 months after the third dose ranged between 1.4–1.5 among RTS,S recipients and was 2.2 and 2.7 among control recipients between months 2.5–14 or 7.5–19, respectively, with the mean difference ranging –1.3 to –0.8 (95% CI union, –1.6 to –0.4).

**Figure 2.**
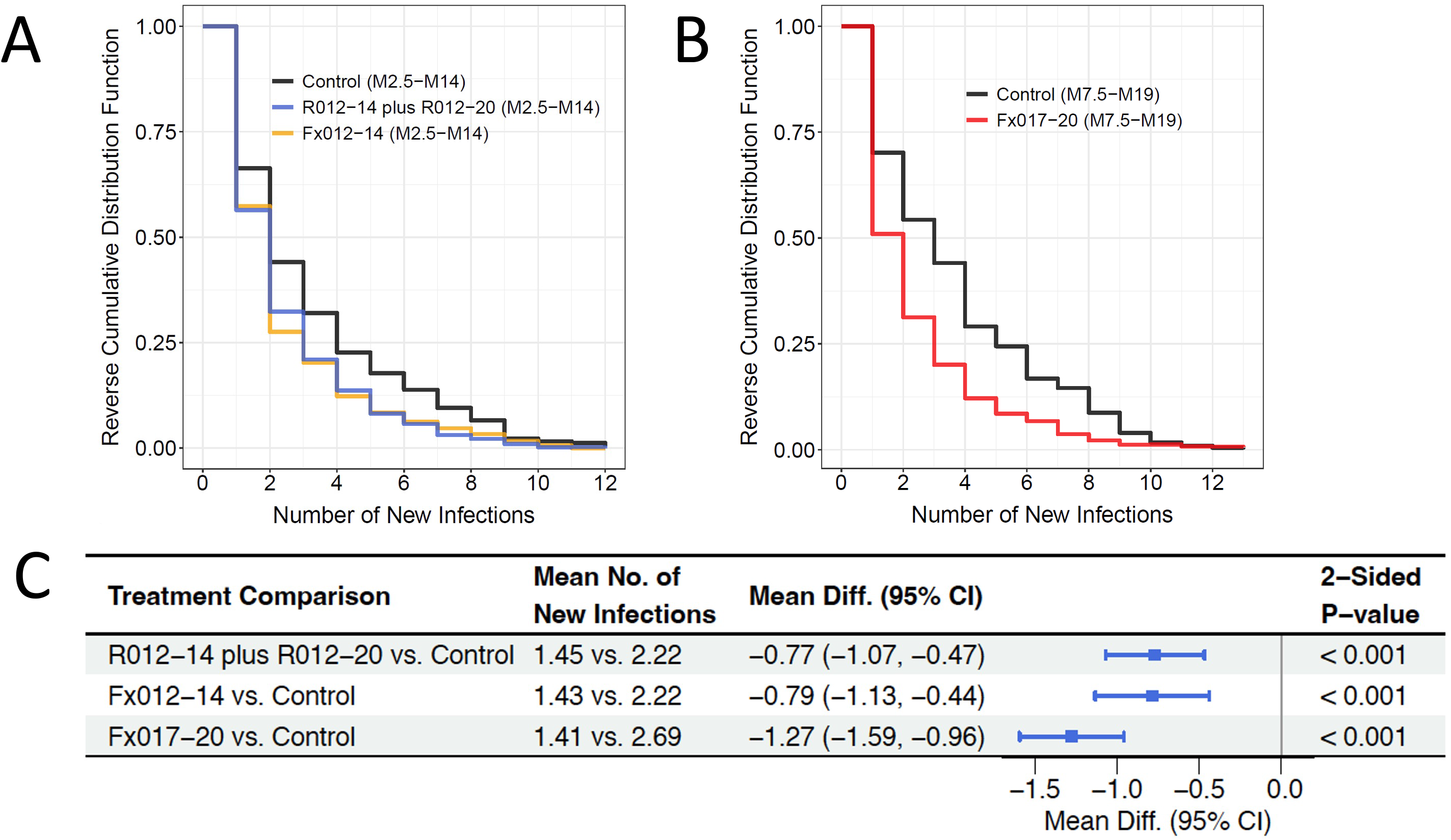
Reverse cumulative distribution function and vaccine effect on the number of new molecular infections in the Per-Protocol (PP) cohort (A) between 14 days after month 2 through month 14 for R012-14 plus R012-20 and Fx012-14 vs. the control regimen and (B) between 14 days after month 7 through month 19 for Fx017-20 vs. the control regimen. (C) Forest plot of vaccine effects on the mean number of new infections vs. the control regimen. CI: confidence interval; PP: Per Protocol cohort; No: number; Diff: Difference.

### Haplotype Variation

RTS,S regimens diminished the COI of the first new infection compared to the control regimen in the PP (**Fig. 3A** and **3C**). Moreover, RTS,S regimens exhibited a significantly greater reduction in the risk of more highly polyclonal first new infections (**Fig. 3B** and **3D**). The estimated risk reduction of pooled R012-14, R012-20, and Fx012-14 vs. control was 29% (95% CI, 13 to 42) against single-haplotype first new infections and 76% (95% CI, 58 to 86) against first new infections with 5 haplotypes (P<0.001 for increasing risk reduction with COI).

**Figure 3.**
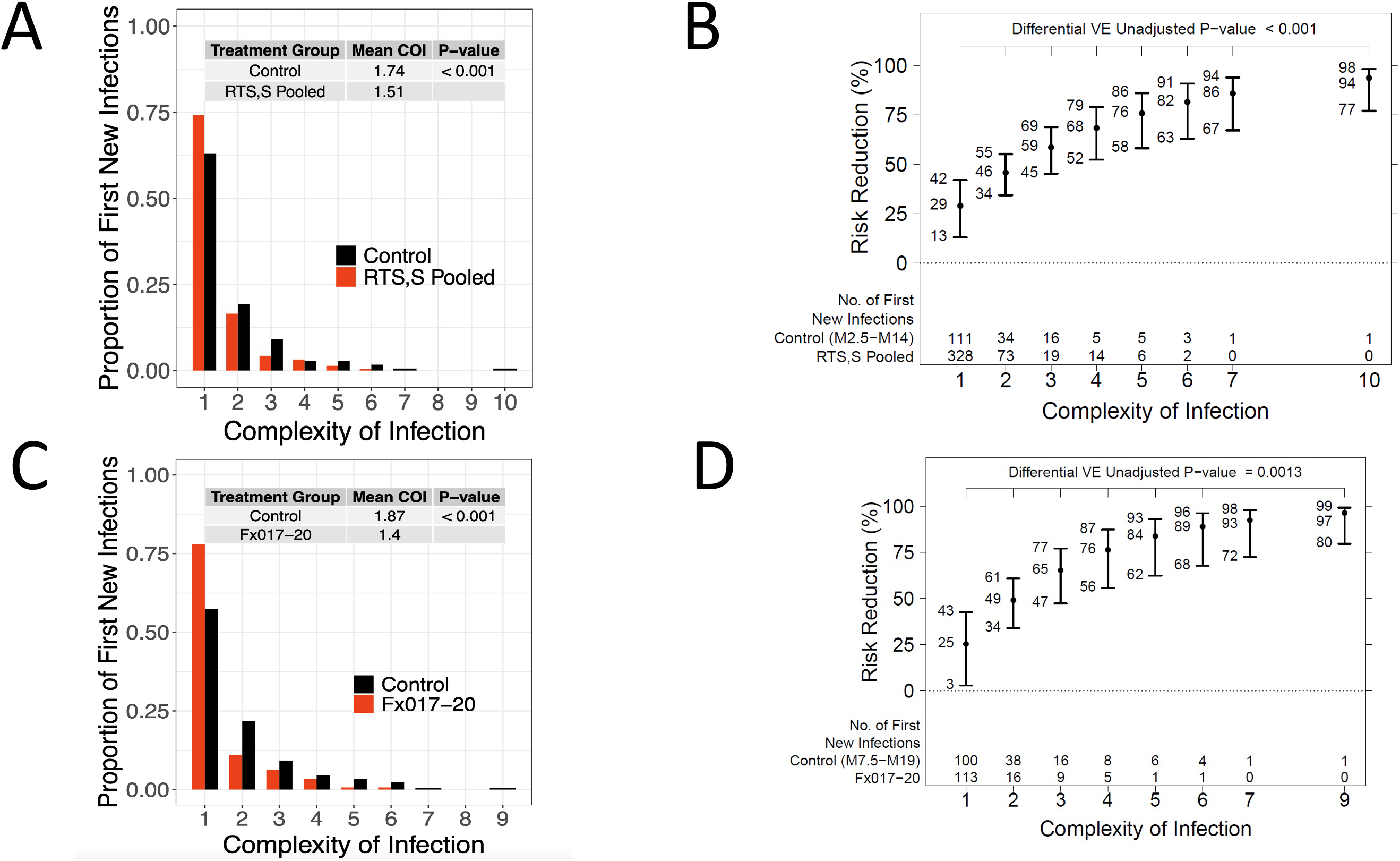
Comparisons of Complexity of infection (COI) of first new genotypic infections between the pooled R012-14, R012-20, and Fx012-14 RTS,S regimens vs. the control regimen (for new infections between 14 days after month 2 through month 13) and Fx017-20 vs. the control regimen (for new infections between 14 days after month 7 through month 19) for the Per-Protocol (PP) cohort: (A), (C) frequencies and (B), (D) risk reduction (1 – hazard ratio) against the first new genotypic infection with a given level of COI.

The genotypic sieve analysis was underpowered given the low 3D7 prevalence and small endpoint counts. No evidence was found for differential VE against the first new infection with a perfect amino acid residue match vs. mismatch to the 3D7 vaccine strain in the *cs* C-terminus full amplicon or haplotypic regions (**Fig. S9** in the Supplementary Appendix). There was a non-significant trend of a VE decline with an increasing degree of residue mismatch to 3D7 in the *cs* C-terminus (**Figure S10** in the Supplementary Appendix). Scanning individual polymorphic amino acid positions, we found hypothesis-generating signals of differential VE of Fx012-14 against first new infection strains with a match vs. mismatch to a 3D7 residue at *cs* C-terminus codon positions 322, 324, and 327 in Th2R (**Fig. S11–S14** in the Supplementary Appendix).

### Vaccine Efficacy Modification by Baseline Parasite Positivity and Intercurrent New Infection

In the PP, 11.6% of participants (7.4% in Agogo and 16.0% in Siaya) were parasite positive at baseline by microscopy and/or genotypic assay. The control-group incidence rate of the first new infection was nearly three times higher in baseline positive (3.0 per person-year at risk [PYR]) than baseline negative participants (1.2 per PYR), suggesting a correlation between baseline positivity and infection risk; therefore, we also analyzed the cumulative number of new genotypic infections (force of infection; FOI) detected after the first vaccination visit and by the month 2 visit (M2-FOI). This covariate is an aggregate proxy of individual-level infection risk due to many factors including seasonal transmission effects, local geography, susceptibility to mosquito bites, and malaria prevention use. M2-FOI could potentially confound the VE-modifying effect of baseline positivity, given that it was correlated with baseline positivity and the calendar date of the first vaccination (**Fig. S15–S16** in the Supplementary Appendix). We also accounted for such potential confounding by adjusting for the indicator of the onset of antimalarial drug treatment between the first vaccination visit and the month 2 visit (M2-mal-tx), which correlated with baseline positivity (**Fig. S17** in the Supplementary Appendix). Additional VE-modification analyses were conducted adjusting for M2-FOI and M2-mal-tx, an adjustment with minimal risk of post-randomization selection bias because vaccination had no discernible effect on M2-FOI or M2-mal-tx (**Fig. S18–S19** in the Supplementary Appendix). Adjusting for M2-FOI, M2-mal-tx, sex, and age, VE of pooled R012-14, R012-20, and Fx012-14 vs. control to prevent the first new genotypic infection in the PP was 37% (95% CI, 23 to 48%) among baseline negative and 68% (95% CI, 50 to 80%) among baseline positive participants (interaction P=0.0053) (**Fig. 4**; model M2-PP in **Fig. S20** in the Supplementary Appendix). VE modification by baseline positivity persisted when restricted to the early follow-up period between 14 days and 4.5 months after the third dose (**Fig. S21** in the Supplementary Appendix) (interaction P=0.083), a period exhibiting relatively little waning of VE. The evidence for baseline positivity as a modifier of VE was consistent across the two study sites, the individual RTS,S regimens with dosing at M0, 1, 2, and the full PP vs. sensitivity ‘third vaccination matching’ analysis (**Fig. S22–S24** in the Supplementary Appendix).

**Figure 4.**
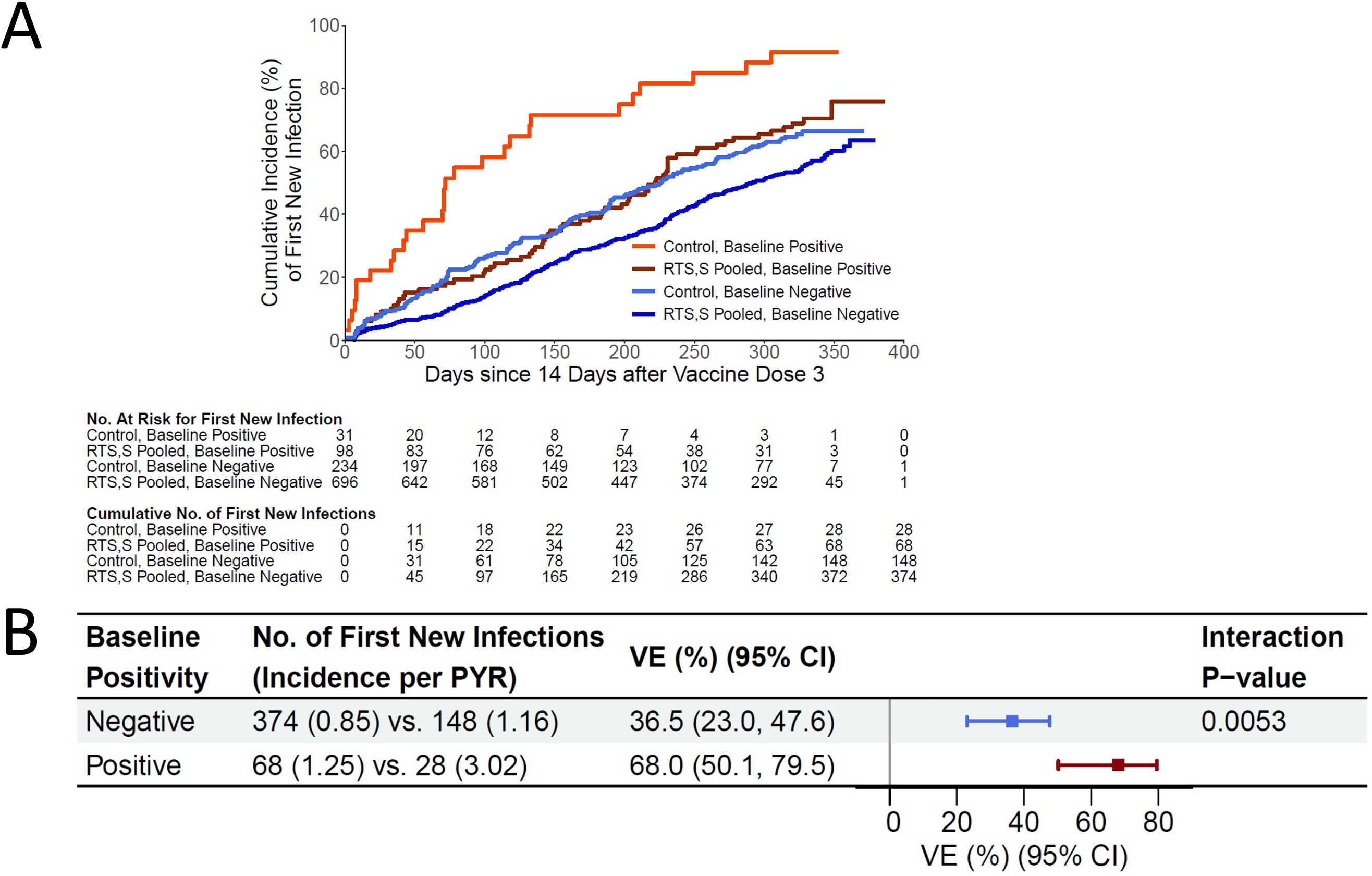
(A) Cumulative incidence and (B) vaccine efficacy against the first new genotypic infection between 14 days after month 2 through month 14 for the pooled R012-14, R012-20, and Fx012-14 RTS,S regimens vs. the control regimen for the Per-Protocol (PP) cohort by baseline malaria infection status while adjusting for month 2 force of infection (M2-FOI). PYR: person-year at risk; VE: vaccine efficacy; CI: confidence interval; No: number.

As an indicator of intercurrent malaria infection between dose 1 and dose 3, M2-FOI has a distinct interpretation compared to baseline positivity, motivating an exploratory analysis of whether M2-FOI itself modifies VE. Adjusting for baseline positivity, M2-mal-tx, sex, and age, VE of the same pooled RTS,S groups vs. control against the first new genotypic infection was 36% (95% CI, 22 to 48%) among PP participants with M2-FOI = 0 and 57% (95% CI, 39 to 69%) among PP with M2-FOI > 0 (interaction P=0.059) (**Fig. 5**; model M6-PP in **Fig. S20** in the Supplementary Appendix). VE modification evidence from a series of Cox models involving both baseline positivity and M2-FOI, including model quality assessment, is summarized in **Fig. S20** in the Supplementary Appendix for the genotypic infection endpoint. A sensitivity analysis, reported in the Supplementary Appendix, concluded that the result of VE modification by baseline positivity was robust to unmeasured confounding.

**Figure 5.**
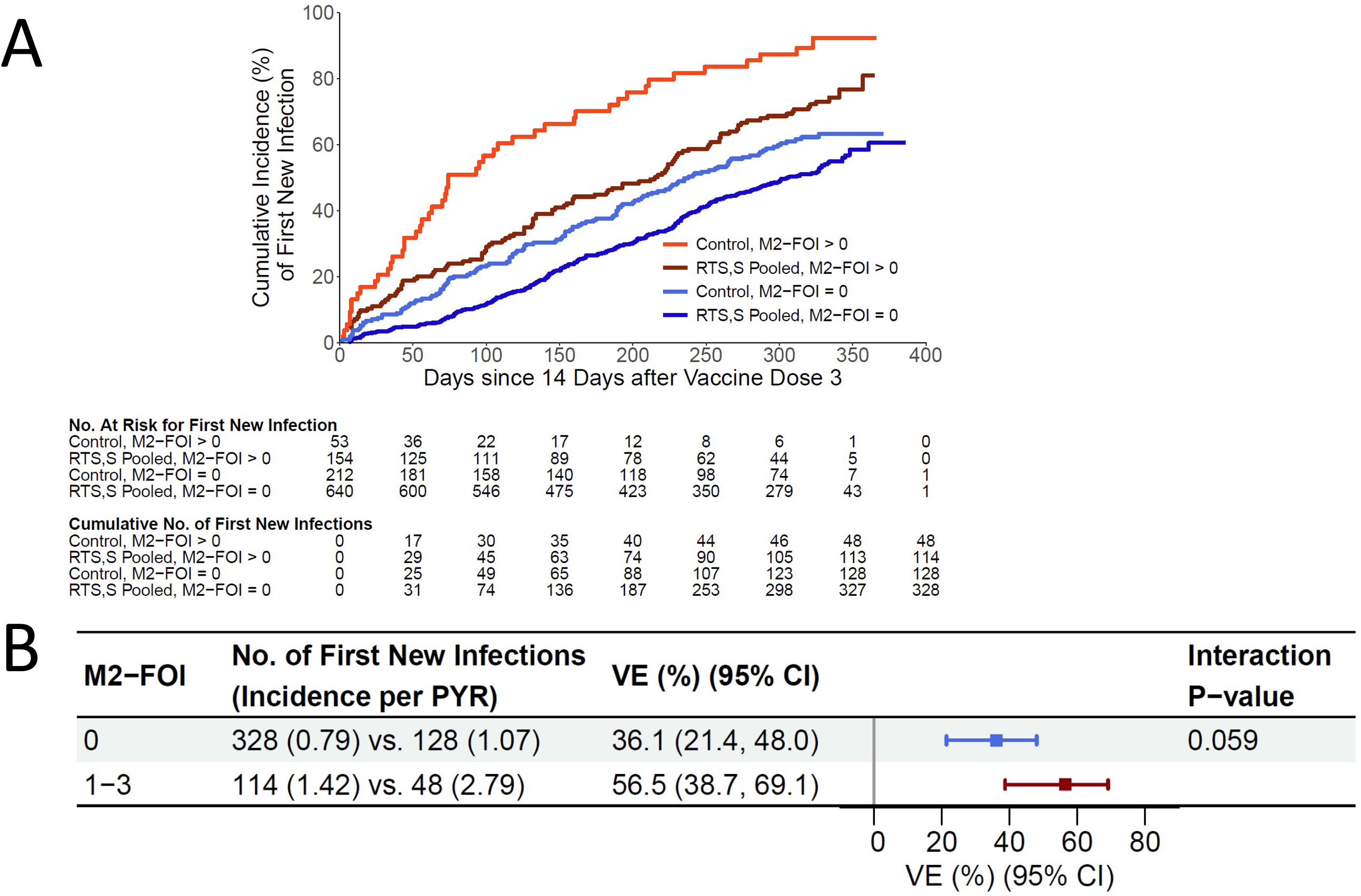
(A) Cumulative incidence and (B) vaccine efficacy against the first new genotypic infection between 14 days after month 2 through month 14 for the pooled R012-14, R012-20, and Fx012-14 RTS,S regimens vs. the control regimen for the per-protocol (PP) cohort by month 2 FOI (M2-FOI) = 0 vs. > 0. PYR: person-year at risk; VE: vaccine efficacy; CI: confidence interval; No: number; M2-FOI: month 2 force of infection.

We do not report the impact of either baseline positivity or M2-FOI on VE against the first new clinical malaria episode. Inference of this causal interaction effect is confounded by a differential propensity of first clinical episodes arising due to persistent asymptomatic infections acquired before the third vaccination, which are much more common in the baseline positive and M2-FOI > 0 subgroups (**Table S5** in the Supplementary Appendix).

## Discussion

The use of genotypically determined infection endpoints in this study has demonstrated unambiguously for the first time that RTS,S achieves some or all of the VE observed through blocking of infections before they reach the blood stage, with the RTS,S groups exhibiting a reduced number of new infections (**Fig. 2**) and a reduced risk of more highly polyclonal first infections (**Fig. 3**). While the previous analysis of parasite genotypic features we performed on specimens from the RTS,S phase 3 trial suggested this in the form of reduced COI,^4^ that study only analyzed specimens from the first cases meeting the primary clinical case definition.

The genotypically determined infection endpoints yielded findings generally concordant with the previous analysis of clinical disease endpoints with regard to the effects of RTS,S vaccine dosage and regimen. While all RTS,S dosage regimens offer significant VE, none of the regimens is superior for the follow-up period we examined. The genotypic endpoints we examined suggest lower instantaneous VE in the Fx012-14 group several months after the third dose (Fig. S7 in the Supplementary Appendix), suggesting that fractional dose regimens may offer slightly less protection than full dose regimens administered on the same schedule.

Additionally, this study demonstrates several ways in which genotypically determined infection endpoints complement clinical disease or microscopy-based infection endpoints.

Importantly, we observed that VE against the first new genotypically detected infection was higher in participants who were baseline parasite-positive (asymptomatically infected with malaria during their first vaccination). Participants who experienced more infections between their first vaccination and month 2 visit (M2-FOI) did not exhibit abrogated VE and trended towards greater protection. This suggests that active infection, and/or higher risk of infection, potentiate RTS,S VE. Because baseline infection status and M2-FOI are correlated and these features were not randomized in the study design, distinguishing their relative impacts is difficult from the current data. Further, we cannot presently distinguish whether variation in infection risk among participants is due to environmental, immunological, or other factors. However, active infections at the time of first vaccination could affect VE by the prior priming of CS-specific T-helper cells provided by natural infection, resulting in enhanced production of protective antibodies and/or a more effective cellular immune response during the liver stage. Similarly, the non-significant trend towards greater VE observed in participants with M2-FOI > 0 could be driven by repeated natural exposure to CS antigen from infectious mosquito bites as a form of heterologous prime-boost.

The observation of increased RTS,S protection in baseline-infected/M2-FOI > 0 participants is unexpected. A large number of studies have documented immunosuppressive effects of acute or asymptomatic malaria infection in various human or rodent model contexts^15–20^, and these observations have led to the hypothesis that erythrocytic-stage malaria infection at the time of vaccination may compromise the development of an efficacious immune response, measured at the level of either clinical disease^5^ or molecularly detected infection.^6^ However, a recent analysis of RTS,S efficacy in the phase 3 clinical trial found that protection against clinical malaria was unaffected by infection status during vaccination.^21^ Our work is the first to demonstrate a positive association between erythrocytic infection present at the first vaccination and VE against infection, and indicates that discordant findings in studies focusing on malaria-naive adults^6^ or mouse models^20^ may reflect fundamentally different mechanisms of pre-erythrocytic immunity development. Future studies will be required to understand this apparent discordance.

There are several important consequences of the observation of greater RTS,S VE in association with baseline parasite positivity and infection risk. First, this finding indicates that future studies of the efficacy of RTS,S and other candidate malaria vaccines may need to take into account local transmission level and/or heterogeneity in infection risk among participants, as differential VE against infection and/or clinical disease as a function of baseline infection status or infection risk could influence vaccine deployment strategy. Second, this finding motivates the inclusion of genotypic endpoints in future intervention studies to further assess the effects of baseline infection status and molecular FOI, as well as immune assays to understand the mechanism of the protective effect.^22^ Future studies should directly address the impact of baseline positivity and infection risk on VE against clinical disease, which we do not report here because the interaction effect of treatment and the potential VE modifier is confounded by differential opportunity among subgroups for the first clinical cases to be associated with asymptomatic infections acquired before the third vaccination, which may increase the risk of subsequent clinical disease.^23^ Additionally, this finding could lead designers of future vaccine and monoclonal antibody (mAb) field trials to re-evaluate the practice of diagnosing and clearing pre-existing malaria infections from subjects during enrollment, a common approach^24–26^ that may limit the approved use of an intervention to uninfected recipients if it is later licensed.

Genotyping the monthly cross-sectional samples collected from all subjects in this study has provided an unprecedented view of asymptomatic and polyclonal infection dynamics in a natural setting. The evaluation of future malaria interventions with comparable data will enable direct measurement of their potential not only to mitigate clinical cases, but also to achieve local disease elimination.

## Trademark

AS01 is a trademark owned by or licensed to the GSK group of companies.

## Data Availability

All sequencing data were submitted to the NCBI Sequence Read Archive (BioProject PRJNA983279) and are available online.

## Acknowledgements

Funding for this trial and publication was provided by GlaxoSmithKline Biologicals SA (study sponsor) and by PATH, an international public health organization, through grants awarded to PATH’s Center for Vaccine Innovation and Access by the Bill & Melinda Gates Foundation and the German Federal Ministry of Education and Research (the latter administered through the KfW Development Bank). The authors and trial partners thank the study participants and their parents/caregivers, and the RTS,S MAL-094 Study Group for their participation and support of malaria clinical research. This work was also partially supported by the National Institute of Allergy and Infectious Diseases, NIH, under award R37AI054165 (to PBG). We wish to thank the Broad Institute Genomics Platform for sample processing and data generation, in particular, Jody Camarata, Natasha Smith, and Rachael Barry. We also thank Susanna Hamilton, Chad Max, and Kalyn Hubbard for product management and Nithya Swaminathan for project management at the Broad Institute. We thank the following PATH colleagues: Scott Gregory for project management and Karen Ivinson for IRB submissions. We thank Anne Bollaerts (GSK) for data analysis and Hildegard Lemaire for sample management at GSK. Business & Decision Life Sciences platform provided editorial assistance and manuscript coordination, on behalf of GSK.

## Contributions

MJ, ML, LDM, SAdjei, TA, DA, DKB, PBYB, SKK, KO, AMS, NW, CFO, CKL, BLM, DFW, PBG, and DEN contributed to the study design and methods. AME, AK, SAdjei, TA, SAnderson, DKB, PBYB, SE, NF, JG, SKK, KO, FR, AMS, NW, OO-A, DFW, and DEN were responsible for data acquisition. MJ, AME, LL, SFS, ML, BS, NSH, DB, LDM, DA, DKB, PBYB, SKK, KO, AMS, NW, CFO, CKL, BLM, DFW, PBG, and DEN performed data analysis, and/or data interpretation. All authors read and edited the manuscript. All authors approved the final version and the decision to submit the manuscript. All authors had full access to all the data and had final responsibility for the decision to submit for publication.

