## Supplementary Appendix for "Baseline malaria infection status and RTS,S/AS01E malaria vaccine efficacy": Supp_Appendix_Text_1Nov2023.pdf

**Figure S12. Vaccine efficacy (VE) of the pooled R012-14 and R012-20 regimens vs. the**

**control regimen against the first new genotypic infection with a 3D7 residue match vs. mismatch at screened-in CS C-terminus amino acid positions in the Per Protocol cohort... 35**

**Figure S17. Distribution of the onset of antimalarial drug treatment between the first vaccination and the month 2 scheduled visit (M2-mal-tx) separately among PP baseline negative vs. baseline positive participants in the pooled control, R012-14, R012-20, and Fx012-14 groups. For percentages in parentheses, the denominator is all PP participants in the pooled control, R012-14, R012-20, and Fx012-14 groups. Spearman's correlation between baseline positivity and M2-mal-tx was 0.33 (P<0.001). PP: Per-Protocol.............40**

**Figure S21. Vaccine efficacy (VE) against the first new genotypic infection in the restricted period between 14 days and 4.5 months after the third dose for the pooled R012-14, R012-20, and Fx012-14 RTS,S regimens vs. the control regimen in subgroups of the Per-Protocol cohort defined by the baseline parasite positivity status while adjusting for the main effects of M2-FOI, M2-mal-tx, sex, age, and baseline levels of BMI and hemoglobin........................................................................................................................ 44**

**Figure S24. Vaccine efficacy (VE) against the first new genotypic infection between 14 days and 12 months after the third dose for the pooled R012-14, R012-20, and Fx012-14 regimens vs. the control regimen in the subcohort of the Per-Protocol cohort restricted to baseline positive participants and three matched baseline negative participants from the same randomization group and study site for each baseline positive participant, with matching performed on the third vaccination date. Subgroups are defined by the baseline parasite positivity status. Model adjusted for the main effects of M2-FOI, M2-mal-tx, sex, age, and baseline levels of BMI and hemoglobin.................................................................47**

LIST OF INVESTIGATORS

**Broad Institute, Cambridge, MA, USA:**
Scott Anderson, B.S., Angela M. Early, Ph.D., Sean English, B.S, Nicholas Fitzgerald, B.S., Jonna Grimsby, Ph.D., Akanksha Khorgade, M.Sc., Bronwyn L. MacInnis, Ph.D., Daniel E. Neafsey, Ph.D., Stephen F. Schaffner, Ph.D., Dyann F. Wirth, Ph.D.

**Centers for Disease Control, Atlanta, GA, USA**
Aaron M. Samuels, M.D., Nelli Westercamp, Ph.D

**GSK Vaccines, Rixensart, Belgium**
Marc Lievens, M.Sc., François Roman, M.D., Opokua Ofori-Anyinam, Ph.D., Anne Bollaerts, M.Sc.

**Emory University, Atlanta, GA, USA**
David Benkeser, Ph.D.

**Fred Hutchinson Cancer Center, Seattle, WA, USA**
Michal Juraska, Ph.D., Li Li, Ph.D., Brian Simpkins, B.A., Peter B. Gilbert, Ph.D.

**Harvard T.H. Chan School of Public Health, Boston, MA, USA**
Nima S. Hejazi, Ph.D., Dyann F. Wirth, Ph.D., Daniel E. Neafsey, Ph.D.

**KEMRI and CDC Research Public Health Collaboration, Kisumu, Kenya**
Dennis K. Bii, M.B.Ch.B, M.P.H., Simon K. Kariuki, Ph.D., Kephas Otieno, M.Sc., Aaron M. Samuels, M.D.

**Kwame Nkrumah University of Science and Technology/ Agogo Presbyterian Hospital, Agogo, Ghana**
Samuel Adjei, M.B., Ch.B., D.T.M., Tsiri Agbenyega, M.B., Ch.B., Ph.D., Daniel Ansong, M.B., Ch.B., F.W.A.C.P., Patrick B.Y. Buabeng, M.B.A.

**PATH Malaria Vaccine Initiative, Washington, DC, USA**
Laina D. Mercer, Ph.D., Christian F. Ockenhouse, M.D., Ph.D., Cynthia K. Lee, Ph.D.

**University of Washington, Seattle, WA, USA**
Qi Wang, B.Eng., Peter B. Gilbert, Ph.D.

AUTHOR CONTRIBUTIONS

CFO, DEN, DFW, CKL, MJ, LDM, PBG, BLM, DAB, LDM, and ML designed the study and analysis plan.

AMS, DKB, SKK, KO, SA, TA, FR, and DA executed the trial and collected the samples.

SE, NF, JG designed and developed the laboratory processes for genotyping

SA designed and developed the laboratory automation for genotyping

AK, AME, SFS filtered the genotyping data.

MJ, AME, SFS, LL, CM, BS, NH, DAB, and QW analyzed the data.

CKL, DEN, OOA, BLM, PBG, MJ, AME, LDM, ML and DFW provided project oversight and supervision.

MJ, AME, DEN, PBG, and DFW vouch for the data and the analysis.

MJ, AME, DEN, PBG, and DFW decided to publish the paper.

DEN, MJ, LL, and AME wrote the first draft.

### SUPPLEMENTARY METHODS

#### SAMPLE RECEIPT AND REGISTRATION

FTA (Flinders Technology Associates) blood spot cards were received by the Broad Institute GenomiCS Platform (GP), tagged with GP sample specific barcodes, both GP and GSK barcodes were registered in the GP laboratory information management system (LIMS), and cards were stored in desiccator cabinets at room temperature until further processing. Samples were held in a room requiring a Broad Institute identification card for access.

#### DNA EXTRACTION AND QUANTITATION

Genomic DNA extraction was performed in batches of 96 *Whatman* FTA cards, including one blank control FTA card. For each FTA card 7 disks were punched out of the blood spot, using an automated laser guided hole puncher, into a distinct well of 96 well plates. Genomic DNA was extracted from the punches using *QIAamp* 96 DNA Blood Kit (Qiagen) using the Bravo (Agilent) for automated liquid handling. DNA samples were registered in LIMS and stored in barcoded tubes. DNA concentration of each sample was quantified using standard automated *PicoGreen* quantification. All steps of the process were tracked in the LIMS.

##### Polymerase Chain Reaction (PCR)

Two *Plasmodium* PCR amplicons, "*cs* C-terminus" and "*sera2*," were amplified in 36,080 samples. The *cs* C-terminus amplicon captures the polymorphic C-terminus T-cell epitope region of *cs.* The *sera2* amplicon is not located within *cs*, but captures part of a comparably polymorphic antigen sequence from elsewhere in the *P. falciparum* genome. The *sera2* locus was used as a control for sieve analysis. Final haplotype calls from the cs C-terminus and *sera2* amplicons were all used to estimate complexity of infection (COI) for individual samples. Full amplicon sizes (including adapter sequence, flow cell attachment sequences and indices) were

400 and 371 for the *cs* C-terminus and *sera2* amplicons, respectively. Plasmodium portions of these amplicons were 333 and 304 bp for *cs* C-terminus and *sera2* amplicons, respectively. The formal *P. falciparum* 3D7 gene IDs and nucleotide coding sequence (CDS) coordinates for these amplicons are:

*cs* C-terminus (PF3D7_0304600): CDS bp 858-1190

*sera2* (PF3D7_0207900): CDS bp 72 - 357

The *cs* C-terminus and *sera2* amplicons were sequence-ready constructs and did not require further library construction after PCR. These PCRs were carried out in two reactions. Round 1 PCR primers contained *Plasmodium* sequence and *Illumina* adapter sequences while round 2 PCR primers were "tailing" primers, containing some overlap of the *Illumina* adapter sequence, flow cell attachment sequences, and an eight bp index on the reverse primer between the adapter sequence and flow cell attachment sequence (primer sequences below).

First-round PCRs for *cs* C-terminus and *sera2* were carried out using the *Hot Star Plus DNA Polymerase Ki*t (Qiagen). Reactions consisted of 5 µl DNA at ~1.5 ng/ml, 10 µl mixed F/R primer (1.0 mM for cs C-terminus, 2.0 mM for SERA), 2µl 10X buffer, 0.8 µl 25 mM MgCl2, 0.16 µl dNTPs (100 mM dNTP mix, Agilent Technologies), 0.08 µl *HotStar Taq* (5U/µl), 3.96 µl nuclease free water. Thermal cycling consisted of 95˚C for 5 min, 30 cycles of [94˚C 30 sec, 60˚C 30 sec, 72˚C 1 min] and 3 min at 72˚C. Second-round PCRs for *cs* C-terminus and *sera2* consisted of 2 µl of PCR1 product, 2.16 µl nuclease free water, 11.72 µl Pfu Buffer, 0.12 µl Pfu DNA polymerase and 10 µl mixed F/R indexed primer (1.6mM). Second-round PCR thermal cycling for *cs* C-terminus consisted of 50˚C for 2 min, 70˚C for 20 min, 95˚C for 10 min, 5 cycles of [95˚C 15 sec, 60˚C 30 sec, 72˚C 1 min], 1 cycle of [95˚C 15 sec, 80˚C 30 sec, 60˚C 30 sec, 72˚C 1 min], 4 cycles of [95˚C 15 sec, 60˚C 30 sec, 72˚C 1 min], 1 cycle of [95˚C 15 sec, 80˚C 30 sec, 60˚C 30 sec, 72˚C 1 min], 4 cycles of [95˚C 15 sec, 60˚C 30 sec, 72˚C 1 min], 5 cycles of [95˚C 15 sec, 80˚C 30 sec, 60˚C 30 sec, 72˚C 1 min]. Second-round PCR thermal cycling for *sera2* consisted of 95˚C for 5 min, 9 cycles of [94˚C 30 sec, 60˚C 30 sec, 72˚C 1 min] and 72˚C for 3 min.

Samples were grouped into batches that were processed sequentially as they were received at the Broad Institute. In total, 127 batches were processed and sequenced, each composed of up to 372 extracted blood spot samples loaded onto four 96-well plates. Each 96-well plate included two negative control samples (a water-only PCR control and a blank *Whatman* card punch extraction control) and 1-2 positive control samples. Batches 1-62 contained a mixture of two custom plasmids (Invitrogen/Thermo Fisher Scientific) as the positive control. These plasmids contained *cs* and *sera2* sequences with variants that have never been sampled in natural populations, making them easy to distinguish from true amplicons. One plasmid was previously used and described in Early *et al*. (2019).[1] The second was constructed using the same protocol. Due to repeated PCR failures with the plasmids, batches 63-127 additionally included genomic *P. falciparum* 3D7 DNA purchased from ATCC. For each batch, a sampling of amplicon products was visually inspected using a *Lab Chip* GX II *Caliper* Instrument (Perkin Elmer).

Indices for sample identification were assigned during PCR so that, within a batch, the same sample was assigned the same index for *cs* C-terminus and *sera2* amplicons. For each batch of 384 samples, indexed *cs* C-terminus PCR2 products were pooled by volume, as were *sera2* PCR2 products. These 2 amplicon pools were purified using a 0.7X solid-phase reversible immobilization (SPRI) cleanup with *Agencourt Ampure* XP beads (Beckman Coulter). For each batch, positive and negative control samples were then assessed and quantified on a *BioAnalyzer* (Agilent Technologies) and *cs* C-terminus and *sera2* products were normalized and pooled together. For automated PCR set-up, pooling, LIMS tracking and messaging, a Bravo Automated Liquid Handling Platform (Agilent Technologies) was used. To avoid PCR contamination, automated setup of PCR2 included tip piercing of PCR1 plate and primer plate covers to avoid amplicon spray going into nested PCR. In addition, PCR workspaces were decontaminated with *DNA ZAP* (Ambion) and negative control wells were visually inspected on an Agilent *BioAnalyzer*.

PRIMER SEQUENCES

Round 1 PCR primers (*cs* C-terminus, sera2; *Plasmodium* sequence in bold; X indicates positions of sample-specific barcode sequences):

cs_C-terminus_Round_1_Forward:
ACACTCTTTCCCTACACGACGCTCTTCCGATCT**TTAAGGAACAAGAAGGATAATACCA**
cs C-terminus_Round_1_Reverse:
GTGACTGGAGTTCAGACGTGTGCTCTTCCGATCT**AAATGACCCAAACCGAAATG**

sera2_Round_1_Forward:
ACACTCTTTCCCTACACGACGCTCTTCCGATCT**TACTTTCCCTTGCCCTTGTG**
SERA_Round_1_Reverse:
GTGACTGGAGTTCAGACGTGTGCTCTTCCGATCT**CACTACAGATGAATCTGCTACAGGA**

Round 2 PCR Primers (*cs* C-terminus, *sera2*):

Round2_Forward:
AATGATACGGCGACCACCGAGATCTACACTCTTTCCCTACACGACGCTCTTCCGATCT
Round 2_Reverse:
CAAGCAGAAGACGGCATACGAGATXXXXXXXXGTGACTGGAGTTCAGACGTGTGCTCTTCCGATCT

**SEQUENCING**

One MiSeq run (2x250 bp paired end with standard sequencing primers) was carried out for each sample batch using standard methods (V2 sequencing chemistry). PhiX library, derived from the well-characterized and small PhiX genome, was spiked in at 15% to add diversity for improved cluster imaging. Sequencing data were demultiplexed to create sample specific sequencing read BAM files. All steps were tracked in the LIMS. Raw Bam files are available on the NCBI Sequence Read Archive database (BioProject PRJNA983279).

#### HAPLOTYPE CALLING

To call haplotypes, we used the PASEC pipeline, following the best practices established in Early *et al* (2019).[1] Sequencing reads were demultiplexed and the overlapping 250-bp read pairs were merged using FLASH[2] and aligned with BWA-MEM[3] v0.7.12-r1039 to the *P. falciparum* 3D7 and plasmid sequences from the amplicon regions. We discarded reads if they contained uncalled bases, had a quality score <20, or if the merged read did not span the entire amplicon region. The remaining reads were clustered on a per-sample basis according to their haplotype sequence: haplotypes with a 1-bp difference were collapsed into a single majority consensus sequence if the intra-sample abundance ratio was >8:1. Following this clustering step, two poly-T runs in cs were masked in all sequences and disregarded in downstream analysis as they are unreliably sequenced with *Illumina* technology (amplicon positions 63-72 and 251-257). We clustered into a single haplotype any reads that were identical outside of these homopolymer regions. As discussed more fully below, we analyzed the data using two separate intrasample read-support thresholds (50 and 325). Haplotypes with intra-sample read support below these values were masked from further analysis. For each remaining haplotype, we called single nucleotide variants in relation to the 3D7 sequence and calculated the total number of differences at the nucleotide and amino acid levels (Hamming distances). The translated peptide coordinates correspond to amino acid positions 294-388 (CS) and 34-118 (SERA2).

#### HAPLOTYPE FILTERING

We implemented several layers of haplotype filtering to control for PCR/sequencing artifacts. At the sample level, we removed any haplotype sequence that was represented by fewer than 1% of reads, that contained an out-of-frame indel, or that started at an alternate position as these have a high probability of being PCR or sequencing errors. At the study level, we assessed patterns of population-level variation. We removed indels that were found in only a single haplotypic background, which left two in-frame indels in *sera2* in the downstream analysis. We analyzed the distribution of Hamming distances and found a bimodal distribution; the majority of distinct haplotypes (96.9% of pre-filtered haplotypes represented by 99.4% of recovered sequences) had a nucleotide Hamming distance <13 for *cs* and <10 for *sera2*. A small set of haplotypes (0.16% of recovered *cs* sequences and 1.2% of recovered *sera2* sequences) greatly exceeded these values (median nucleotide Hamming distance = 191 for *cs* and 27 for *sera2*). We found that this latter group contained chimeric sequences joining a natural sequence and a positive control plasmid sequence. We therefore filtered out any haplotype with a nucleotide Hamming distance >=13 for *cs* and >=10 for *sera2.*

Finally, we considered the co-occurence of haplotypes. Haplotypes were categorized as (1) those seen only in the presence of one or more other haplotypes, and (2) those that are seen at least once by themselves. Haplotypes in category (1) include those we wished to filter out, while those in category (2) are presumed to be the ones that we want to preserve. For each haplotype, we calculated the geometric mean sequencing depth for all instances in the dataset. We then calculated the number of haplotypes that were filtered from each category by a minimum coverage threshold. The number of additional haplotypes removed with each increment in the threshold is shown in the two figures separately for the two amplicons. Based on these distributions, the filtering step was chosen to be: any haplotype seen only in association with other haplotypes, and with a mean geometric coverage below the threshold was eliminated, where the threshold was set to be 5000 for each amplicon.

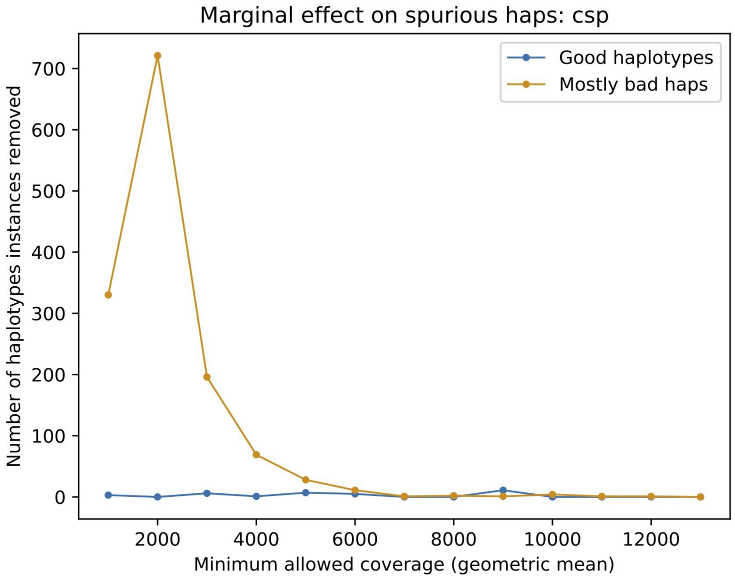
Marginal effect on spurious haps: csp
Good haplotypes
Mostly bad haps
Number of haplotypes instances removed
0
100
200
300
400
500
600
700
2000
4000
6000
8000
10000
12000
Minimum allowed coverage (geometric mean)

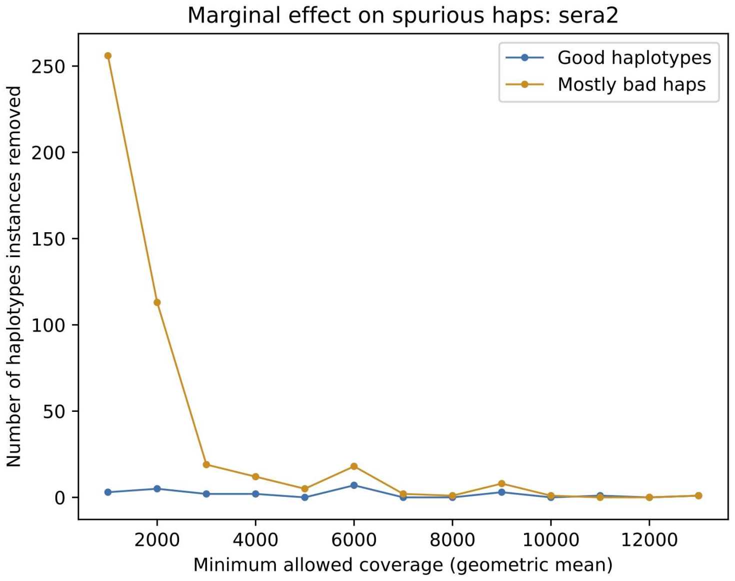
Marginal effect on spurious haps: sera2
Good haplotypes
Mostly bad haps
Number of haplotypes instances removed
0
50
100
150
200
250
2000
4000
6000
8000
10000
12000
Minimum allowed coverage (geometric mean)

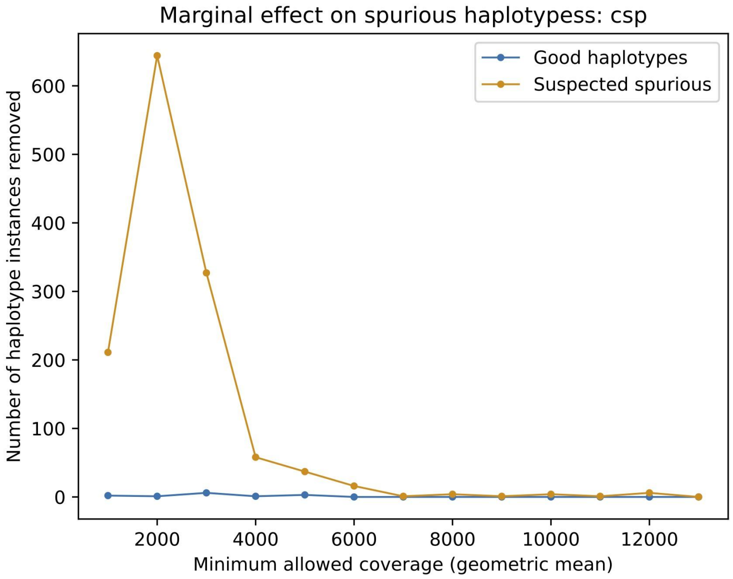

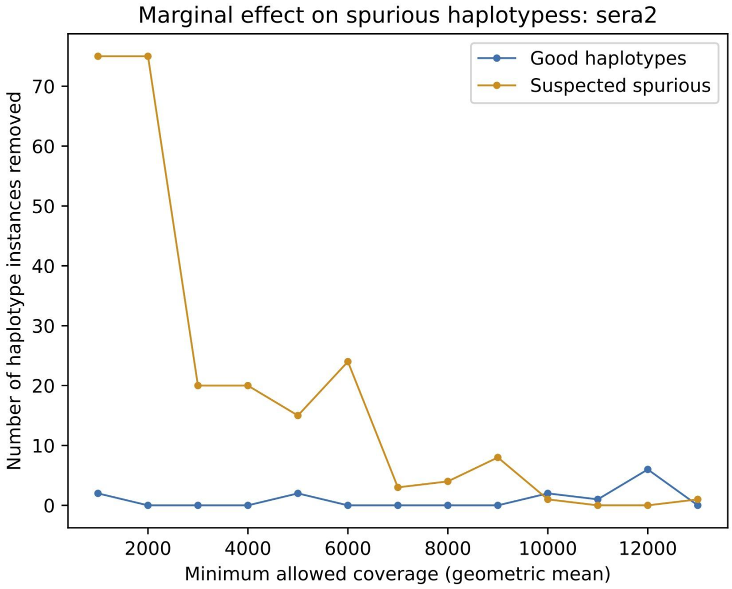

#### DETERMINATION OF INTRA-SAMPLE READ SUPPORT THRESHOLD

We set an initial intra-sample read support threshold of ≥50 reads per haplotype based on data generated in Early *et al* (2019)[1] and from 128 control samples sequenced in the initial stages of this study. After completing data generation, we had greater power to detect low levels of contamination and reassessed this threshold, ultimately raising it to 325 reads per haplotype. Contamination signals were expected to diverge from what was seen in previous amplicon studies (for example, Neafsey & Juraska, *et al*, 2015[4] and Early, *et al*, 2019[1]) because both parasite-positive and parasite-negative samples were included in the PCR and sequencing performed in this study.

Determining the optimal threshold was a two-step process. First, we generated a numerical estimate of the number of false positives being generated by contamination. We calculated an empirical probability of haplotypes co-occurring on a plate, based on haplotypes with mid-range coverage depth, assuming that these were neither "donors" nor "recipients" of contamination. This estimate is shown by the orange line in the figure below. We assumed that excess above this rate represents false positives, with everything else being true positives. We then increased the minimum coverage threshold and calculated the true and false positive/negative rate for each threshold.

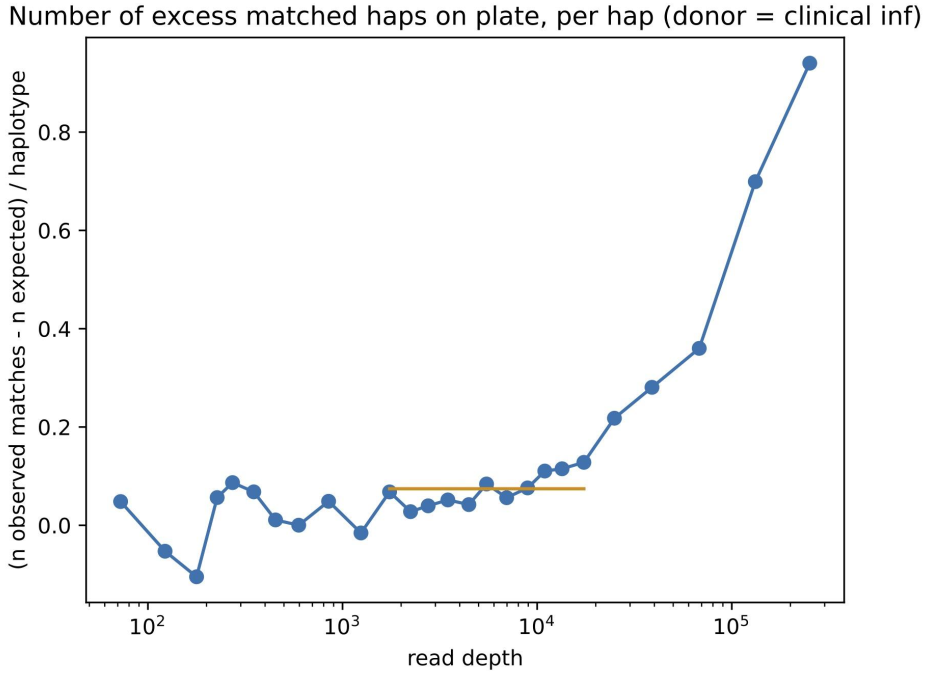

To actually set the new threshold, we used the F1 score, which is given by

$$F1\ Score = 2 \times \frac{recall \times precision}{recall + precision}.$$

F1 was maximized at a threshold of 325 reads. Accordingly, we set the threshold at that value.

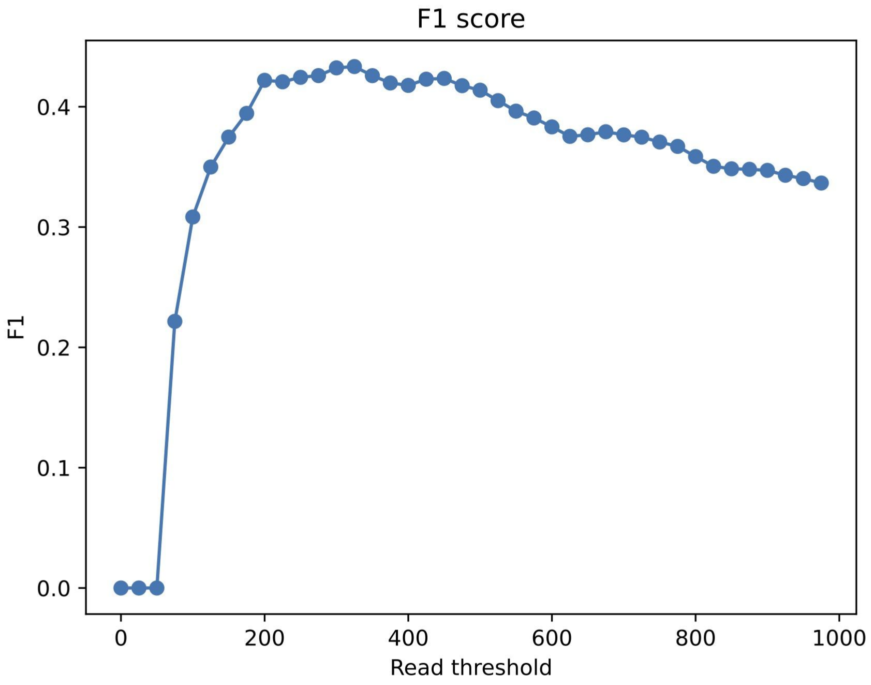

#### NEW INFECTION DEFINITION

A new infection is declared on a date for a subject who has at least one haplotype that has not been seen in that subject on an earlier date. Thus, a new infection can be identified even if a new haplotype is seen only in one amplicon. A new infection is also declared based on the reappearance of a haplotype, provided if at least three intervening visits occurred during which that haplotype was not seen in the subject. For this purpose, both scheduled and unscheduled visits are counted, but multiple visits on the same day are treated as a single visit.

This definition was set based on preliminary data. With the full dataset in hand, we were able to evaluate the effect of the three-visit gap. To do so, we calculated the rate of seeing the same haplotype again in a subject as a function of the number of intervening visits for which the haplotype was not detected. We then compared that rate to the estimated probability of seeing the same haplotype again by chance due to re-infection with a new parasite strain carrying the same haplotype. The latter was calculated based on the haplotype's frequency in the population (at that study site) and on the number of haplotypes seen in the later visit. Note that the expected re-infections are biased upward for small gap sizes. This is because it is based on the number of observed haplotypes, and that number is on average higher shortly after malaria has been observed in that subject; the asymptote of the expected curve gives a better sense of the true expected re-infection rate.

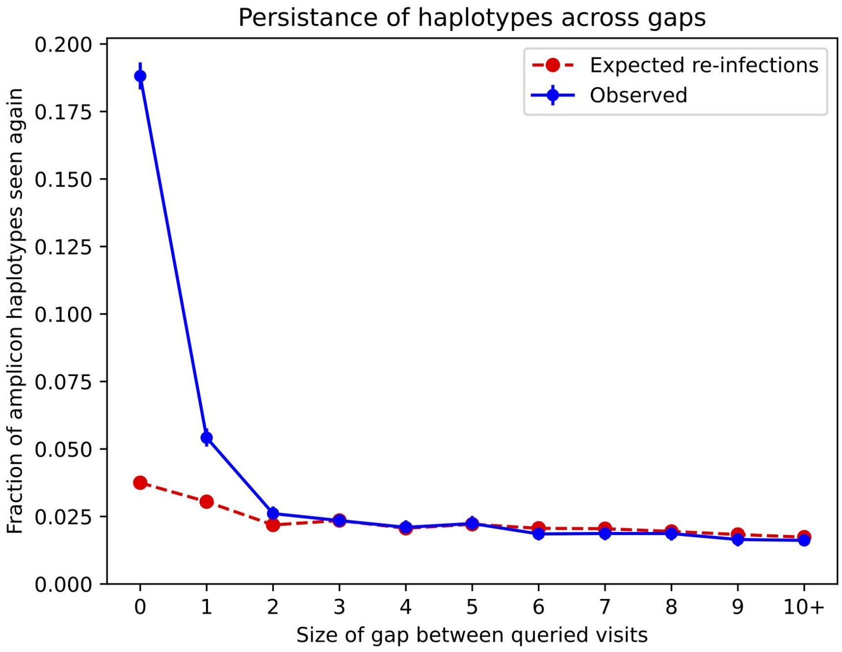

#### STATISTICAL METHODS FOR ASSESSING VARIATION IN VACCINE EFFICACY (VE) WITH SEQUENCE FEATURES OF INFECTING PARASITES (SIEVE ANALYSIS)

It was of interest to assess whether and how VE against the first new infection depended on genotypic characteristics of infecting parasites. First, we pre-specified in a treatment-blinded manner a set of candidate haplotype-level amino acid sequence features and sample-level parasite complexity measures (described in the Statistical Analysis Plan included in the Supplementary Appendix). Next, for each treatment comparison including that of pooled R012-20, R012-14, and Fx012-14 groups vs. control, we screened out binary haplotype features lacking ≥ 25 treatment-pooled first new infections representing each level of the binary feature to maximize statistical power. Screened-in amplicon-specific features exhibited missing values because haplotype detection at one but not the other amplicon was sufficient for calling a new genotypic infection. We assessed variation in VE by amplicon-specific features using augmented inverse probability weighting methods[5,6] that accommodate missing sequences. Logistic regression models were employed for estimating the probability of a missing feature and constructing the augmentation term. Complete-case versions of these methods[7] were applied for analyzing COI, which exhibited no missingness as an aggregate measure across both amplicons. Many first new infections were polyclonal (COI>1); to account for haplotype multiplicity per amplicon, analyses of haplotype-level features were performed on data sets comprising a single randomly selected haplotype from a sample, with multiple outputation[8] used to aggregate results as applied previously.[4]

#### HYPOTHESIS TESTING MULTIPLICITY ADJUSTMENT FOR SIEVE ANALYSIS

Multiple hypothesis testing correction for “sieve effect” tests of differential VE across levels of a haplotype feature was applied separately within the following sets of features: 3D7 haplotype match binary features, 3D7 Hamming distances, and within-host complexity measures other than COI.

#### SUPPLEMENTARY STATISTICAL DATA ANALYSIS

##### SENSITIVITY ANALYSIS TO QUANTIFY THE ROBUSTNESS OF THE VACCINE EFFICACY MODIFICATION RESULTS TO UNMEASURED CONFOUNDING

For each level *a* of baseline parasite positivity status (BPPS), let VE(*a*) be causal vaccine efficacy under a hypothetical assignment of every trial participant to BPPS value *a*, where *a* = 0 indicates negative and *a* = 1 indicates positive. Following Mathur et al. (2022),[9] we define a causal VE interaction parameter by CxR = (1 – VE(1))/(1 – VE(0)), where CxR departing from one indicates causal VE interaction. In the main article, Cox model analyses were conducted to estimate the statistical parameter StatR corresponding to CxR, StatR = (1 – VEph(1))/(1 – VEph(0)), which is the ratio of vaccine vs. placebo hazard ratios controlling for all measured potential confounding variables. If all confounders were included and the Cox model was correctly specified, then these statistical analyses showing StatR < 1 indicate the causal inference CxR < 1.

However, unmeasured confounding could make CxR = 1 (no causal VE interaction) even though StatR < 1. Therefore, we apply the Mathur et al. (2022)[9] sensitivity analysis method to quantify how strong unmeasured confounding would have to be to explain away an observed causal association, that is, to determine the strength of unmeasured confounding for the observed VE interaction result to not be causal. This is quantified by two E-values: the E-value for an effect heterogeneity point estimate (E-value.pt.est), and the E-value for the upper confidence limit (E-value.ul). E-value.pt.est is the minimum magnitude that at least one of the four within-stratum confounding strengths (defined in Table S1) must have such that fully controlling for confounding would have shifted the estimate of StatR to the null (i.e. CxR = 1). E-value.ul is the same except such that fully controlling for confounding would have shifted the upper 95% confidence limit of CxR to include one. Large values of E-value.pt.est and E-value.ul above one support robustness of causal interaction, with greater values implying greater robustness. Attaining E-value.ul > 1 is a requirement for credibility of the causal inference.

The E-values were computed for each of the three interaction analyses for the new malaria infection endpoint considered in the main article: (1) For BPPS and VE in the total vaccinated cohort (TVC) through 20 months; (2) for BPPS and VE in the Per Protocol (PP) cohort through 14 months; and (3) for Month 2 Force of Infection ( M2-FOI) and VE in the PP cohort through 14 months.

Table S2 shows the results. For the first two rows, the fact E-value.ul > 1 supports some robustness of the finding of a causal interaction, whereas for the third row E-value.ul = 1 does not. To understand whether the E-values (1.873, 1.416) for row 1 and (1.897, 1.404) for row 2 provide high robustness, we follow recommended practice[10] to estimate the confounding strengths of all of the observed baseline potential confounders that have been considered (age, sex, body mass index (BMI), hemoglobin, malaria treatment, study site), both individually and jointly. If these observed confounding strengths tend to be lower than the E-values, it helps support a robust result. Tables S3 and S4 show the confounding strengths of all the observed baseline potential confounders that were included in the analyses (1) and (2), respectively. In those two analyses, the E-values for the effect heterogeneity point estimate are greater than 21 of the 24 (91.7%) observed confounding strengths and greater than 20 of the 24 (87.5%) of the observed confounding strengths of controlled potential confounders, respectively. We note that confounding strengths 1 and 2 of the indicator of the antimalarial drug treatment between the first vaccination and the month-2 scheduled visit are larger than the rest of confounding strengths presented in Tables S3 and S4. This may be due to the fact that baseline parasite positive participants were more likely to receive drug treatment between the first vaccination and the M2 visit. It may be considered implausible that unmeasured confounders could have such confounding strengths. In addition, given the control of six known confounders, it arguably is implausible that unmeasured confounders are strong enough to attain the confounding strengths that are larger than the E-values for point estimates. Based on this result, the inferences of a causal VE interaction in analyses (1) and (2) is robust to unmeasured confounding.

Table S1. Definition of the four within-stratum confounding strengths for a binary unmeasured confounder.[9]

| |
|---|
| For the vaccine arm, the relative risk that the unmeasured confounder takes value 1 for the BPPS = 1 subgroup vs. the BPPS = 0 subgroup, within levels of the measured confounders included in the analysis |
| Same as 1. for the placebo arm |
| For the vaccine arm, the relative risk of new malaria infection for the unmeasured confounder taking value 1 vs. 0, within levels of the measured confounders included in the analysis |
| Same as 3. for the placebo arm |

BPPS: Baseline parasite positivity status

Table S2. E-values for the point estimate and for the upper 95% confidence limit for estimation of causal VE interaction.

| Analysis | StatR point estimate (95% CI) | E.value.pt.est | E-value.ul |
|---|---|---|---|
| BPPS modifier of VE over 0-20 months in TVC | 0.493<br>(0.315, 0.771) | 1.873 | 1.416 |
| BPPS modifier of VE over 2.5-14 months in PP | 0.481<br>(0.297, 0.779) | 1.897 | 1.404 |
| M2-FOI modifier of VE over 2.5-14 months in PP | 0.678<br>(0.455, 1.011) | 1.550 | 1.000 |

BPPS: Baseline parasite positivity status; CI: confidence interval; VE: vaccine efficacy; E.value.pt.est: E-value for an effect heterogeneity point estimate; E-value.ul: E-value for the upper confidence limit; StatR: statistical parameter corresponding to the causal VE interaction parameter: CxR = (1 – VE(1))/(1 – VE(0))

Table S3. Confounding strengths of all the observed baseline potential confounders that were included in the analysis with BPPS as the modifier of VE over 0 to 20 months in the TVC.

| Observed confounder | Confounding strength 1 | Confounding strength 2 | Confounding strength 3 | Confounding strength 4 |
|---|---|---|---|---|
| Age | 1.207 | 1.051 | 1.168 | 1.125 |
| Sex | 1.125 | 1.126 | 1.159 | 1.202 |
| BMI | 1.024 | 1.006 | 1.169 | 1.125 |
| Hemoglobin | 1.688 | 1.679 | 1.279 | 1.232 |
| Antimalarial drug treatment | 3.937 | 4.321 | 1.356 | 1.177 |
| Site (Agogo, Siaya) | 1.363 | 1.398 | 1.642 | 1.244 |

BMI: body mass index; BPPS: baseline parasite positivity status; TVC: total vaccinated cohort; VE: vaccine efficacy

Table S4. Confounding strengths of all the observed baseline potential confounders that were included in the analysis with BPPS as the modifier of VE over 2.5-14 months in the ATP

| Observed confounder | Confounding strength 1 | Confounding strength 2 | Confounding strength 3 | Confounding strength 4 |
|---|---|---|---|---|
| Age | 1.204 | 1.052 | 1.714 | 1.402 |
| Sex | 1.104 | 1.142 | 1.173 | 1.302 |
| BMI | 1.098 | 1.053 | 1.307 | 1.143 |
| Hemoglobin | 1.718 | 1.647 | 1.667 | 1.445 |
| Antimalarial drug treatment | 4.279 | 4.026 | 1.510 | 1.430 |
| Site (Agogo, Siaya) | 1.425 | 1.452 | 1.976 | 1.875 |

ATP: according to protocol; BMI: body mass index; BPPS: baseline parasite positivity status; VE: vaccine efficacy

*Details on confounding strengths*

Let X, Y, and Z be the indicators for baseline parasite positivity, whether an individual had new malaria infection during the follow-up, and vaccination, respectively. Specifically, $I(Y = 1) = I(T \leq t)$, where T is the new-infection time of an individual and t is the ending time point of the follow-up, set at M20 (606 days) and M14 (431 days) for the TVC and ATP analyses, respectively. Let U denote the uncontrolled confounder(s) within stratum Z = z. The sensitivity parameters $RR_{UY|Z=z, X=x}$ and $RR_{UY|X=x}$ are given by (Mathur et al., 2022)[9]

$$RR_{UY|Z=z,X=x} = \frac{\max_{u} E[Y|Z=z, X=x, U_x=u]}{\min_{u} E[Y|Z=z, X=x, U_x=u]}, z \in \{0,1\},$$
$$RR_{UY|X=x} = \max\{RR_{UY|Z=0,X=x}, RR_{UY|Z=1,X=x}\}, x \in \{0,1\}.$$

In addition, sensitivity parameters $RR_{ZU|X=1}$ and $RR_{ZU|X=0}$ are defined as (Mathur et al., 2022)[9]

$$RR_{ZU|X=x} = \max_{u} \left\{\frac{P(U_x=u|X=x, Z=1)}{P(U_x=u|X=x, Z=0)}\right\}.$$

To estimate the confounding strengths of all the observed baseline potential confounders that have been considered individually, we consider the median dichotomization of each controlled baseline confounder that is continuous. Then, we use Kaplan-Meier estimators and empirical frequency estimators to estimate $E[Y \mid Z = z, X = x, U_x = u] = P(T \leq t \mid Z = z, X = x, U_x = u)$ and $P(U_x = u \mid X = x, Z = z)$, respectively, and all confounding strengths can hence be estimated.

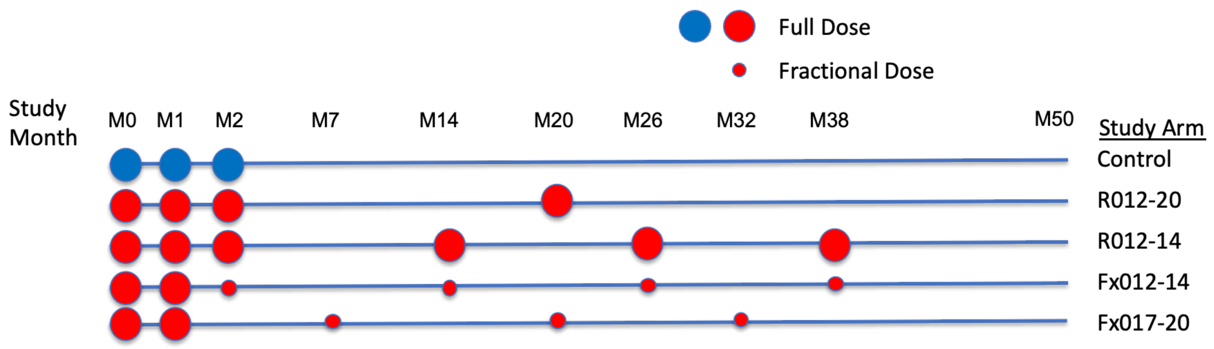

**Figure S1. Vaccination dosage and schedule.**
Samples were collected passively during febrile clinic visits and cross-sectionally at monthly intervals through study month 20 (M20).

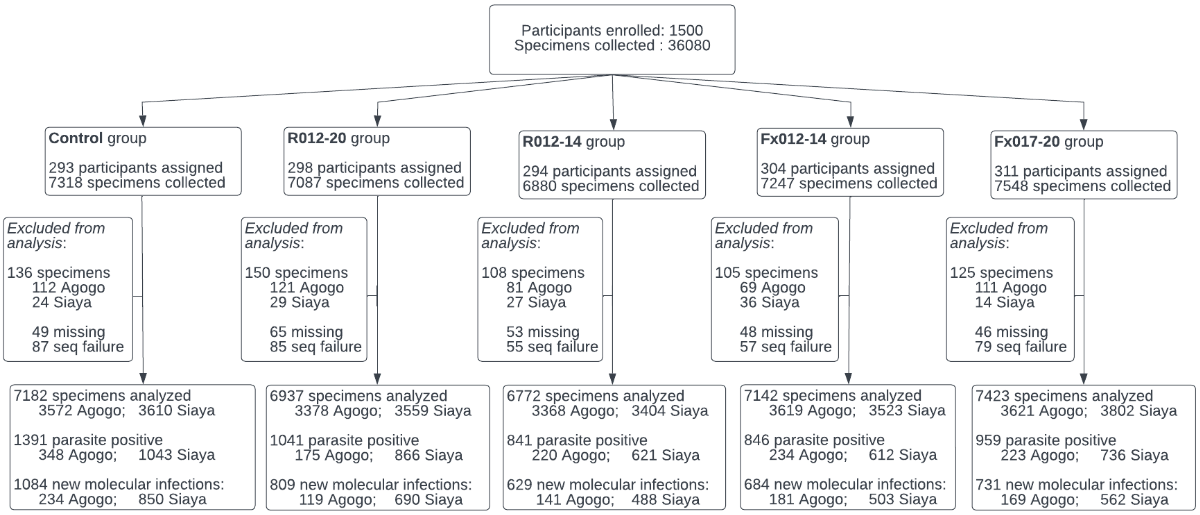

**Figure S2. Specimen collection and genotype data generation by study group in the ES cohort.**
ES: Exposed Set; seq: sequencing

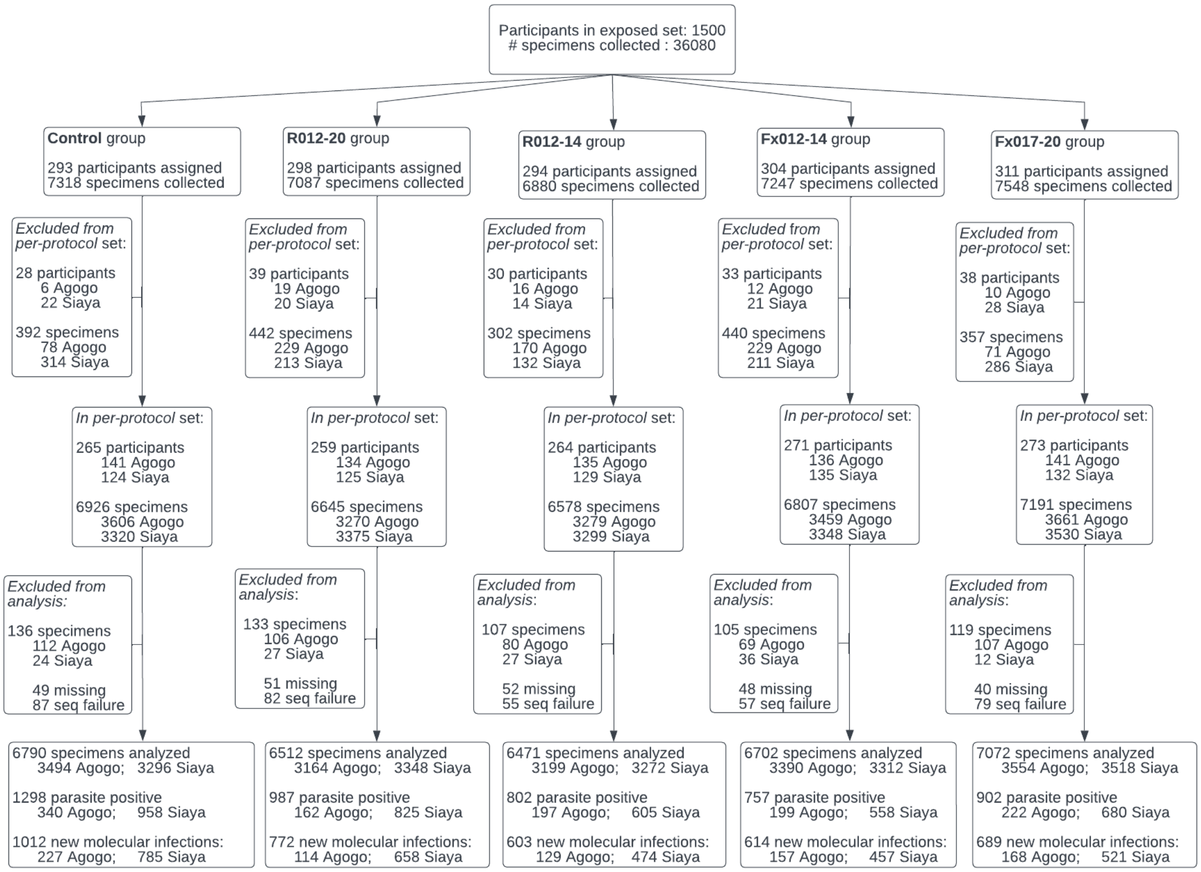

**Figure S3. Specimen collection and genotype data generation by study group in the PP cohort.**
PP: Per-Protocol; seq: sequencing

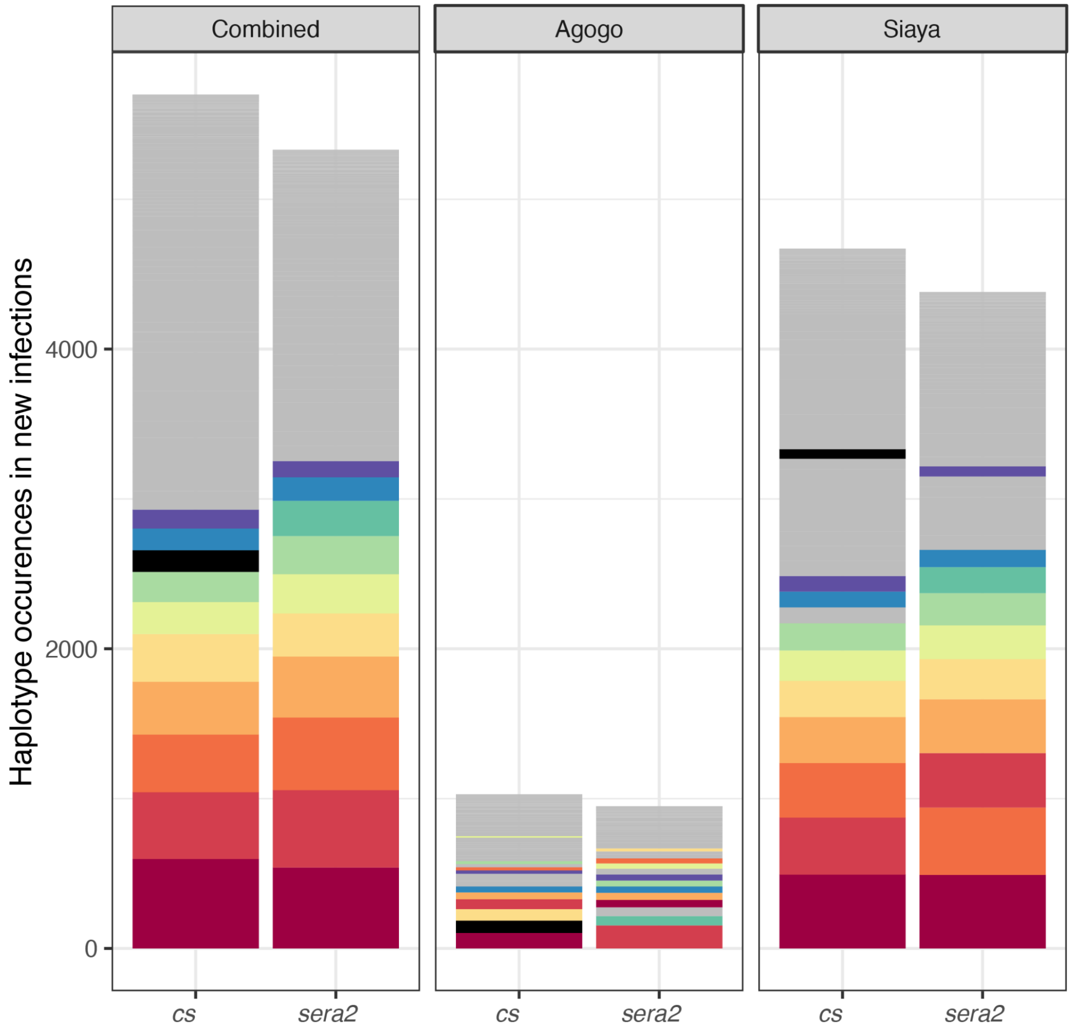

**Figure S4. Frequency of cs and *sera2* haplotypes retrieved from new infections.** Both the *cs* and *sera2* amplicon regions are highly diverse with no single haplotype accounting for more than 17.4% of amplicons at a single site. In total, we observed 178 and 160 distinct haplotypes for *cs* and *sera2*, respectively. The counts for the 10 most frequent haplotypes are colored above, with a single color representing the same amplicon sequence across sites. No relationship exists between the *cs* and *sera2* coloring. The cs sequence matching the 3D7 reference is colored black; it ranked 2 in Agogo, 20 in Siaya, and 8 in the combined data set. The *sera2* sequence matching the 3D7 reference was at a low frequency and is not notated in the figure; it appeared in 11 new infections from Agogo and was never sampled in a new infection from Siaya.

**A**

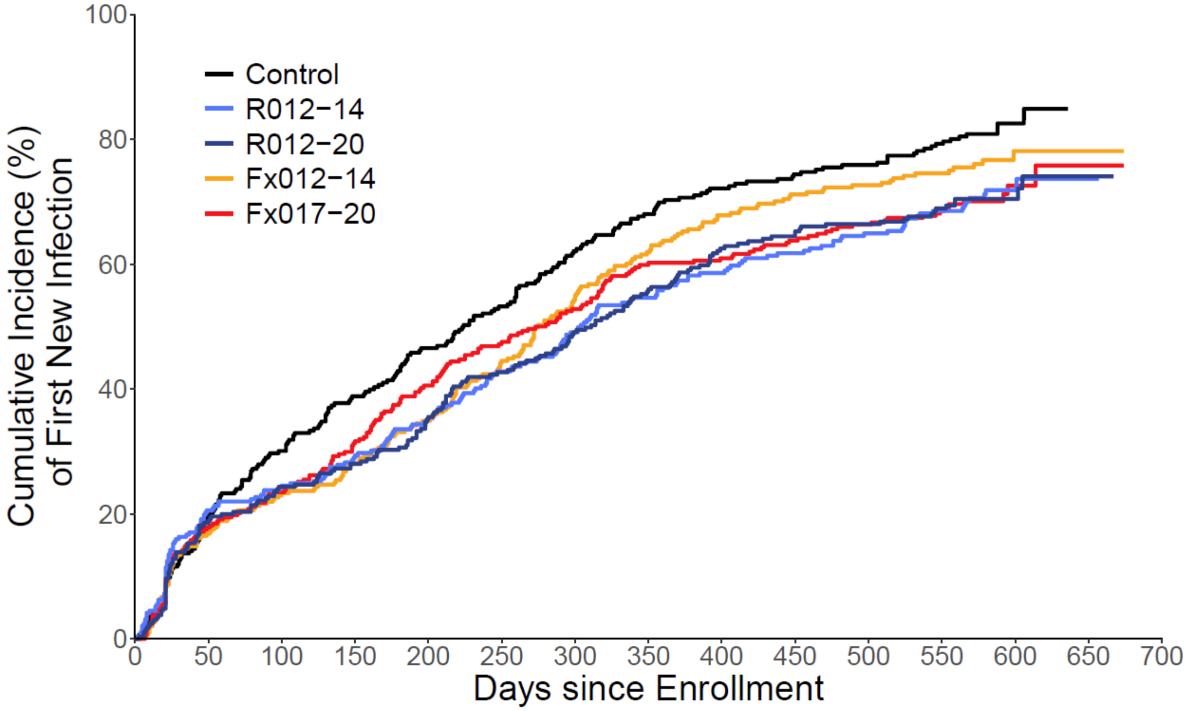

**No. At Risk for First New Infection**

| | | | | | | | | | | | | | | | |
|---|---|---|---|---|---|---|---|---|---|---|---|---|---|---|---|
| Control | 293 | 225 | 195 | 166 | 145 | 126 | 102 | 85 | 74 | 68 | 64 | 54 | 12 | 0 | 0 |
| R012−14 | 298 | 224 | 207 | 188 | 168 | 147 | 129 | 114 | 104 | 96 | 88 | 78 | 15 | 1 | 0 |
| R012−20 | 294 | 227 | 207 | 192 | 172 | 152 | 134 | 115 | 97 | 90 | 84 | 69 | 18 | 2 | 0 |
| Fx012−14 | 304 | 244 | 223 | 205 | 183 | 156 | 125 | 103 | 86 | 77 | 73 | 56 | 14 | 2 | 0 |
| Fx017−20 | 311 | 246 | 226 | 201 | 172 | 150 | 134 | 113 | 110 | 100 | 93 | 84 | 17 | 1 | 0 |

**Cumulative No. of First New Infections**

| | | | | | | | | | | | | | | | |
|---|---|---|---|---|---|---|---|---|---|---|---|---|---|---|---|
| Control | 0 | 56 | 85 | 109 | 130 | 148 | 172 | 188 | 199 | 205 | 209 | 218 | 224 | 225 | 225 |
| R012−14 | 0 | 59 | 70 | 83 | 99 | 118 | 134 | 148 | 158 | 166 | 174 | 183 | 190 | 191 | 191 |
| R012−20 | 0 | 54 | 69 | 79 | 99 | 118 | 135 | 153 | 170 | 177 | 180 | 186 | 189 | 191 | 191 |
| Fx012−14 | 0 | 50 | 69 | 83 | 102 | 129 | 158 | 177 | 193 | 202 | 206 | 211 | 216 | 216 | 216 |
| Fx017−20 | 0 | 54 | 71 | 95 | 121 | 141 | 156 | 177 | 179 | 187 | 194 | 200 | 206 | 207 | 207 |

**B**

| Treatment Comparison | No. of First New Infections (Incidence per PYR) | VE (%) (95% CI) | 2−Sided P−values: Unadj. | FWER−adj. | FDR−adj. |
|---|---|---|---|---|---|
| R012−20 vs. Control | 191 (0.82) vs. 225 (1.13) | 29.7 (14.7, 42.1) | < 0.001 | < 0.001 | < 0.001 |
| R012−14 vs. Control | 191 (0.82) vs. 225 (1.13) | 31.2 (16.4, 43.3) | < 0.001 | < 0.001 | < 0.001 |
| Fx012−14 vs. Control | 216 (0.93) vs. 225 (1.13) | 24.6 (9.0, 37.6) | 0.0033 | −− | −− |
| Fx017−20 vs. Control | 207 (0.85) vs. 225 (1.13) | 28.8 (13.9, 41.1) | < 0.001 | −− | −− |

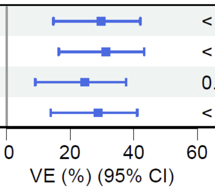

**Figure S5. Cumulative incidence (A) and vaccine efficacy (VE) (B) against the first new genotypic infection between enrollment and month 20 in the ES (Exposed Set).**
VE: vaccine efficacy; PYR: person-years at risk; Unadj: unadjusted; FWER: family-wise error rate; FDR-adj: false discovery rate adjusted; CI: confidence interval; No: number

**A**

| Treatment Comparison | No. of First New Infections (Incidence per PYR) | HR (95% CI) | 2−Sided P−values Unadj. | FWER−adj. | FDR−adj. |
|---|---|---|---|---|---|
| Fx012−14 vs. R012−20 plus R012−14 | 160 (0.94) vs. 282 (0.87) | 1.077 (0.884, 1.311) | 0.46 | 0.93 | 0.86 |
| Fx017−20 vs. R012−20 plus R012−14 | 145 (0.95)/174 (1.51)** vs. 282 (0.87)/176 (1.28)** | 1.024 (0.805, 1.299) | 0.86 | 0.93 | 0.86 |
| Fx017−20 vs. Fx012−14 | 145 (0.95)/174 (1.51)** vs. 160 (0.94)/176 (1.28)** | 0.926 (0.709, 1.197) | 0.54 | −− | −− |

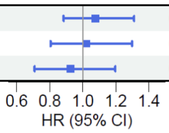

**B**

| Treatment Comparison | No. of First New Infections (Incidence per PYR) | HR (95% CI) | 2−Sided P−values Unadj. | FWER−adj. | FDR−adj. |
|---|---|---|---|---|---|
| R012−14 vs. R012−20 | 191 (0.82) vs. 191 (0.82) | 0.969 (0.793, 1.185) | 0.76 | 1 | 0.76 |
| Fx012−14 vs. R012−20 | 216 (0.93) vs. 191 (0.82) | 1.065 (0.876, 1.294) | 0.53 | 1 | 0.76 |
| Fx017−20 vs. R012−20 | 207 (0.85) vs. 191 (0.82) | 0.960 (0.788, 1.170) | 0.69 | 1 | 0.76 |
| Fx012−14 vs. R012−14 | 216 (0.93) vs. 191 (0.82) | 1.103 (0.908, 1.341) | 0.32 | 0.97 | 0.65 |
| Fx017−20 vs. R012−14 | 207 (0.85) vs. 191 (0.82) | 1.007 (0.826, 1.226) | 0.95 | 0.97 | 0.95 |
| Fx017−20 vs. Fx012−14 | 207 (0.85) vs. 216 (0.93) | 0.926 (0.765, 1.122) | 0.43 | 0.97 | 0.65 |

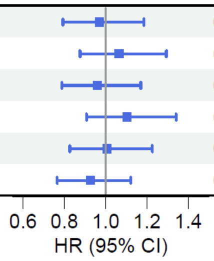

**Figure S6. Hazard ratio (HR) of the first new genotypic infection comparing RTS,S regimens head-to-head in the Per Protocol (PP) (A) and Exposed Set (ES) (B) cohorts.** VE: vaccine efficacy; PYR: person-years at risk; Unadj: unadjusted; FWER: family-wise error rate; FDR-adj: false discovery rate adjusted; CI: confidence interval; No: number

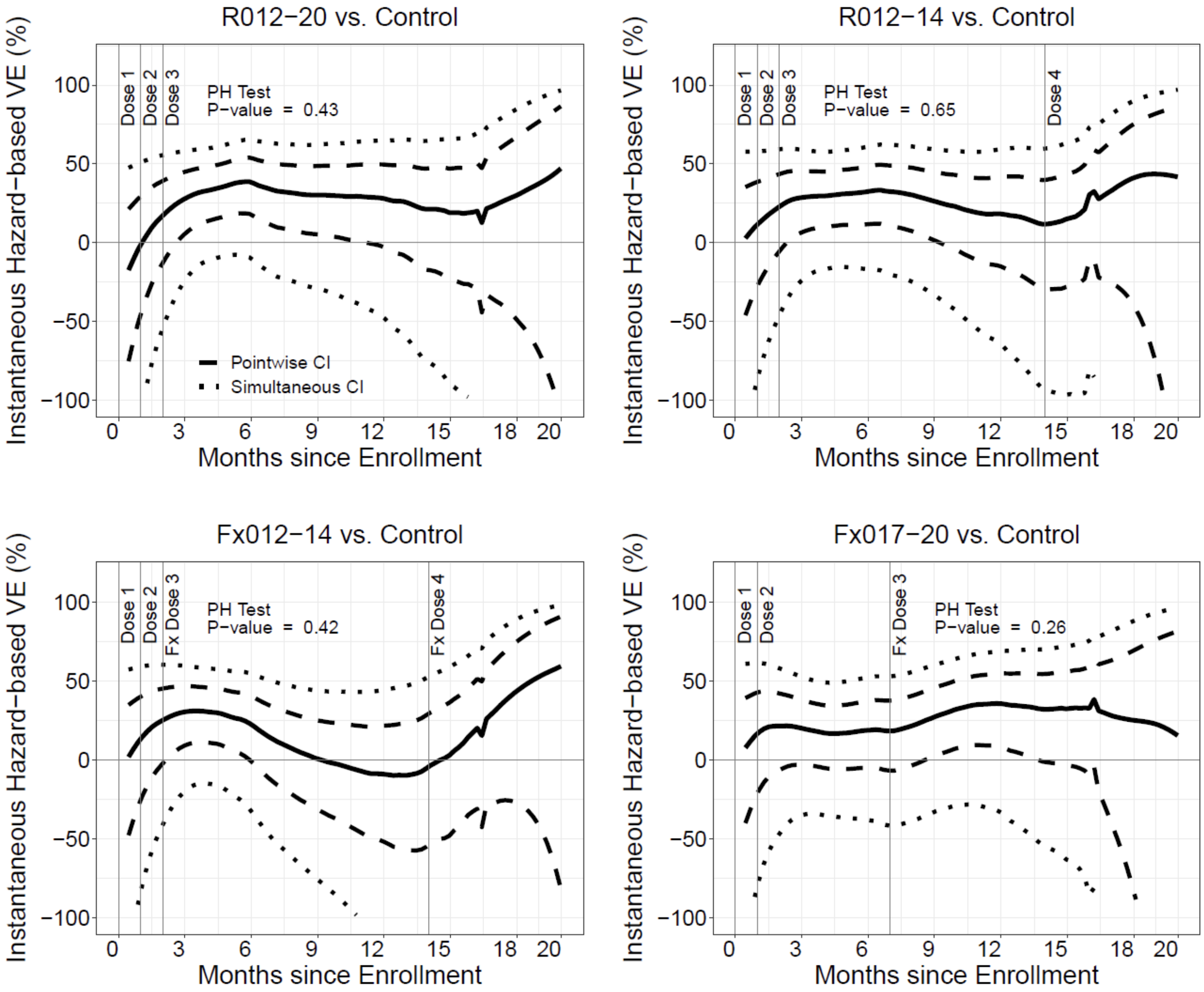

**Figure S7. Instantaneous vaccine efficacy against the first new genotypic infection over time since enrollment/first vaccination in the Exposed Set.**

Shown are 95% pointwise confidence intervals (dashed) and the 95% simultaneous confidence band (dotted). Optimal bandwidth was calculated using a bootstrap procedure. A p-value is reported from the test of whether instantaneous vaccine efficacy (VE) varies over time. CI: Confidence interval

**A**

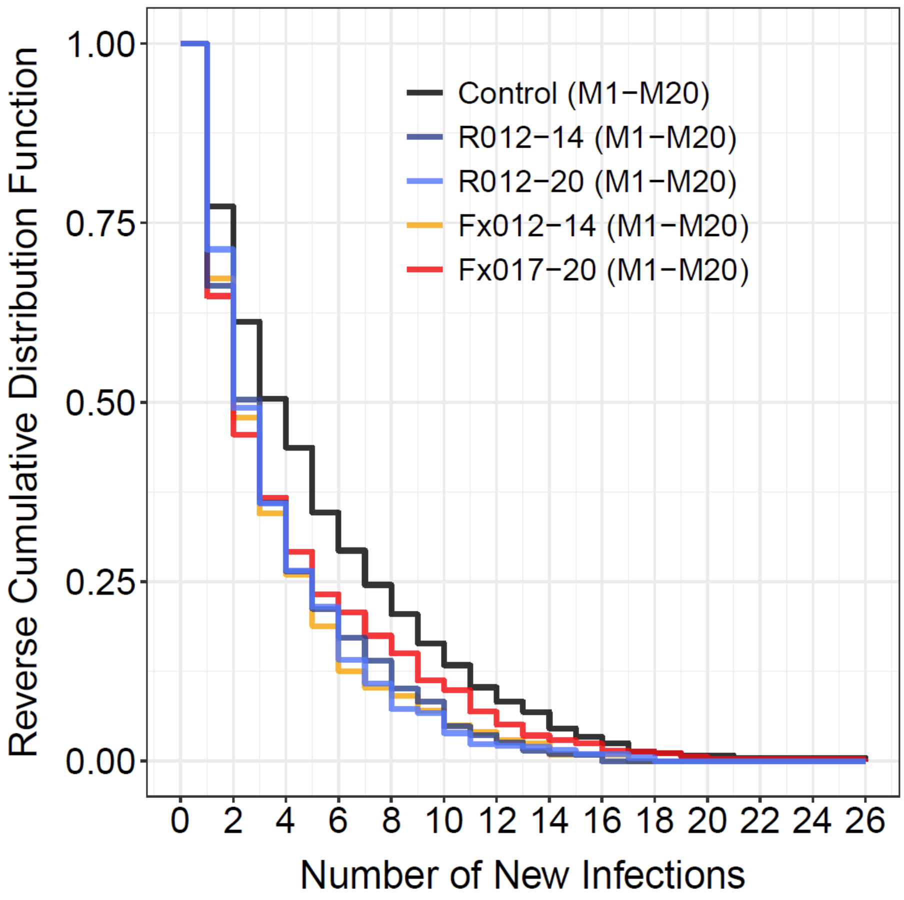

**B**

| Treatment Comparison | Mean No. of New Infections | Mean Diff. (95% CI) | 2–Sided P–values Unadj. | FWER–adj. | FDR–adj. |
|---|---|---|---|---|---|
| R012−20 vs. Control | 2.64 vs. 4.14 | −1.50 (−1.98, −1.02) | < 0.001 | < 0.001 | < 0.001 |
| R012−14 vs. Control | 2.59 vs. 4.14 | −1.54 (−2.03, −1.06) | < 0.001 | < 0.001 | < 0.001 |
| Fx012−14 vs. Control | 2.56 vs. 4.14 | −1.57 (−2.06, −1.08) | < 0.001 | −− | −− |
| Fx017−20 vs. Control | 3.03 vs. 4.14 | −1.11 (−1.64, −0.59) | < 0.001 | −− | −− |

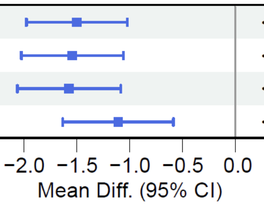

**Figure S8. Vaccine effects on the number of new genotypic infections between enrollment and month 20 in the ES.**

Reverse cumulative distribution of the number of new infections (A) and vaccine effects on the mean number of new infections vs. the control regimen (B) between the first dose and month 20. Unadj: unadjusted; FWER: family-wise error rate; FDR: false discovery rate; No: number; ES: Exposed Set; diff: difference; Adj: adjusted

| Treatment Comparison | Locus; Haplotype | No. of First New Infections (Incidence per PYR) | VE (%) (95% CI) | P-value | 2−sided Differential VE P−values: Unadj. | FWER−adj. | FDR−adj. |
|---|---|---|---|---|---|---|---|
| RTS,S Pooled vs. Control | CSP | | | | 0.76 | 1.00 | 0.95 |
| | 3D7 match | 16 (0.033) vs. 6 (0.046) | 32.8 (−56.8, 71.2) | 0.36 | | | |
| | 3D7 mismatch | 362 (0.73) vs. 151 (1.10) | 41.2 (29.4, 51.0) | <0.001 | | | |
| RTS,S Pooled vs. Control | Th2R | | | | 0.76 | 1.00 | 0.95 |
| | 3D7 match | 16 (0.033) vs. 6 (0.046) | 32.8 (−56.9, 71.2) | 0.36 | | | |
| | 3D7 mismatch | 362 (0.73) vs. 151 (1.10) | 41.2 (29.4, 51.0) | <0.001 | | | |
| RTS,S Pooled vs. Control | Th3R | | | | 0.50 | 1.00 | 0.95 |
| | 3D7 match | 23 (0.046) vs. 12 (0.09) | 52.2 (9.3, 74.8) | 0.024 | | | |
| | 3D7 mismatch | 355 (0.72) vs. 145 (1.05) | 40.0 (27.9, 50.1) | <0.001 | | | |
| RTS,S Pooled vs. Control | DV10 | | | | 0.96 | 1.00 | 0.96 |
| | 3D7 match | 20 (0.04) vs. 9 (0.064) | 41.8 (−21.6, 72.2) | 0.15 | | | |
| | 3D7 mismatch | 358 (0.72) vs. 148 (1.08) | 40.8 (29.0, 50.7) | <0.001 | | | |
| RTS,S Pooled vs. Control | LD | | | | 0.58 | 1.00 | 0.95 |
| | 3D7 match | 340 (0.69) vs. 143 (1.04) | 41.9 (29.8, 51.9) | <0.001 | | | |
| | 3D7 mismatch | 38 (0.076) vs. 14 (0.10) | 30.9 (−23.4, 61.3) | 0.21 | | | |
| R012−14 plus R012−20 vs. Control | Th3R | | | | 0.47 | 0.93 | 0.70 |
| | 3D7 match | 14 (0.045) vs. 12 (0.09) | 55.9 (9.3, 78.5) | 0.026 | | | |
| | 3D7 mismatch | 228 (0.70) vs. 145 (1.05) | 41.8 (29.1, 52.3) | <0.001 | | | |
| R012−14 plus R012−20 vs. Control | DV10 | | | | 0.88 | 0.93 | 0.88 |
| | 3D7 match | 13 (0.04) vs. 9 (0.064) | 39.2 (−37.4, 73.1) | 0.23 | | | |
| | 3D7 mismatch | 229 (0.70) vs. 148 (1.08) | 43.0 (30.6, 53.2) | <0.001 | | | |
| R012−14 plus R012−20 vs. Control | LD | | | | 0.26 | 0.77 | 0.70 |
| | 3D7 match | 213 (0.65) vs. 143 (1.04) | 44.9 (32.5, 55.1) | <0.001 | | | |
| | 3D7 mismatch | 29 (0.09) vs. 14 (0.10) | 20.5 (−44.3, 56.3) | 0.45 | | | |
| Fx012−14 vs. Control | Th3R | | | | 0.79 | 0.79 | 0.79 |
| | 3D7 match | 8 (0.049) vs. 12 (0.091) | 42.9 (−26.1, 74.1) | 0.17 | | | |
| | 3D7 mismatch | 128 (0.75) vs. 145 (1.05) | 36.1 (20.0, 49.0) | <0.001 | | | |
| Fx012−14 vs. Control | LD | | | | 0.39 | 0.78 | 0.78 |
| | 3D7 match | 127 (0.75) vs. 143 (1.04) | 34.9 (18.3, 48.1) | <0.001 | | | |
| | 3D7 mismatch | 9 (0.05) vs. 14 (0.10) | 54.9 (−1.6, 80.0) | 0.055 | | | |
| Fx017−20 vs. Control | Th3R | | | | 0.34 | 0.68 | 0.68 |
| | 3D7 match | 6 (0.039) vs. 13 (0.11) | 62.2 (4.9, 84.9) | 0.039 | | | |
| | 3D7 mismatch | 123 (0.81) vs. 145 (1.26) | 39.7 (24.2, 52.0) | <0.001 | | | |
| Fx017−20 vs. Control | LD | | | | 0.90 | 0.90 | 0.90 |
| | 3D7 match | 116 (0.76) vs. 141 (1.22) | 41.2 (26.0, 53.3) | <0.001 | | | |
| | 3D7 mismatch | 13 (0.086) vs. 17 (0.15) | 43.9 (−9.6, 71.2) | 0.091 | | | |

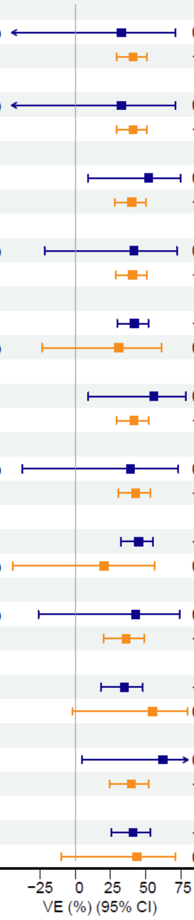

**Figure S9. Vaccine efficacy against the first new genotypic infection with a match vs. mismatch to the 3D7 amino acid sequence in screened-in CS C-terminus haplotypic regions in the Per Protocol cohort.**

"RTS,S Pooled" designates the R012-14, R012-20, and Fx012-14 regimens combined. Unadj: unadjusted; FWER: family-wise error rate; FDR: false discovery rate; VE: vaccine efficacy; adj: adjusted; CI: confidence interval; No: number

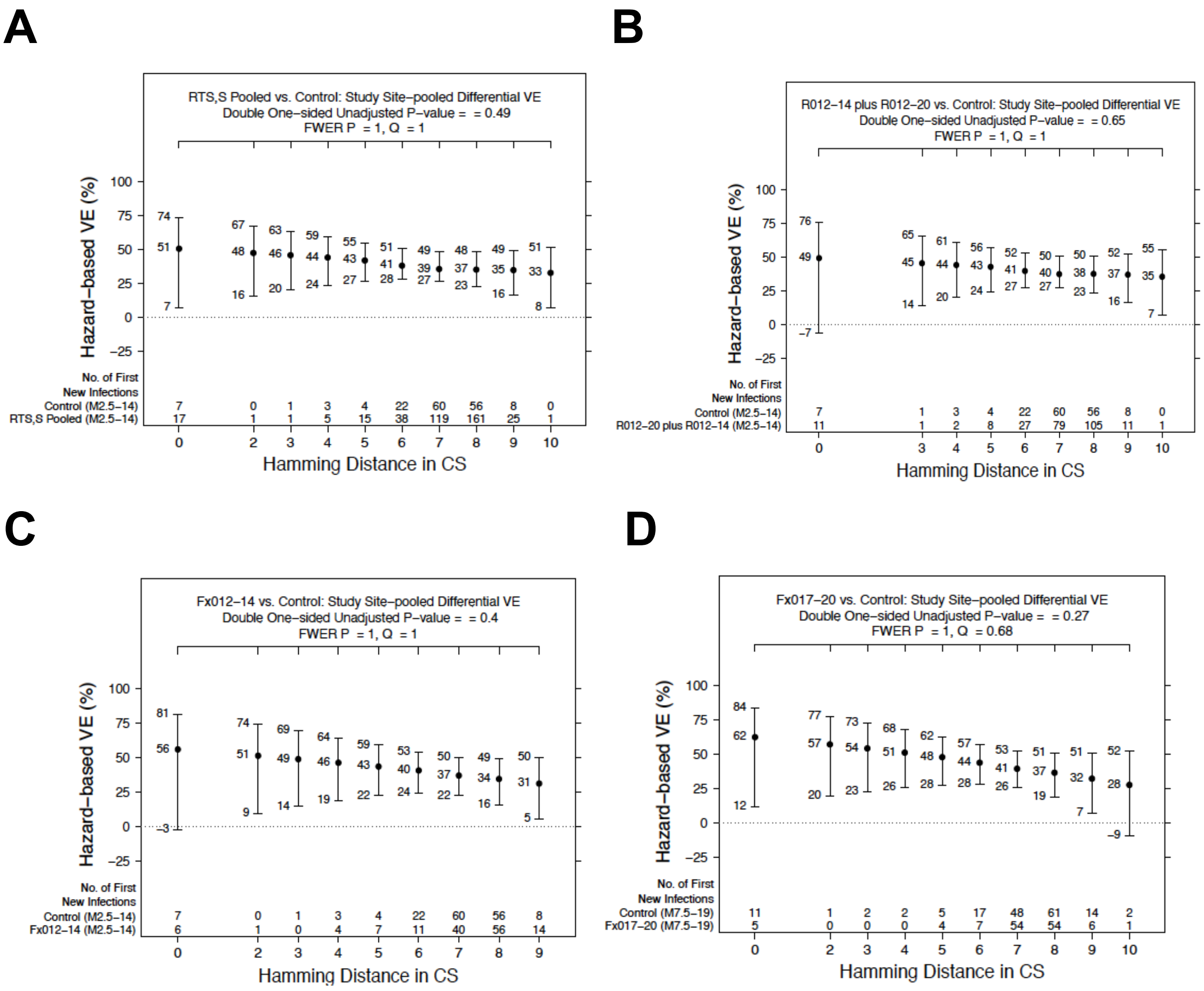

**Figure S10. Vaccine efficacy against the first new genotypic infection by Hamming distance to the 3D7 vaccine strain in the CS C-terminus for the Per Protocol cohort.** Shown are comparisons of pooled R012-14, R012-20, and Fx012-14 regimens (A), pooled R012-14 and R012-20 regimens (B), Fx012-14 (C), and Fx017-20 (D), each vs. the control regimen. VE: vaccine efficacy; FWER: family-wise error rate; FDR-adj: false discovery rate adjusted; No: number

| Amino Acid Position | Locus; Haplotype | No. of First New Infections (Incidence per PYR) | VE (%) (95% CI) | P−value | 2−sided Differential VE P−values: Unadj. | FWER−adj. | FDR−adj. |
|---|---|---|---|---|---|---|---|
| 298 | DV10 | | | | 0.43 | 1 | 0.72 |
| | 3D7 match | 327 (0.66) vs. 131 (0.96) | 39.1 (26.2, 49.8) | 4.1e−07 | | | |
| | 3D7 mismatch | 51 (0.10) vs. 26 (0.19) | 49.6 (21.8, 67.5) | 0.0023 | | | |
| 301 | DV10 | | | | 0.95 | 1 | 0.95 |
| | 3D7 match | 21 (0.042) vs. 9 (0.064) | 39.6 (−25.6, 70.9) | 0.18 | | | |
| | 3D7 mismatch | 357 (0.72) vs. 148 (1.08) | 40.9 (29.1, 50.8) | 1.6e−08 | | | |
| 314 | Th2R | | | | 0.87 | 1 | 0.93 |
| | 3D7 match | 215 (0.43) vs. 90 (0.66) | 40.2 (24.5, 52.6) | 1.5e−05 | | | |
| | 3D7 mismatch | 163 (0.33) vs. 67 (0.49) | 41.8 (24.1, 55.4) | 6.5e−05 | | | |
| 317 | Th2R | | | | 0.74 | 1 | 0.85 |
| | 3D7 match | 37 (0.075) vs. 14 (0.10) | 35.1 (−14.9, 63.3) | 0.14 | | | |
| | 3D7 mismatch | 341 (0.69) vs. 143 (1.04) | 41.3 (29.4, 51.3) | 1.6e−08 | | | |
| 318 | Th2R | | | | 0.10 | 1 | 0.39 |
| | 3D7 match | 76 (0.15) vs. 23 (0.17) | 18.7 (−25.4, 47.3) | 0.35 | | | |
| | 3D7 mismatch | 302 (0.61) vs. 134 (0.97) | 44.6 (32.9, 54.3) | 1.7e−09 | | | |
| 321 | Th2R | | | | 0.38 | 1 | 0.72 |
| | 3D7 match | 49 (0.098) vs. 24 (0.17) | 50.6 (22.7, 68.4) | 0.002 | | | |
| | 3D7 mismatch | 329 (0.67) vs. 133 (0.97) | 39.1 (26.4, 49.6) | 2.7e−07 | | | |
| 322 | Th2R | | | | 0.092 | 1 | 0.39 |
| | 3D7 match | 146 (0.29) vs. 49 (0.36) | 27.3 (0.7, 46.8) | 0.045 | | | |
| | 3D7 mismatch | 232 (0.47) vs. 108 (0.79) | 46.9 (34.4, 57.0) | 4.1e−09 | | | |
| 324 | Th2R | | | | 0.18 | 1 | 0.52 |
| | 3D7 match | 234 (0.47) vs. 106 (0.77) | 45.8 (32.7, 56.3) | 2.8e−08 | | | |
| | 3D7 mismatch | 144 (0.29) vs. 51 (0.37) | 30.4 (6.0, 48.5) | 0.018 | | | |
| 327 | Th2R | | | | 0.062 | 0.94 | 0.39 |
| | 3D7 match | 309 (0.62) vs. 138 (1.01) | 44.8 (33.3, 54.3) | 7.3e−10 | | | |
| | 3D7 mismatch | 69 (0.14) vs. 19 (0.14) | 11.3 (−42.7, 44.8) | 0.62 | | | |
| 352 | Th3R | | | | 0.65 | 1 | 0.81 |
| | 3D7 match | 312 (0.63) vs. 133 (0.97) | 41.8 (29.4, 52.0) | 3.8e−08 | | | |
| | 3D7 mismatch | 66 (0.13) vs. 24 (0.17) | 35.1 (−0.5, 58.1) | 0.053 | | | |
| 354 | Th3R | | | | 0.65 | 1 | 0.81 |
| | 3D7 match | 359 (0.72) vs. 150 (1.10) | 41.4 (29.7, 51.1) | 7.7e−09 | | | |
| | 3D7 mismatch | 19 (0.038) vs. 7 (0.048) | 28.9 (−59.8, 68.4) | 0.41 | | | |
| 356 | Th3R | | | | 0.63 | 1 | 0.81 |
| | 3D7 match | 341 (0.69) vs. 143 (1.04) | 41.8 (29.7, 51.8) | 1.9e−08 | | | |
| | 3D7 mismatch | 37 (0.075) vs. 14 (0.10) | 32.2 (−21.4, 62.2) | 0.19 | | | |
| 357 | Th3R | | | | 0.38 | 1 | 0.72 |
| | 3D7 match | 134 (0.27) vs. 52 (0.38) | 34.3 (11.1, 51.4) | 0.0064 | | | |
| | 3D7 mismatch | 244 (0.49) vs. 105 (0.77) | 44.1 (30.6, 55.0) | 1.5e−07 | | | |
| 359 | Th3R | | | | 0.21 | 1 | 0.52 |
| | 3D7 match | 319 (0.64) vs. 136 (0.99) | 43.5 (31.5, 53.4) | 6.6e−09 | | | |
| | 3D7 mismatch | 59 (0.12) vs. 21 (0.15) | 22.3 (−23.5, 51.1) | 0.29 | | | |
| 361 | Th3R | | | | 0.089 | 1 | 0.39 |
| | 3D7 match | 168 (0.34) vs. 80 (0.58) | 49.4 (35.1, 60.6) | 8.4e−08 | | | |
| | 3D7 mismatch | 210 (0.42) vs. 77 (0.56) | 31.8 (12.8, 46.7) | 0.0023 | | | |

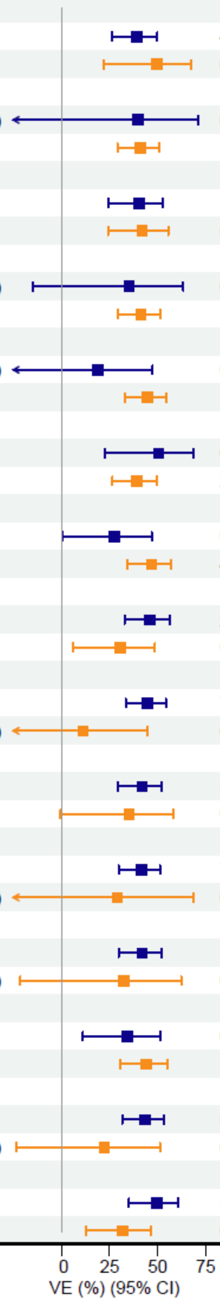

**Figure S11. Vaccine efficacy (VE) of the pooled R012-14, R012-20, and Fx012-14 regimens vs. the control regimen against the first new genotypic infection with a 3D7 residue match vs. mismatch at screened-in CS C-terminus amino acid positions in the Per Protocol Cohort.**

Unadj: unadjusted; FWER: family-wise error rate; FDR: false discovery rate; CI: confidence interval; No: number; adj: adjusted; PYR: person-years at risk

| Amino Acid Position | Locus; Haplotype | No. of First New Infections (Incidence per PYR) | VE (%) (95% CI) | P-value | 2-sided Differential VE P-values: Unadj. | FWER-adj. | FDR-adj. |
|---|---|---|---|---|---|---|---|
| 298 | DV10 | | | | 0.47 | 1 | 0.79 |
| | 3D7 match | 210 (0.65) vs. 131 (0.96) | 41.1 (27.6, 52.0) | 4.7e−07 | | | |
| | 3D7 mismatch | 32 (0.099) vs. 26 (0.19) | 51.4 (20.8, 70.1) | 0.0037 | | | |
| 301 | DV10 | | | | 0.75 | 1 | 0.92 |
| | 3D7 match | 14 (0.043) vs. 9 (0.064) | 35.0 (−45.3, 70.9) | 0.29 | | | |
| | 3D7 mismatch | 228 (0.70) vs. 148 (1.08) | 43.2 (30.9, 53.4) | 1.8e−08 | | | |
| 314 | Th2R | | | | 0.82 | 1 | 0.92 |
| | 3D7 match | 139 (0.43) vs. 90 (0.66) | 41.8 (25.2, 54.6) | 2.3e−05 | | | |
| | 3D7 mismatch | 103 (0.32) vs. 67 (0.49) | 44.2 (25.6, 58.2) | 7.4e−05 | | | |
| 317 | Th2R | | | | 0.97 | 1 | 0.97 |
| | 3D7 match | 21 (0.064) vs. 14 (0.10) | 42.0 (−10.6, 69.6) | 0.098 | | | |
| | 3D7 mismatch | 221 (0.68) vs. 143 (1.04) | 42.8 (30.1, 53.1) | 4.4e−08 | | | |
| 318 | Th2R | | | | 0.067 | 1 | 0.77 |
| | 3D7 match | 51 (0.16) vs. 23 (0.17) | 16.1 (−32.4, 46.8) | 0.45 | | | |
| | 3D7 mismatch | 191 (0.59) vs. 134 (0.97) | 47.2 (34.9, 57.2) | 2.5e−09 | | | |
| 321 | Th2R | | | | 0.41 | 1 | 0.77 |
| | 3D7 match | 31 (0.096) vs. 24 (0.17) | 52.6 (22.2, 71.2) | 0.0031 | | | |
| | 3D7 mismatch | 211 (0.65) vs. 133 (0.97) | 40.9 (27.6, 51.8) | 4.2e−07 | | | |
| 322 | Th2R | | | | 0.38 | 1 | 0.77 |
| | 3D7 match | 88 (0.27) vs. 49 (0.36) | 35.4 (9.6, 53.9) | 0.011 | | | |
| | 3D7 mismatch | 154 (0.47) vs. 108 (0.79) | 46.0 (32.3, 57.0) | 1.1e−07 | | | |
| 324 | Th2R | | | | 0.56 | 1 | 0.79 |
| | 3D7 match | 156 (0.48) vs. 106 (0.77) | 45.0 (30.7, 56.3) | 3.8e−07 | | | |
| | 3D7 mismatch | 86 (0.27) vs. 51 (0.37) | 38.1 (14.0, 55.5) | 0.0043 | | | |
| 327 | Th2R | | | | 0.19 | 1 | 0.77 |
| | 3D7 match | 201 (0.62) vs. 138 (1.01) | 45.6 (33.3, 55.6) | 4.8e−09 | | | |
| | 3D7 mismatch | 41 (0.13) vs. 19 (0.14) | 22.0 (−29.9, 53.1) | 0.34 | | | |
| 352 | Th3R | | | | 0.86 | 1 | 0.92 |
| | 3D7 match | 204 (0.63) vs. 133 (0.97) | 43.1 (30.0, 53.8) | 1.1e−07 | | | |
| | 3D7 mismatch | 38 (0.12) vs. 24 (0.17) | 40.4 (3.5, 63.2) | 0.035 | | | |
| 354 | Th3R | | | | 0.24 | 1 | 0.77 |
| | 3D7 match | 226 (0.70) vs. 150 (1.10) | 44.2 (32.2, 54.1) | 4.9e−09 | | | |
| | 3D7 mismatch | 16 (0.048) vs. 7 (0.048) | 7.7 (−108.8, 59.2) | 0.85 | | | |
| 356 | Th3R | | | | 0.30 | 1 | 0.77 |
| | 3D7 match | 213 (0.66) vs. 143 (1.04) | 44.7 (32.3, 54.9) | 1.1e−08 | | | |
| | 3D7 mismatch | 29 (0.088) vs. 14 (0.10) | 22.8 (−40.5, 57.5) | 0.40 | | | |
| 357 | Th3R | | | | 0.40 | 1 | 0.77 |
| | 3D7 match | 88 (0.27) vs. 52 (0.38) | 36.1 (11.8, 53.6) | 0.0063 | | | |
| | 3D7 mismatch | 154 (0.47) vs. 105 (0.77) | 46.0 (31.6, 57.3) | 3.1e−07 | | | |
| 359 | Th3R | | | | 0.58 | 1 | 0.79 |
| | 3D7 match | 208 (0.64) vs. 136 (0.99) | 43.9 (31.0, 54.3) | 4e−08 | | | |
| | 3D7 mismatch | 34 (0.10) vs. 21 (0.15) | 34.6 (−8.2, 60.5) | 0.099 | | | |
| 361 | Th3R | | | | 0.24 | 1 | 0.77 |
| | 3D7 match | 109 (0.34) vs. 80 (0.58) | 49.1 (33.3, 61.2) | 9.3e−07 | | | |
| | 3D7 mismatch | 133 (0.41) vs. 77 (0.56) | 36.3 (16.9, 51.2) | 0.00089 | | | |

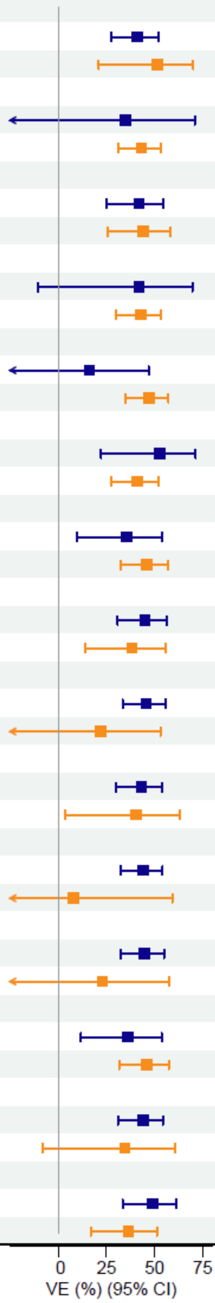

**Figure S12. Vaccine efficacy (VE) of the pooled R012-14 and R012-20 regimens vs. the control regimen against the first new genotypic infection with a 3D7 residue match vs. mismatch at screened-in CS C-terminus amino acid positions in the Per Protocol cohort.** PYR: person-years at risk; Unadj: unadjusted; FWER adj: family-wise error rate adjusted; FDR-adj: false discovery rate adjusted; CI: confidence interval; No: number

| Amino Acid Position | Locus; Haplotype | No. of First New Infections (Incidence per PYR) | VE (%) (95% CI) | P-value | 2−sided Differential VE P−values: Unadj. | FWER−adj. | FDR−adj. |
|---|---|---|---|---|---|---|---|
| 298 | DV10 | | | | 0.46 | 1 | 0.67 |
| | 3D7 match | 117 (0.69) vs. 131 (0.96) | 34.3 (16.8, 48.1) | 0.00048 | | | |
| | 3D7 mismatch | 19 (0.11) vs. 26 (0.19) | 47.7 (8.2, 70.2) | 0.024 | | | |
| 314 | Th2R | | | | 0.63 | 1 | 0.69 |
| | 3D7 match | 76 (0.45) vs. 90 (0.66) | 39.4 (18.9, 54.7) | 0.00075 | | | |
| | 3D7 mismatch | 60 (0.35) vs. 67 (0.49) | 32.7 (6.8, 51.4) | 0.017 | | | |
| 317 | Th2R | | | | 0.51 | 1 | 0.67 |
| | 3D7 match | 16 (0.096) vs. 14 (0.10) | 22.6 (−46.6, 59.1) | 0.43 | | | |
| | 3D7 mismatch | 120 (0.70) vs. 143 (1.04) | 38.0 (22.1, 50.6) | 3.9e−05 | | | |
| 318 | Th2R | | | | 0.51 | 1 | 0.67 |
| | 3D7 match | 25 (0.15) vs. 23 (0.17) | 25.6 (−26.2, 56.1) | 0.27 | | | |
| | 3D7 mismatch | 111 (0.65) vs. 134 (0.97) | 38.5 (22.3, 51.4) | 4.9e−05 | | | |
| 321 | Th2R | | | | 0.61 | 1 | 0.69 |
| | 3D7 match | 17 (0.10) vs. 24 (0.17) | 44.3 (5.4, 67.2) | 0.03 | | | |
| | 3D7 mismatch | 119 (0.70) vs. 133 (0.97) | 35.5 (18.7, 48.8) | 0.00021 | | | |
| 322 | Th2R | | | | 0.015 | 0.19 | 0.13 |
| | 3D7 match | 58 (0.34) vs. 49 (0.36) | 10.0 (−29.5, 37.4) | 0.57 | | | |
| | 3D7 mismatch | 78 (0.46) vs. 108 (0.79) | 48.2 (32.2, 60.5) | 1.7e−06 | | | |
| 324 | Th2R | | | | 0.021 | 0.25 | 0.13 |
| | 3D7 match | 78 (0.46) vs. 106 (0.77) | 48.0 (31.3, 60.7) | 4.1e−06 | | | |
| | 3D7 mismatch | 58 (0.34) vs. 51 (0.37) | 12.2 (−24.7, 38.1) | 0.47 | | | |
| 327 | Th2R | | | | 0.04 | 0.44 | 0.17 |
| | 3D7 match | 109 (0.64) vs. 138 (1.01) | 42.4 (27.0, 54.5) | 4.7e−06 | | | |
| | 3D7 mismatch | 27 (0.16) vs. 19 (0.14) | −6.3 (−83.3, 38.3) | 0.83 | | | |
| 352 | Th3R | | | | 0.47 | 1 | 0.67 |
| | 3D7 match | 108 (0.64) vs. 133 (0.97) | 38.9 (22.4, 51.8) | 5.3e−05 | | | |
| | 3D7 mismatch | 28 (0.16) vs. 24 (0.17) | 24.5 (−27.2, 55.2) | 0.29 | | | |
| 356 | Th3R | | | | 0.39 | 1 | 0.67 |
| | 3D7 match | 127 (0.75) vs. 143 (1.04) | 34.9 (18.3, 48.1) | 2e−04 | | | |
| | 3D7 mismatch | 9 (0.05) vs. 14 (0.10) | 54.9 (−1.8, 80.0) | 0.055 | | | |
| 357 | Th3R | | | | 0.72 | 1 | 0.72 |
| | 3D7 match | 46 (0.27) vs. 52 (0.38) | 33.0 (3.1, 53.7) | 0.033 | | | |
| | 3D7 mismatch | 90 (0.53) vs. 105 (0.77) | 38.3 (19.8, 52.6) | 0.00031 | | | |
| 359 | Th3R | | | | 0.086 | 0.86 | 0.28 |
| | 3D7 match | 111 (0.65) vs. 136 (0.99) | 41.6 (25.9, 53.9) | 9.2e−06 | | | |
| | 3D7 mismatch | 25 (0.15) vs. 21 (0.15) | 2.7 (−66.3, 43.0) | 0.92 | | | |
| 361 | Th3R | | | | 0.20 | 1 | 0.53 |
| | 3D7 match | 59 (0.35) vs. 80 (0.58) | 45.1 (25.2, 59.8) | 0.00015 | | | |
| | 3D7 mismatch | 77 (0.45) vs. 77 (0.56) | 27.5 (2.0, 46.4) | 0.036 | | | |

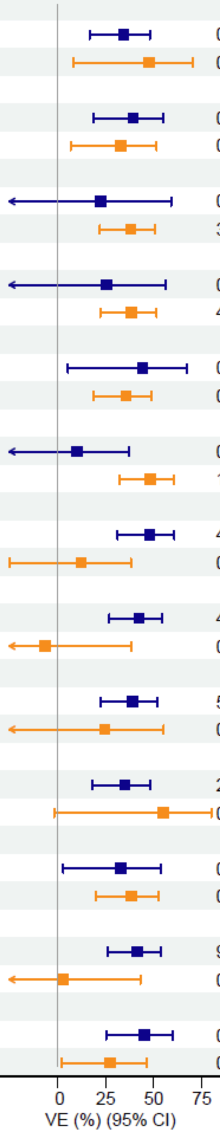

**Figure S13. Vaccine efficacy (VE) of the Fx012-14 regimen vs. the control regimen against the first new genotypic infection with a 3D7 residue match vs. mismatch at screened-in CS C-terminus amino acid positions in the Per Protocol cohort.**

PYR: person-years at risk; Unadj: unadjusted; FWER-adj: family-wise error rate adjusted; FDR-adj: false discovery rate adjusted; CI: confidence interval; No: number

| Amino Acid Position | Locus; Haplotype | No. of First New Infections (Incidence per PYR) | VE (%) (95% CI) | P-value | 2-sided Differential VE P-values Unadj. | FWER-adj. | FDR-adj. |
|---|---|---|---|---|---|---|---|
| 298 | DV10 | | | | 0.45 | 1 | 0.91 |
| | 3D7 match | 111 (0.73) vs. 132 (1.15) | 39.4 (23.4, 52.1) | 2.9e−05 | | | |
| | 3D7 mismatch | 18 (0.12) vs. 26 (0.22) | 51.3 (17.0, 71.4) | 0.0081 | | | |
| 314 | Th2R | | | | 0.67 | 1 | 0.91 |
| | 3D7 match | 72 (0.47) vs. 93 (0.80) | 43.6 (25.1, 57.6) | 7.7e−05 | | | |
| | 3D7 mismatch | 57 (0.37) vs. 65 (0.57) | 38.1 (13.7, 55.6) | 0.0047 | | | |
| 317 | Th2R | | | | 0.051 | 0.67 | 0.36 |
| | 3D7 match | 8 (0.055) vs. 22 (0.19) | 71.4 (38.8, 86.7) | 0.0013 | | | |
| | 3D7 mismatch | 121 (0.79) vs. 136 (1.18) | 36.8 (20.2, 49.9) | 0.00011 | | | |
| 318 | Th2R | | | | 0.22 | 1 | 0.77 |
| | 3D7 match | 20 (0.13) vs. 32 (0.28) | 56.2 (26.7, 73.8) | 0.0017 | | | |
| | 3D7 mismatch | 109 (0.72) vs. 126 (1.09) | 37.6 (20.7, 50.9) | 0.00012 | | | |
| 321 | Th2R | | | | 0.025 | 0.34 | 0.34 |
| | 3D7 match | 12 (0.082) vs. 29 (0.26) | 69.5 (43.5, 83.6) | 0.00016 | | | |
| | 3D7 mismatch | 117 (0.77) vs. 129 (1.11) | 35.2 (18.0, 48.8) | 0.00031 | | | |
| 322 | Th2R | | | | 0.38 | 1 | 0.91 |
| | 3D7 match | 35 (0.23) vs. 49 (0.43) | 49.4 (24.5, 66.0) | 0.00083 | | | |
| | 3D7 mismatch | 94 (0.62) vs. 109 (0.94) | 37.6 (19.2, 51.8) | 0.00035 | | | |
| 324 | Th2R | | | | 0.84 | 1 | 0.91 |
| | 3D7 match | 82 (0.54) vs. 101 (0.88) | 42.3 (24.3, 56.0) | 7.1e−05 | | | |
| | 3D7 mismatch | 47 (0.31) vs. 57 (0.49) | 39.6 (14.0, 57.6) | 0.0051 | | | |
| 327 | Th2R | | | | 0.77 | 1 | 0.91 |
| | 3D7 match | 114 (0.75) vs. 139 (1.20) | 40.7 (25.2, 53.0) | 1e−05 | | | |
| | 3D7 mismatch | 15 (0.099) vs. 19 (0.17) | 46.1 (1.3, 70.6) | 0.045 | | | |
| 352 | Th3R | | | | 0.84 | 1 | 0.91 |
| | 3D7 match | 103 (0.68) vs. 124 (1.07) | 40.6 (24.0, 53.5) | 3.2e−05 | | | |
| | 3D7 mismatch | 26 (0.17) vs. 34 (0.30) | 43.7 (8.2, 65.5) | 0.021 | | | |
| 354 | Th3R | | | | 0.20 | 1 | 0.77 |
| | 3D7 match | 119 (0.78) vs. 151 (1.30) | 43.4 (29.1, 54.8) | 7.5e−07 | | | |
| | 3D7 mismatch | 10 (0.067) vs. 7 (0.063) | −1.6 (−143.4, 57.6) | 0.97 | | | |
| 356 | Th3R | | | | 0.97 | 1 | 0.97 |
| | 3D7 match | 116 (0.76) vs. 142 (1.23) | 41.4 (26.3, 53.4) | 5.1e−06 | | | |
| | 3D7 mismatch | 13 (0.083) vs. 16 (0.14) | 42.2 (−15.3, 71.0) | 0.12 | | | |
| 357 | Th3R | | | | 0.56 | 1 | 0.91 |
| | 3D7 match | 41 (0.27) vs. 47 (0.40) | 35.4 (4.9, 56.1) | 0.027 | | | |
| | 3D7 mismatch | 88 (0.58) vs. 111 (0.96) | 43.8 (26.7, 56.8) | 2e−05 | | | |
| 359 | Th3R | | | | 0.76 | 1 | 0.91 |
| | 3D7 match | 117 (0.77) vs. 142 (1.23) | 40.9 (25.7, 52.9) | 6.4e−06 | | | |
| | 3D7 mismatch | 12 (0.078) vs. 16 (0.14) | 47.1 (−3.9, 73.0) | 0.064 | | | |
| 361 | Th3R | | | | 0.78 | 1 | 0.91 |
| | 3D7 match | 65 (0.43) vs. 82 (0.71) | 43.0 (22.8, 57.9) | 0.00029 | | | |
| | 3D7 mismatch | 64 (0.42) vs. 76 (0.66) | 39.5 (17.3, 55.7) | 0.0016 | | | |

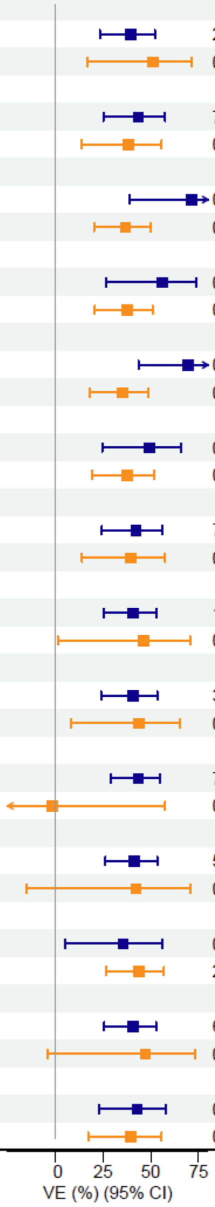

**Figure S14. Vaccine efficacy of the Fx017-20 regimen vs. the control regimen against the first new genotypic infection with a 3D7 residue match vs. mismatch at screened-in CS C-terminus amino acid positions in the Per Protocol cohort.**

PYR: person-years at risk; Unadj: unadjusted; FWER-adj: family-wise error rate adjusted; FDR-adj: false discovery rate adjusted; CI: confidence interval; No: number

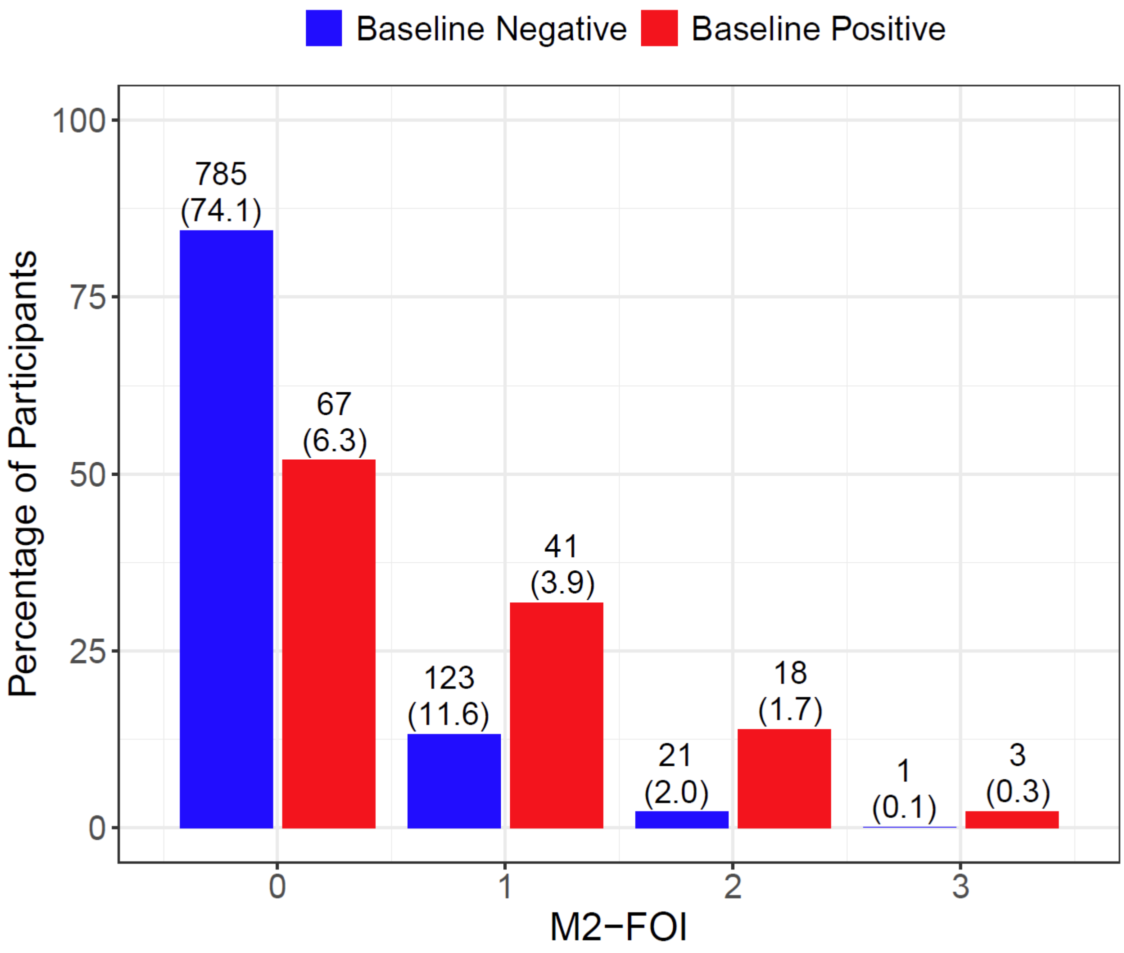

**Figure S15. Distribution of M2-FOI separately among PP baseline negative vs. baseline positive participants in the pooled control, R012-14, R012-20, and Fx012-14 groups. For percentages in parentheses, the denominator is all PP participants in the pooled control, R012-14, R012-20, and Fx012-14 groups. Spearman's correlation between baseline positivity and M2-FOI was 0.28 (P<0.001).**

M2-FOI: month 2 force of infection; PP: Per-Protocol

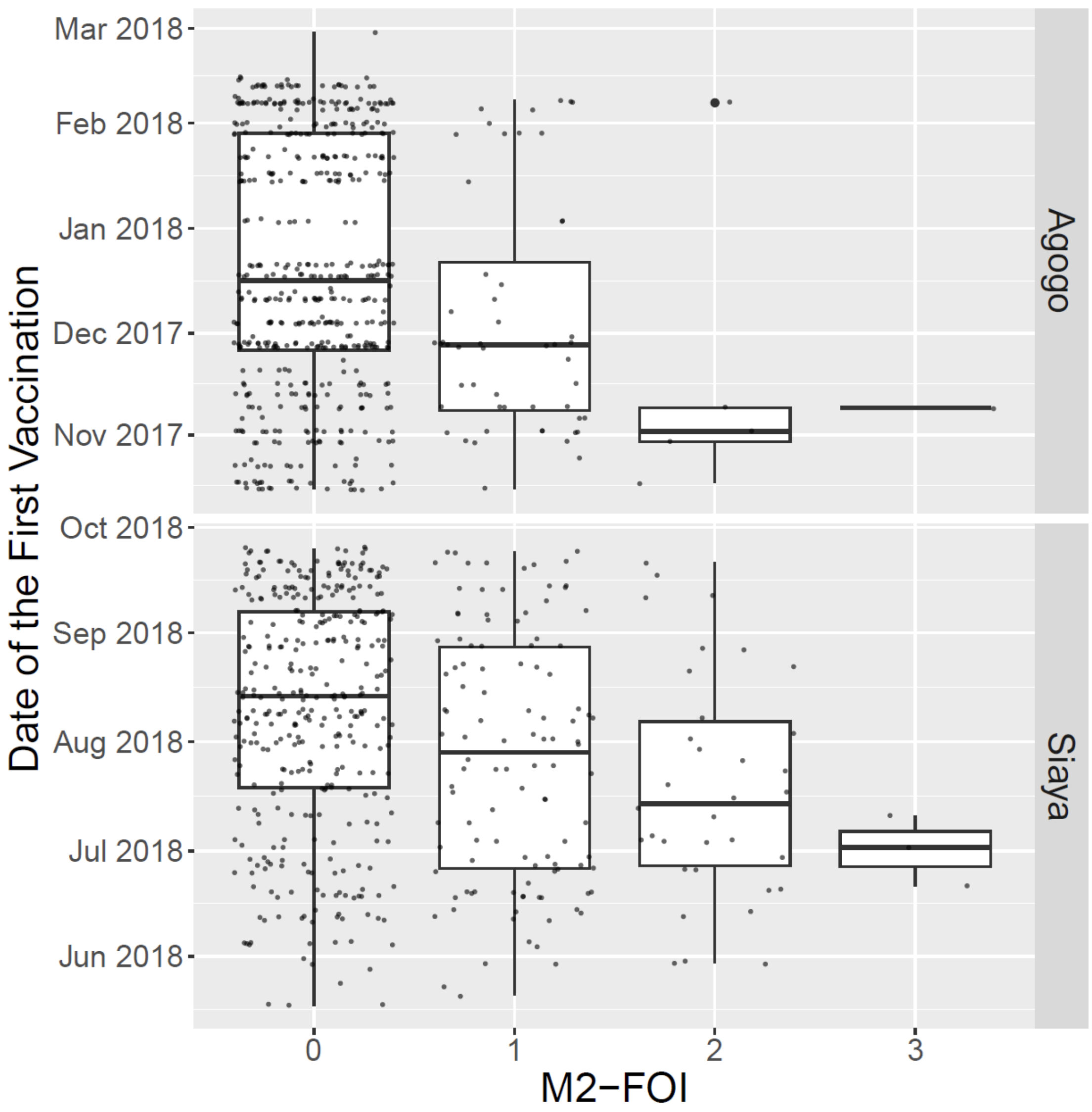

**Figure S16. Date of the first vaccination of PP participants in the pooled control, R012-14, R012-20, and Fx012-14 groups by M2-FOI and stratified by study site.**
M2-FOI: month 2 force of infection; PP: Per-Protocol

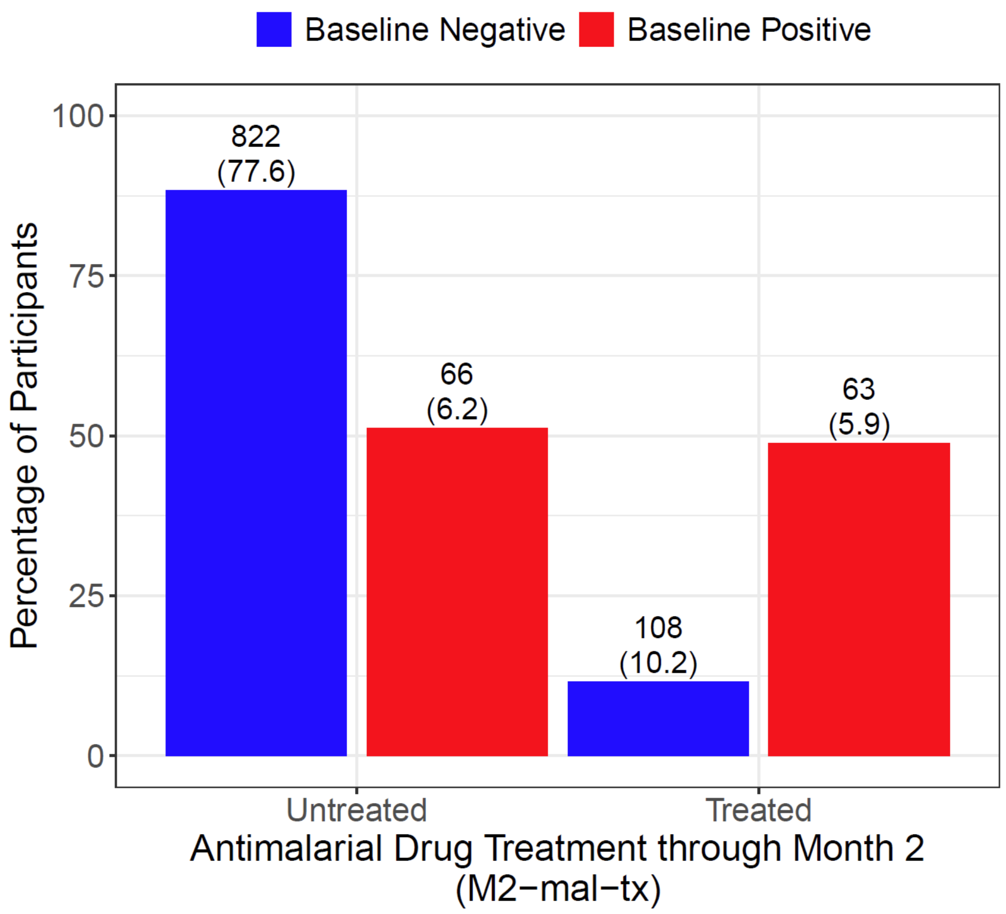

**Figure S17. Distribution of the onset of antimalarial drug treatment between the first vaccination and the month 2 scheduled visit (M2-mal-tx) separately among PP baseline negative vs. baseline positive participants in the pooled control, R012-14, R012-20, and Fx012-14 groups. For percentages in parentheses, the denominator is all PP participants in the pooled control, R012-14, R012-20, and Fx012-14 groups. Spearman's correlation between baseline positivity and M2-mal-tx was 0.33 (P<0.001).** PP: Per-Protocol

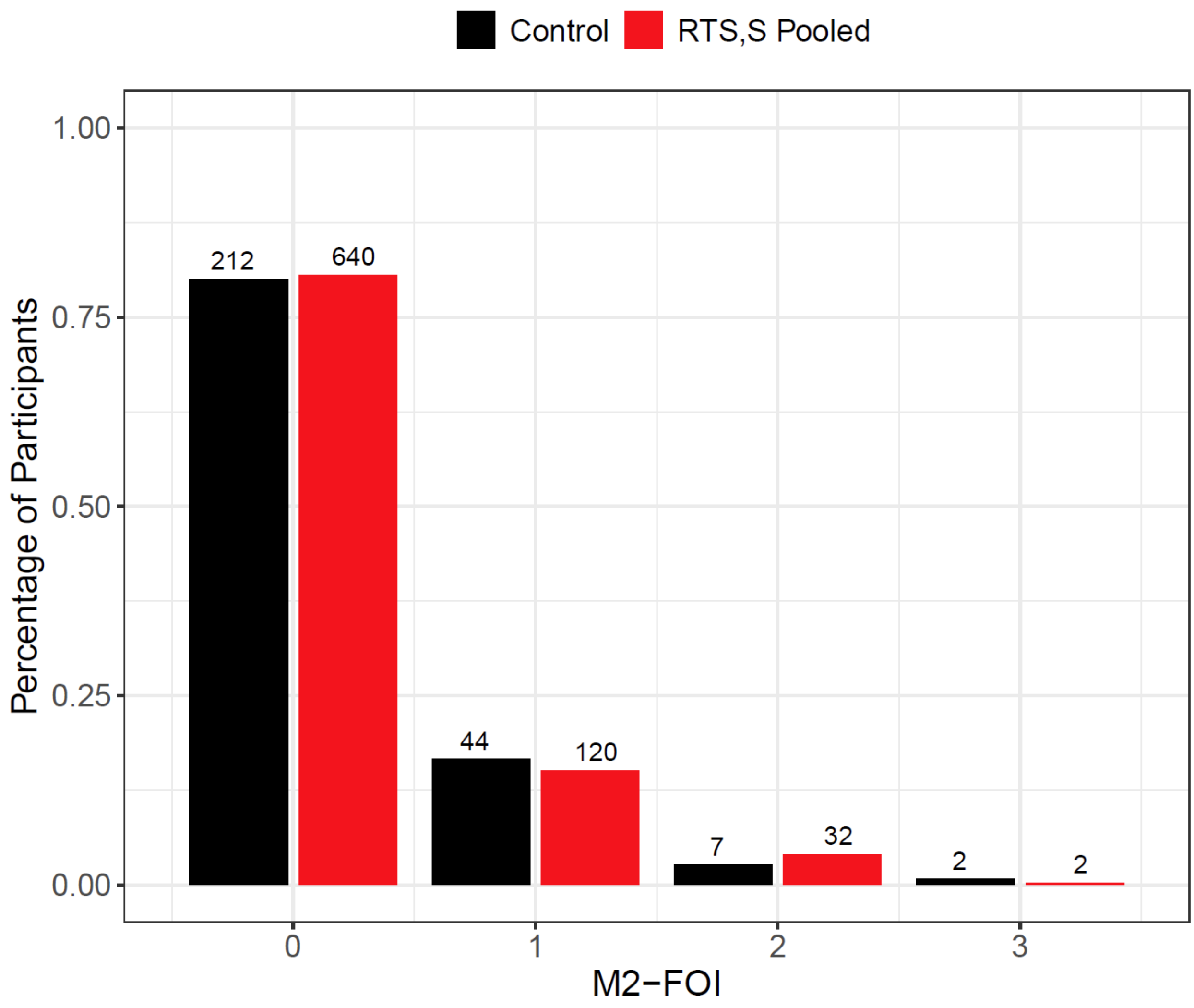

**Figure S18. Distribution of M2-FOI among Per-Protocol participants separately in the control vs. pooled R012-14, R012-20, and Fx012-14 groups.**
M2-FOI: month 2 force of infection.

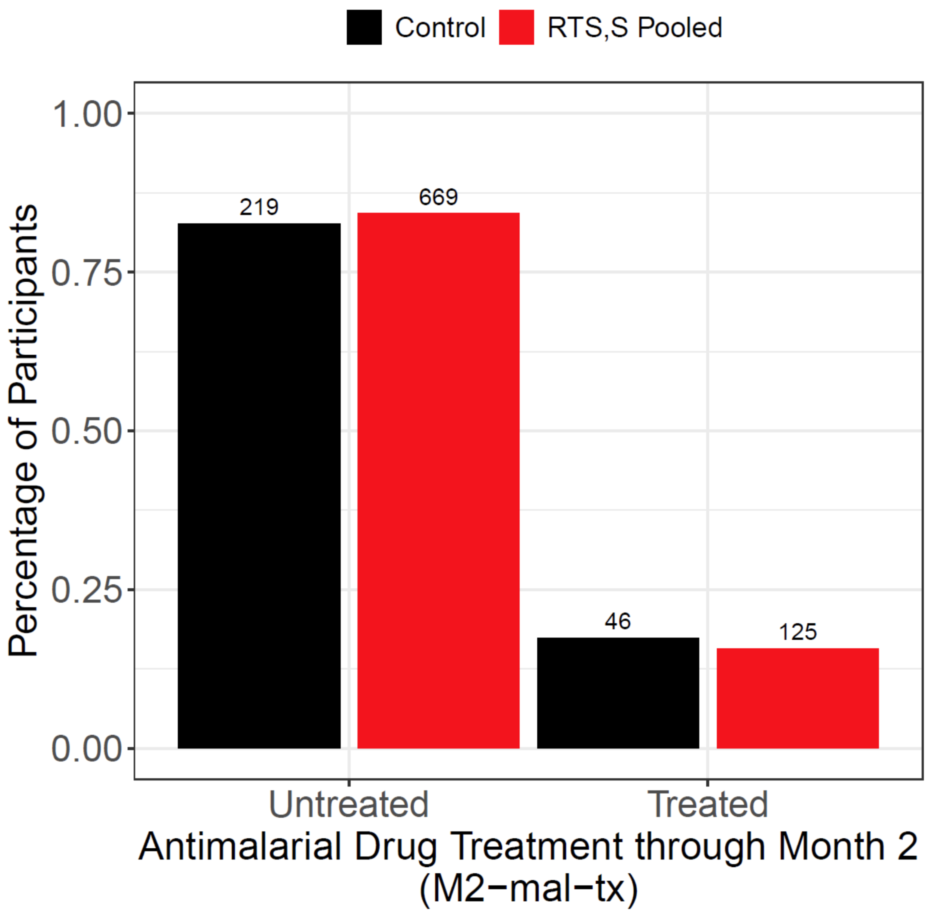

**Figure S19. Distribution of the onset of antimalarial drug treatment between the first vaccination and the month 2 scheduled visit (M2-mal-tx) among Per-Protocol participants separately in the control vs. pooled R012-14, R012-20, and Fx012-14 groups.**

| | | | Interaction P-value | |
|---|---|---|---|---|
| Model* | Covariates Adjusted for | AIC | Treatment × Baseline Positivity | Treatment × M2-FOI |
| M1-PP | Tx, BP, Tx × BP | 6997.4 | 0.0029 | – |
| M2-PP | Tx, BP, Tx × BP, ordinal M2-FOI | 6992.7 | 0.0053 | – |
| M3-PP | Tx, BP, ordinal M2-FOI, Tx × BP, Tx × ordinal M2-FOI | 6992.0 | 0.073 | 0.093 |
| M4-PP | Tx, BP, ordinal M2-FOI, Tx × BP, Tx × ordinal M2-FOI, BP × ordinal M2-FOI | 6991.3 | 0.05 | 0.071 |
| M5-PP | Tx, I(M2-FOI>0), Tx × I(M2-FOI>0) | 6992.2 | – | 0.057 |
| M6-PP | Tx, I(M2-FOI>0), BP, Tx × I(M2-FOI>0) | 6993.3 | – | 0.059 |
| M7-PP | Tx, I(M2-FOI>0), BP, Tx × I(M2-FOI>0), Tx × BP | 6991.4 | 0.044 | 0.33 |
| M8-PP | Tx, I(M2-FOI>0), BP, Tx × I(M2-FOI>0), Tx × BP, BP × I(M2-FOI>0) | 6990.2 | 0.021 | 0.31 |

*Each Cox model was additionally adjusted for M2-mal-tx, sex, age, BMI, and hemoglobin

**Figure S20. Summary of Cox model evidence for vaccine efficacy (VE) (pooled R012-14, R012-20, and Fx012-14 vs. control) against the first new infection modified by baseline positivity (models M1-PP–M4-PP) and by M2-FOI (models M5-PP–M8-PP), with model quality assessed using Akaike's Information Criterion (AIC).**

Tx: indicator of randomized assignment to an RTS,S regimen; BP: indicator of baseline parasite positivity; M2-FOI: month 2 force of infection; M2-mal-tx: indicator of the onset of antimalarial drug treatment between the first vaccination and the month 2 scheduled visit; BMI: body mass index

| Baseline Positivity | No. of First New Infections (Incidence per PYR) | VE (%) (95% CI) | | Interaction P−value |
|---|---|---|---|---|
| Negative | 124 (0.59) vs. 71 (1.10) | 49.9 (32.6, 62.7) | | 0.083 |
| Positive | 25 (0.90) vs. 20 (3.25) | 72.2 (49.5, 84.7) | | |

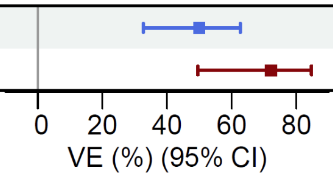

**Figure S21. Vaccine efficacy (VE) against the first new genotypic infection in the restricted period between 14 days and 4.5 months after the third dose for the pooled R012-14, R012-20, and Fx012-14 RTS,S regimens vs. the control regimen in subgroups of the Per-Protocol cohort defined by the baseline parasite positivity status while adjusting for the main effects of M2-FOI, M2-mal-tx, sex, age, and baseline levels of BMI and hemoglobin.**

PYR: person-years at risk; M2-FOI: month 2 force of infection; M2-mal-tx: indicator of the onset of antimalarial drug treatment between the first vaccination and the month 2 scheduled visit; No: number; CI: confidence interval; BMI: body mass index

| Study Site | Baseline Positivity | No. of First New Infections (Incidence per PYR) | VE (%) (95% CI) | Interaction P−value |
|---|---|---|---|---|
| Agogo | Negative | 145 (0.55) vs. 59 (0.69) | 23.8 (−3.4, 43.8) | 0.4 |
| | Positive | 19 (0.94) vs. 9 (1.97) | 48.0 (−20.1, 77.5) | |
| Siaya | Negative | 229 (1.30) vs. 89 (2.10) | 44.0 (28.1, 56.4) | 0.03 |
| | Positive | 49 (1.43) vs. 19 (4.03) | 70.9 (50.3, 83.0) | |

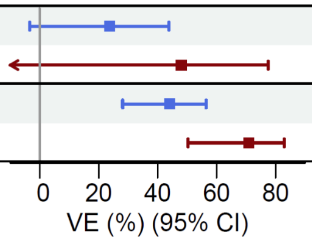

**Figure S22. Study site-specific vaccine efficacy (VE) against the first new genotypic infection between 14 days and 12 months after the third dose for the pooled R012-14, R012-20, and Fx012-14 RTS,S regimens vs. the control regimen in subgroups of the Per Protocol cohort defined by the baseline parasite positivity status while adjusting for the main effects of M2-FOI, M2-mal-tx, sex, age, and baseline levels of BMI and hemoglobin.** PYR: person-years at risk; M2-FOI: month 2 force of infection; M2-mal-tx: indicator of the onset of antimalarial drug treatment between the first vaccination and the month 2 scheduled visit. No: number; CI: confidence interval; BMI: body mass index

| Comparison | Baseline Positivity | No. of First New Infections (Incidence per PYR) | VE (%) (95% CI) | Interaction P−value |
|---|---|---|---|---|
| R012−20 plus R012−14 vs. Control | Negative | 226 (0.79) vs. 148 (1.16) | 41.2 (27.4, 52.4) | 0.083 |
| | Positive | 56 (1.37) vs. 28 (3.02) | 62.5 (40.4, 76.3) | |
| Fx012−14 vs. Control | Negative | 148 (0.95) vs. 148 (1.16) | 28.2 (9.3, 43.1) | < 0.001 |
| | Positive | 12 (0.89) vs. 28 (3.02) | 79.5 (59.2, 89.7) | |

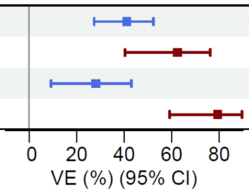

**Figure S23. Vaccine efficacy (VE) of individual RTS,S regimens vs. the control regimen against the first new genotypic infection among PP baseline negative vs. baseline positive participants while adjusting for the main effects of M2-FOI, M2-mal-tx, sex, age, and baseline levels of BMI and hemoglobin.**

PYR: person-years at risk; M2-FOI: month 2 force of infection; M2-mal-tx: indicator of the onset of antimalarial drug treatment between the first vaccination and the month 2 scheduled visit.
PP: Per-Protocol; No: number; BMI: body mass index

| Baseline Positivity | No. of First New Infections (Incidence per PYR) | VE (%) (95% CI) | Interaction P−value |
|---|---|---|---|
| Negative | 176 (1) vs. 71 (1.5) | 37.3 (16.9, 52.6) | 0.011 |
| Positive | 66 (1.3) vs. 26 (3) | 68.7 (50.0, 80.3) | |

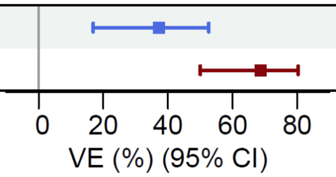

**Figure S24. Vaccine efficacy (VE) against the first new genotypic infection between 14 days and 12 months after the third dose for the pooled R012-14, R012-20, and Fx012-14 regimens vs. the control regimen in the subcohort of the Per-Protocol cohort restricted to baseline positive participants and three matched baseline negative participants from the same randomization group and study site for each baseline positive participant, with matching performed on the third vaccination date. Subgroups are defined by the baseline parasite positivity status. Model adjusted for the main effects of M2-FOI, M2-mal-tx, sex, age, and baseline levels of BMI and hemoglobin.**

The sampling of matched baseline negative participants was repeated 1000 times, and sample medians of the estimated VEs and 95% confidence limits are shown.

M2-FOI: month 2 force of infection; M2-mal-tx: indicator of the onset of antimalarial drug treatment between the first vaccination and the month 2 scheduled visit. No: number; PYR: person-years at risk; CI: confidence interval; BMI: body mass index

| Subgroup | Treatment Group | No. of PP Participants with Clinical Endpoint between M2.5–14 | Parasite Positive by M2[a], N (%) | ≥1 Haplotype in Sample from First New M2.5–14 Clinical Endpoint Detected in Samples by M2[b], N (%) |
|---|---|---|---|---|
| All | Pooled M0, 1, 2 Regimens | 342 | 150 (43.9) | 42 (12.3) |
| All | Pooled RTS,S M0, 1, 2 Regimens | 232 | 110 (47.4) | 29 (12.5) |
| All | Control | 110 | 40 (36.4) | 13 (11.8) |
| All | R012-20 | 69 | 38 (55.1) | 9 (13.0) |
| All | R012-14 | 71 | 37 (52.1) | 7 (9.9) |
| All | Fx012-14 | 92 | 35 (38.0) | 13 (14.1) |
| Baseline Negative | Pooled M0, 1, 2 Regimens | 280 | 88 (31.4) | 24 (8.6) |
| Baseline Negative | Pooled RTS,S M0, 1, 2 Regimens | 188 | 66 (35.1) | 18 (9.6) |
| Baseline Negative | Control | 92 | 22 (23.9) | 6 (6.5) |
| Baseline Negative | R012-20 | 48 | 17 (35.4) | 4 (8.3) |
| Baseline Negative | R012-14 | 60 | 26 (43.3) | 6 (10.0) |
| Baseline Negative | Fx012-14 | 80 | 23 (28.7) | 8 (10.0) |
| Baseline Positive | Pooled M0, 1, 2 Regimens | 62 | 62 (100.0) | 18 (29.0) |
| Baseline Positive | Pooled RTS,S M0, 1, 2 Regimens | 44 | 44 (100.0) | 11 (25.0) |
| Baseline Positive | Control | 18 | 18 (100.0) | 7 (38.9) |
| Baseline Positive | R012-20 | 21 | 21 (100.0) | 5 (23.8) |
| Baseline Positive | R012-14 | 11 | 11 (100.0) | 1 (9.1) |
| Baseline Positive | Fx012-14 | 12 | 12 (100.0) | 5 (41.7) |
| M2-FOI = 0 | Pooled M0, 1, 2 Regimens | 234 | 42 (17.9) | 6 (2.6) |
| M2-FOI = 0 | Pooled RTS,S M0, 1, 2 Regimens | 154 | 32 (20.8) | 4 (2.6) |
| M2-FOI = 0 | Control | 80 | 10 (12.5) | 2 (2.5) |
| M2-FOI = 0 | R012-20 | 41 | 10 (24.4) | 0 (0.0) |
| M2-FOI = 0 | R012-14 | 46 | 12 (26.1) | 0 (0.0) |
| M2-FOI = 0 | Fx012-14 | 67 | 10 (14.9) | 4 (6.0) |
| M2-FOI > 0 | Pooled M0, 1, 2 Regimens | 108 | 108 (100.0) | 36 (33.3) |
| M2-FOI > 0 | Pooled RTS,S M0, 1, 2 Regimens | 78 | 78 (100.0) | 25 (32.1) |
| M2-FOI > 0 | Control | 30 | 30 (100.0) | 11 (36.7) |
| M2-FOI > 0 | R012-20 | 28 | 28 (100.0) | 9 (32.1) |
| M2-FOI > 0 | R012-14 | 25 | 25 (100.0) | 7 (28.0) |
| M2-FOI > 0 | Fx012-14 | 25 | 25 (100.0) | 9 (36.0) |

[a] Either genotypic or microscopic detection of parasite positivity in samples collected before or at the month 2 visit

[b] At least 1 haplotype in the sample associated with the first new clinical malaria episode between M2.5–14 was detected in samples collected before or at the month 2 visit

**Table S5. First new clinical malaria episodes in Per-Protocol (PP) participants as potentially persistent asymptomatic infections acquired before the third vaccination.** Percentages in parentheses use the number of PP participants with a clinical malaria episode between M2.5–14 as the denominator. No: number; N: number; M2-FOI: month 2 force of infection

### Statistical Analysis Plan for:

### Molecular Detection and Genotyping of Plasmodium falciparum Parasites in Young African Children after Immunization with RTS,S/AS01$_E$ Malaria Vaccine

#### Protocol Number: 206213 (MALARIA-095)

An Ancillary Study Protocol to GlaxoSmithKline Phase IIb RTS,S/AS01$_E$ Malaria Vaccine Trial (Study Number 204889 [MALARIA-094]) entitled, "Efficacy, safety and immunogenicity study of GSK Biologicals candidate malaria vaccine (SB257049) evaluating schedules with or without fractional doses, early dose 4 and yearly doses, in children 5–17 months of age."

August 25, 2023

Michal Juraska[a], Ph.D.
David Benkeser[b], Ph.D.
Peter B. Gilbert[a], Ph.D.
Li Li[a], Ph.D.

[a]Department of Biostatistics, Bioinformatics, and Epidemiology, Vaccine and Infectious Disease Division, Fred Hutchinson Cancer Center
[b]Department of Biostatistics and Bioinformatics, Emory University

Version 1.7

### SAP MODIFICATION HISTORY

The version history of, and modifications to, this statistical analysis plan are described below.

Date: June 2, 2021
SAP version: 1.0

Date: October 13, 2021
SAP version: 1.1

Modifications:

- Section 3: Modified the ATP cohort definition as follows:

  "The According-To-Protocol (ATP) cohort includes all participants who received the first three vaccinations according to the MAL-094 protocol procedures and who are observed to be at risk for ~~the first new~~ a molecularly confirmed malaria infection at 14 days post-Dose 3."

  Deleted the sentence

  "The ATP cohort is a subset of the TVC and excludes participants with first new molecularly confirmed malaria infection detected prior to or at 14 days post-Dose 3."

- Section 5.1.5: Deleted the sentence "Because 7 months post dose three and Month 14 are not at immunization study visits, the analyses use exact dates to calculate failure time values (to the day)." because it posed an issue in operationalizing the condition "50% of scheduled visits" in the definition of the observed/known primary endpoint 2 due to missed scheduled visits and variability in within-window visit times. Any operational definition required a departure from this sentence. It was concluded that endpoints within the shorter-term follow-up period should be calculated in the same way as those within the primary follow-up period.

- Section 7.5: Revised the sentence

  "The cumulative distribution functions will be estimated in two ways: first, by stratifying by study site using the Kaplan-Meier estimator, and second, by adjusting for other baseline participant characteristics via targeted maximum likelihood estimation (TMLE, Moore and van der Laan, 2009)."

  as

  "The cumulative distribution functions will be estimated in two ways: first, by using the Nelson-Aalen estimator for the cumulative hazard function, and second, by adjusting for other baseline participant characteristics via targeted maximum likelihood estimation (TMLE, Moore and van der Laan, 2009)."

The Kaplan-Meier estimator was replaced with the Nelson-Aalen estimator because the simultaneous confidence band program is available for the Nelson-Aalen estimator only. This program does not accommodate stratification, therefore an unstratified estimator is considered, which matches the kernel-smoothed estimation of instantaneous hazard functions and their contrasts, which is also unstratified.

- Section 7.5: In the last paragraph, revised the sentence

  "These analyses will be done for each individual and pair of study groups specified in Tables 2, 3, and 4, except the analyses comparing active vaccine study groups and using the control group for bias-correction from possible secular trends will not be done (i.e., comparisons denoted by asterisks in Tables 2, 3, 4)."

  as

  "These analyses will be done for each individual and pair of study groups specified in Table 5, analogously to the analysis of instantaneous kernel-smoothed hazard functions and their contrasts."

Date: April 4, 2022
SAP version: 1.2

Modifications:

- Added Section 9 on post-hoc exploratory analyses.

Date: October 24, 2022
SAP version: 1.3

Modifications:

- Added Section 5.1.1.1 describing the modified post-hoc definition of molecular parasite positivity for application in Month 32 analyses.

- Added the statement

  "All analyses using follow-up through Month 32 will use the modified post-hoc definition of molecular parasite positivity as described in Section 5.1.1.1."

  at the beginning of Section 7.7 to reiterate the difference in the SAP-specified read threshold for molecular parasite positivity in analyses through Month 20 vs. Month 32 of follow-up.

Date: March 8, 2023
SAP version: 1.4

Modifications:

- Added Section 9.1 on sensitivity of overall VE estimates to the read count threshold for parasite positivity.
- Added Section 9.4 on the post-hoc analysis of modification of RTS,S vaccine efficacy by baseline parasite positivity.

Date: April 7, 2023
SAP version: 1.5

Modifications:

- Section 8: Revised the comparison of all active arms pooled vs. control to be active arms GP2–GP4 vs. control in all ATP sieve analyses in pursuit of exploratory objective 1.
- Section 8.4.2: Clarified one- vs. two-sided hypothesis testing and statistical significance in the following statements:

  "For the binary haplotype-level features in (a)–(c), we will estimate genotype-specific hazard-ratio (relative) VE with 95% CIs and a two-sided test for differential genotype-specific (relative) VE using…"

  "One-sided testing will be performed for comparisons to the control GP1 regimen of 3D7 Hamming distances, and two-sided testing will be performed for head-to-head RTS,S regimen comparisons of 3D7 Hamming distances as well as for all comparisons of all other quantitative sequence features. Statistical significance is defined as in Section 7.8 except, for multiplicity sets in which one-sided testing is performed, the same definition of FWER and FDR statistical significance is applied to doubled one-sided p-values."

- Added Section 8.4.3 specifying hypothesis testing multiplicity sets considered in the sieve analysis for primary endpoint 1.

Date: July 31, 2023
SAP version: 1.6

Modifications:

- Revised Section 9.4 to formalize and align statistical analyses with the separate scientific questions about baseline parasite positivity and month 2 force of infection (M2-FOI) as potential VE modifiers of interest.

- Added Section 9.4.2.3 describing a sensitivity analysis to unmeasured confounding.

Date: August 25, 2023
SAP version: 1.7

Modifications:

- Added in Section 9.4 the indicator of the onset of antimalarial drug treatment between the first vaccination and the month 2 scheduled visit as an additional adjustment covariate in the Cox models to control for potential confounding.

5.1.6 Primary Endpoints Assessed over Longer-term Follow-up through Month 32 19

7.6.1 Handling of Loss to Follow-up and Missing Visit Data for Primary Endpoint 2 28

9.4.2.2 Cox Proportional Hazards Analysis for Assessing M2-FOI as a VE Modifier 47

### 1 SYNOPSIS

The goal of the amplicon sequencing and genotyping study (hereafter referred to as 'genotyping study') is to assess vaccine efficacy against molecularly confirmed malaria infection using ultra-sensitive molecular amplification and sequencing methodology to detect Plasmodium falciparum (P. falciparum) parasites from serial blood samples to be collected from children immunized with the primary and yearly booster immunizations of the RTS,S/AS01E vaccine as part of their participation in Protocol MALARIA-094 (MAL-094). Genomic analysis will be performed on parasites from blood spot samples collected from children aged 5-17 months when first immunized with RTS,S/AS01E on different dosage and schedule regimens under the MAL-094 clinical study parent protocol (Study Number 204889), entitled "Efficacy, safety and immunogenicity study of GSK Biologicals' candidate malaria vaccine (SB257049) evaluating schedules with or without fractional doses, early dose 4 and yearly doses, in children 5-17 months of age."

The MAL-094 study, being conducted at the Agogo, Ghana and Siaya, Kenya study sites in East and West Africa, assesses proof of concept for a fractional (Fx) dose schedule under conditions of natural exposure. The study also assesses the role of third dose spacing in a Fx dose schedule, the effect of an earlier full fourth dose at Month 14, and the effect of multiple Fx versus full yearly doses. The study is conducted in children 5-17 months of age at first vaccination living in areas of moderate to high malaria transmission. In addition to assessing vaccine efficacy against clinical malaria disease, this study will for the first time assess vaccine efficacy against asymptomatic and submicroscopic parasitemia in dried blood spots collected in monthly surveys from every child enrolled in this study using ultra-sensitive molecular genotyping methods. A full description of the design and sampling methodology is described in the MAL-094 trial protocol.

In the present MAL-095 study, samples generated by the MAL-094 trial are used in combination with cutting-edge genomic and statistical analysis to generate a better understanding of the RTS,S/AS01E vaccine. Whereas traditional PCR-based infection screens yield a simple binary outcome of positive or negative infection status, amplicon sequencing data provide information on the complexity of infection (COI), a measure of the number of genetically distinct parasite lineages in each participant sample in the trial, and can be used to estimate the rate at which new infections occur and parasite lineages turn over. The resulting data yield not only the infection status of each participant, but also information on the incidence rates of new malaria infection. These data allow assessment of VE of each vaccine study arm to prevent new genetically distinct malaria infection.

In addition to supporting the primary objective of evaluating differences in overall VE against new malaria infection among the arms of the MAL-094 trial, amplicon sequencing data enable investigations into the nature of immunity generated by vaccination and natural disease exposure. The longitudinal sampling design allows for exploration of whether and how prior infection history impacts participants' susceptibility to new infections. There will also be assessment of the degree to which allele-specific protection contributes to overall VE, and how VE depends on genetic distance of infecting parasites to the vaccine strain, as performed in the MALARIA-055 ancillary genotyping study (Benkeser, Juraska, and Gilbert, 2018).

The design of the MAL-094 trial, in combination with the unique data generation capacity developed at the Broad Institute for previous RTS,S work, creates an opportunity to significantly improve the efficacy and understanding of RTS,S/AS01$_E$ vaccination, and to investigate the dynamics of naturally acquired immunity to malaria infection.

Enrollment of MAL-094 began on October 16, 2017 in Agogo and on May 18, 2018 in Siaya. Enrollment was completed at Agogo on February 6, 2018 and on September 25, 2018 in Siaya. The two planned MAL-095 data analyses will be conducted after all enrolled participants reach the Month 20 visit (which occurred in June, 2020) and after all enrolled participants reach the Month 32 visit (which is estimated to occur in July, 2021).

Given similarities in the molecular data generation and statistical approaches between this project and the ancillary genotyping study performed for the phase 3 MALARIA-055 study (MALARIA-066), there is confidence to perform the necessary data generation and analyses to accomplish the study objectives.

### 2 RANDOMIZED TREATMENT ARMS

This study is an ancillary study of the GlaxoSmithKline (GSK) RTS,S/AS01E Phase IIb randomized trial (Study Number 204889) that is performed across both study sites Agogo and Siaya. The original randomized trial (MAL-094) vaccinates approximately 1,500 subjects 5-17 months of age in multi-centers according to the following groups:

(GP1) Control group, i.e., rabies vaccine at Month 0, Month 1, Month 2;

(GP2) Group R012-20 - a course of RTS,S/AS01E full dose at Month 0, Month 1, Month 2, and a full dose at Month 20;

(GP3) Group R012-14-mD - a course of RTS,S/AS01E full dose at Month 0, Month 1, Month 2, and yearly full doses at Month 14, Month 26, Month 38;

(GP4) Fractionated (Fx) Group Fx012-14-mFxD - a course of RTS,S/AS01E full dose at Month 0 and Month 1, followed by RTS,S/AS01E 1/5th dose at Month 2, Month 14, Month 26, and Month 38.

(GP5) Group Fx017-mFxD - a course of RTS,S/AS01E full dose at Month 0 and Month 1, followed by RTS,S/AS01E 1/5th dose at Month 7, Month 20, and Month 32.

A total of 300 subjects are randomly allocated to each treatment group (Figure 1 below from the MAL-094 protocol).

Figure 1.  MAL-094 protocol describing study arms and study visits

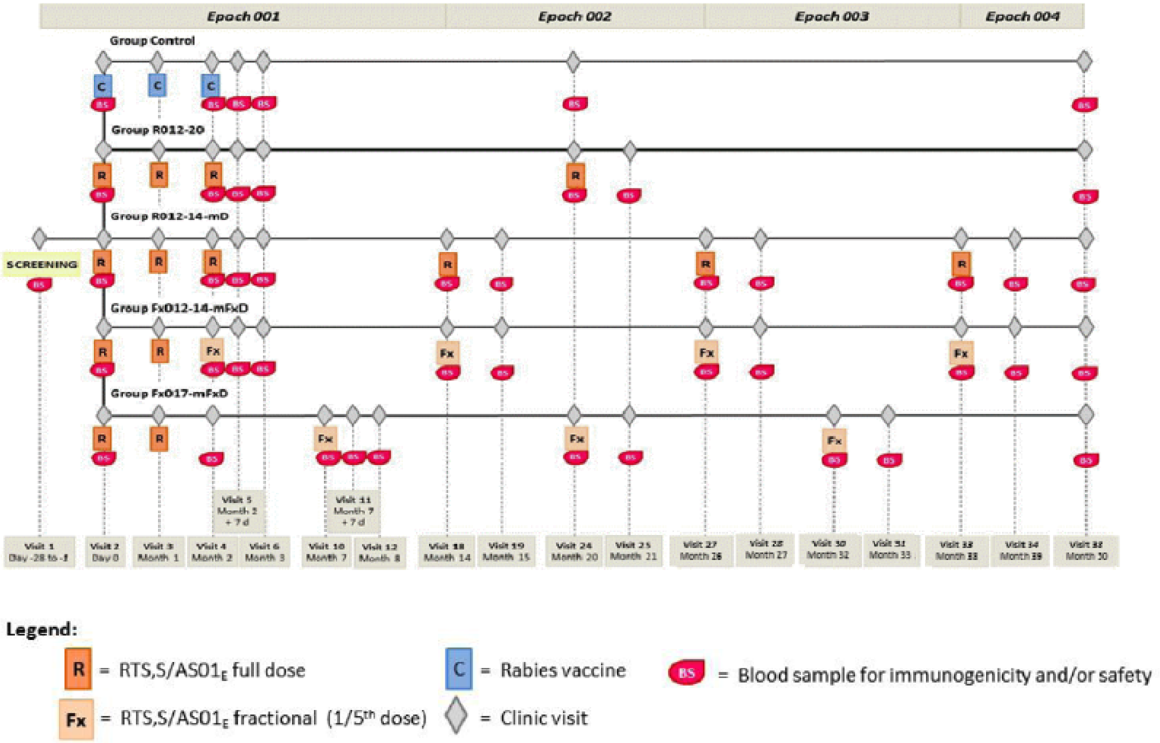

### 3 STUDY COHORTS

Analyses will be conducted in two distinct study cohorts:

(Cohort 1) The According-To-Protocol (ATP) cohort includes all participants who received the first three vaccinations according to the MAL-094 protocol procedures and who are observed to be at risk for a molecularly confirmed malaria infection at 14 days post-Dose 3.

(Cohort 2) The Total Vaccinated Cohort (TVC) is all participants who received at least one dose of a study vaccine.

Primary and exploratory analyses of vaccine efficacy will be conducted both in ATP and TVC cohorts, with participants analyzed according to the as-treated principle (i.e., actually received treatments). The same ATP cohort will be analyzed in vaccine efficacy analyses of both primary study endpoints defined in Sections 5.1.4 and 5.1.5.

### 4 OBJECTIVES

#### 4.1 Context for MAL-095 Objectives Relative to MAL-094 Parent Protocol Objectives

The primary efficacy objective of MAL-094 assesses relative vaccine efficacy against clinical malaria over one year post Dose 3 of the Group 4 fractional dosing regimen versus the standard RTS,S/AS01E regimen. Note that the latter standard regimen consists of pooling participants in Groups 2 and 3, as these groups have the same vaccination schedule through the Month 14 visit, and the randomization principle holds. The MAL-094 study additionally includes secondary objectives to study vaccine efficacy and relative vaccine efficacy against clinical malaria and infection for all of the study arms.

A central contribution of MAL-095 is to provide more robust and extensive assessment of vaccine efficacy and relative vaccine efficacy against molecularly confirmed malaria infection. The MAL-095 study is the first trial to rigorously study vaccine efficacy and relative vaccine efficacy against malaria infection using molecular techniques. MAL-095 uses an analysis structure that gives special attention to the comparison of novel RTS,S/AS01E vaccine regimens to the standard RTS,S/AS01E regimen, as noted below.

#### 4.2 Primary Objective

The primary objective of MAL-095 is to estimate and compare vaccine efficacy (VE) against new molecularly confirmed malaria infection(s) across parasite genotypes among the vaccine arms of the trial, using amplicon sequencing and genotyping to detect new infections (see Section 5.1.1 for the definition of a new infection). Infections will be detected by molecular methods from monthly samples or samples collected at unscheduled visits for clinical assessment.

Throughout, vaccine efficacy refers to a comparison of a study endpoint between an active RTS,S vaccine regimen (one of Groups 2 through 5) vs. the rabies control arm, and relative vaccine efficacy refers to a comparison of a study endpoint between two active RTS,S vaccine regimens (among Groups 2 through 5). Vaccine efficacy and relative vaccine efficacy are assessed based on two types of outcomes:

(Primary endpoint 1) first new molecularly confirmed ***P. falciparum*** infection detection over a specified follow-up period;

(Primary endpoint 2) all new molecularly confirmed ***P. falciparum*** infection detections accumulated over a specified follow-up period.

As stated in the MAL-095 protocol, vaccine efficacy and relative vaccine efficacy against each primary outcome type will be assessed in analyses of both the ATP and TVC cohorts, as co-primary objectives.

The follow-up periods for assessing the two outcome types in the primary and exploratory analyses, for each of the ATP and TVC cohort analyses, are specified in Section 5.

#### 4.3 Exploratory Objectives

The following exploratory objectives will be addressed for both outcome types:

1. Exploratory objective 1: To assess whether and how VE against new malaria infection based on molecular detection (active case detection) depends on genotypic characteristics of the exposing malaria parasites.

2. Exploratory objective 2: To study whether and how prior infection and/or vaccination has a relationship to subsequent infection by measuring the molecular Force of Infection (mol[FOI]) and assessing the relationship between mol[FOI] and subsequent malaria infection risk. The variable mol[FOI] is defined in Section 6.

3. Exploratory objective 3: To assess whether and how prior infection by a particular parasite genotype, not necessarily a 3D7 vaccine genotype, reduces the likelihood of re-infection by a parasite with the same genotype.

### 5 STUDY ENDPOINTS

#### 5.1 Primary Endpoints

##### 5.1.1 Definition of New Malaria Infection

Following vaccination, the study will distinguish new from existing infections using an amplicon sequencing-based strategy that involves deep sequencing of small, highly variable regions of the parasite genome. This sequencing allows both:

1. highly sensitive detection of parasitemia (analogous to conventional PCR-based detection), and

2. identification of genetically distinct parasite populations within and between affected individuals.

A new molecularly confirmed malaria infection endpoint is defined by a detected new infection from genomic analysis of dried blood spot samples originating either from active monthly screening for infection or from unscheduled visits intended for the assessment of clinical malaria; thus, molecular detection of a new malaria infection measures either an asymptomatic/subclinical or clinical malaria infection. Later in this section the definition of “new malaria infection” is provided.

Genotype data for the ***P. falciparum*** circumsporozoite (CS) C-terminus and SERA2 loci are measured from each dried blood spot sample, from which the observed haplotype(s) at each amplicon following filtration of PCR and sequencing errors are defined. However, data from certain samples at certain study visits may be missing or excluded from the final data analysis, for reasons including the sample: is not collected; is not collected with full or clear identification; does not yield any parasite genetic material upon process; or does not yield a clear and unambiguous genotyping result for any reason. In other words, for some samples it will not be possible to define the observed haplotype(s) at each amplicon in a sufficiently reliable fashion to use the data for analysis.

Associated with each sample with molecular detection of malaria infection is the complexity of infection (COI), defined as the maximum of (i) the number of unique haplotypes for the CS C-terminus amplicon, and (ii) the number of unique haplotypes for the SERA2 amplicon in the sample. It is expected that the cause of $n = COI > 1$ is multiple distinct malaria infections; thus, COI itself is one estimate of the number of new molecularly confirmed malaria infections that have occurred up to the time of the given sample. However, simultaneous super-infection is possible, and some new malaria infections could be undetected if enough time elapses between infection and sampling, or if antimalarial treatment clears an infection.

The following paragraphs provides more detail on the definition of a new malaria infection.

*Malaria infection*
A ***P. falciparum*** infection is defined as amplicon nucleotide sequence-based parasite positive status defined as detection of at least one haplotype supported by at least 50 nucleotide sequence reads and 1% of total reads for at least one of the two amplicons.

*New malaria infection*
As there is no guarantee of complete clearance of infections, a new infection may originate from a post-enrollment sample that goes from negative to positive, or from a post-enrollment sample that contains a new haplotype that was not previously observed in several previous samples from the same participant. A new infection for the primary objective is defined as at least one of the haplotypes in the post-enrollment sample not being detected in the previous three samples drawn from the same participant regardless of the visit type (scheduled or unscheduled). An exception is made if a haplotype is detected in the first post-enrollment sample not collected on the day of enrollment, in which case it counts as a new infection if the same haplotype was not detected in the single sample from the enrollment visit. Haplotype detection requires observation of at least 50 reads and 1% of total reads supporting the haplotype within a sample. Note that a molecularly confirmed infection in a sample from the enrollment visit does not count as a new infection because the sample was collected prior to administration of the first dose, and therefore the sample should not contribute to measuring the vaccine effect. A molecularly confirmed infection in a sample from an unscheduled visit that occurred on the day of enrollment does not count as a new

infection either because we cannot rule out that it is a persistent infection acquired prior to enrollment.

###### 5.1.1.1 Modified Definition of Molecular Parasite Positivity for Month 32 Analyses

During inspection of sequencing data in support of the M20 interim analysis, a post hoc decision was made to revise the minimum read count for calling a haplotype as present in a sample from 50 to 325 sequencing reads. The 50 read threshold established during formulation of the original SAP v1.0 was chosen to optimize precision and recall using a truth set of sequencing data from mock samples. However, upon analysis of the much larger M20 trial dataset, it was determined that a revised threshold of 325 reads would more effectively reduce signals of false positive haplotype calls due to sporadic transfer of DNA among samples within sequencing batches housed in the same multi-well plate.

##### 5.1.2 Definition of Primary Endpoint 1

We define the time to primary endpoint 1 variable as the number of days from the time origin in each analysis to the visit date associated with the first molecular detection of a new malaria infection, irrespective of

- the number of missed scheduled visits preceding this event,
- whether this event occurred within a certain time interval following the onset of antimalarial treatment, and/or
- an earlier occurrence of secondary-case clinical malaria (with no molecular detection).

For participants with no molecular detection of a new infection at any visit within a considered follow-up period, their time to primary endpoint 1 will be right-censored at the collection date of the last parasite negative sample within the follow-up period in each analysis.

##### 5.1.3 Definition of Primary Endpoint 2

For a participant with no missed scheduled visits and no missed samples from attended visits, we estimate the total number of new molecularly confirmed malaria infections in a given follow-up period as the number of occurrences of molecular detection of a new malaria infection during this period regardless of

- whether any such detection occurred within a certain time interval following the onset of antimalarial treatment, and/or
- any occurrence(s) of secondary-case clinical malaria.

For a participant with missed scheduled visits or missed samples from attended visits, the definition of an observed/known primary endpoint 2 in a given follow-up period accounts for missing sequence data, and the definition is stated in Section 7.6.1.

A descriptive sensitivity analysis will be conducted to compare rates of primary endpoint 1, defined in Section 5.1.2, to those of first new infection by either molecular detection or passive detection of secondary-case clinical malaria per the MAL-094 protocol definition. The descriptive analysis will include tabulating proportions of molecularly detected new infections that are in samples from visits with confirmed secondary-case clinical malaria, and proportions of negative samples for clinical malaria ascertainment that yielded a positive result by the molecular method of detection. Clinical case detection is not included in the primary endpoint definitions under the premise that each infection requires a molecular presentation. Because an infection that was acquired pre-enrollment may first clinically manifest after enrollment, ignoring clinical cases in primary endpoint 1 precludes from erroneously calling a clinical infection the first new infection. Also, ignoring clinical infection events avoids double-counting of infections in primary endpoint 2.

##### 5.1.4 Primary Endpoints Assessed during Primary Follow-up

The two primary endpoints for the ATP cohort analyses are:

1. Time from 14 days post-Dose 3 to molecular detection of the first new malaria infection through to 12 months post-Dose 3

2. Number of new molecularly confirmed malaria infections during the same follow-up period used for ATP primary endpoint 1

The two primary endpoints for the TVC analyses are:

1. Time from first vaccination to molecular detection of the first new malaria infection through to the Month 20 study visit post first vaccination

2. Number of new molecularly confirmed malaria infections during the same follow-up period used for TVC primary endpoint 1

For the measurement of new malaria infection detection endpoints in the ATP cohort, "through to xx months post-Dose 3" (e.g., xx=12 months) means through to the Month yy study visit (e.g., if xx=12, then yy=14 for Groups 1-4 and yy=19 for Group 5). This approach rather than measurement of exactly xx months post-Dose 3 (irrespective of visit date) is used to ensure that follow-up for all study arms only includes time periods before Dose 4, which is administered at the Month 14 study visit for Groups 3 and 4 and at the Month 20 study visit for Groups 2 and 5. If the Month yy visit is missed, the time-to-event is right-censored at the last parasite negative sample collection date prior to the missed Month yy visit.

For analyses of the TVC, for both primary endpoints, new molecularly confirmed malaria infections are counted after the first vaccination through to the Month 20 visit, for all study arms and all comparisons. Infections detected in any sample collected on the day of enrollment (whether from the enrollment visit or an unscheduled visit completed on the day of enrollment) are excluded, i.e., not treated as new infections for either primary endpoint because, in these samples, we cannot rule out a persistent infection acquired prior to enrollment.

For analyses of the ATP cohort, for both primary endpoints, the comparator group (rabies control vaccination) window of follow-up for capturing outcomes differs depending on the vaccine group to which it is compared. In particular, the primary objectives in the ATP cohort study primary endpoints occurring after 14 days post-Dose 3 through to the visit scheduled 12 months post-Dose 3 (Month 14 for Groups 2, 3, 4 and Month 19 for Group 5), such that the follow-up period for registering events is Month 2.5-14 for each of Groups 2, 3, 4 vs. Control and is Month 7.5-19 for Group 5 vs. Control (the first row of Table 1 describes all of these follow-up periods).

For comparing the vaccine study groups head-to-head, all pairwise comparisons for the TVC analyses use a follow-up period starting from first vaccination through to the Month 20 study visit, as for the analyses of each RTS,S/AS01E vaccine arm vs. control. The ATP cohort analyses for these pairwise study group comparisons use a follow-up period of 11.5 months starting at 14 days post-Dose 3 (Table 1).

Table 1. Follow-up Intervals (in Months) Since First Vaccination for Counting New Malaria Infections for the Primary Analyses in the ATP Cohort (Each Entry is for the Comparison of Two Study Arms, where 2.5 indicates 14 days post Dose 3 in Groups 1-4, 7.5 indicates 14 days post Dose 3 in Group 5, 14 indicates the Month 14 study visit, and 19 indicates the Month 19 Study Visit)*

<table>
<tr><th></th><th>GP2 R012-20**</th><th>GP3 R012-14**</th><th>GP4 Fx012-14</th><th>GP5 Fx017</th></tr>
<tr><td>GP1 Cont.</td><td colspan="2">2.5-14 vs. 2.5-14</td><td>2.5-14 vs. 2.5-14</td><td>7.5-19 vs. 7.5-19</td></tr>
<tr><td>GP2 R012-20</td><td></td><td></td><td rowspan="2">2.5-14 vs. 2.5-14</td><td rowspan="2">2.5-14 vs. 7.5-19</td></tr>
<tr><td>GP3 R012-14</td><td></td><td></td></tr>
<tr><td>GP4 Fx012-14</td><td></td><td></td><td></td><td>2.5-14 vs. 7.5-19</td></tr>
</table>

*"Study visit" refers to a monthly longitudinal sampling visit through Month 20.
**GP2 and GP3 are pooled for comparison with GP1 over 2.5-14 months given GP2 and GP3 are identical during this follow-up period. Similarly, GP2 and GP3 are pooled for comparison with GP4 during 2.5-14 months and are pooled for comparison with GP5 (2.5-14 months for GP2 and GP3 vs. 7.5-19 months for GP5).

Table 2 lists the set of primary vaccine efficacy and relative vaccine efficacy analyses that are done, with follow-up periods, and grouping the analyses into three sets (vs. control, vs. the standard R012-20 vaccine regimen, and head-to-head novel vaccine regimens).

Table 2. Study Groups Compared to Assess Vaccine Efficacy or Relative Vaccine Efficacy Based on New Malaria Infection Endpoint Data Through Month 20 (Primary Endpoints)

| Analysis Type (Cohort) | Study Group and Follow-up Comparison |
|---|---|
| Assess Vaccine Efficacy (TVC) | GP2 M0-20 vs. GP1 M0-20 |
| | GP3 M0-20 vs. GP1 M0-20 |
| | GP4 M0-20 vs. GP1 M0-20 |
| | GP5 M0-20 vs. GP1 M0-20 |
| Assess Vaccine Efficacy (ATP) | GP2 + GP3 pooled M2.5-14 vs. GP1 M2.5-14 |

| | |
|---|---|
| | GP4 M2.5-14 vs. GP1 M2.5-14or T2 |
| | GP5 M7.5-19 vs. GP1 M7.5-19 |
| Assess Relative Vaccine Efficacy vs. Standard regimen R012-20 (TVC) | GP3 M0-20 vs. GP2 M0-20 |
| | GP4 M0-20 vs. GP2 M0-20 |
| | GP5 M0-20 vs. GP2 M0-20 |
| Assess Relative Vaccine Efficacy vs. Standard regimen R012-20 (ATP) | GP4 M2.5-14 vs. GP2 + GP3 pooled M2.5-14 |
| | GP5 M7.5-19 vs. GP2 + GP3 pooled M2.5-14* |
| Assess Relative Vaccine Efficacy (Other Comparisons) (TVC) | GP4 M0-20 vs. GP3 M0-20 |
| | GP5 M0-20 vs. GP3 M0-20 |
| | GP5 M0-20 vs. GP4 M0-20 |
| Assess Relative Vaccine Efficacy (Other Comparisons) (ATP) | GP5 M7.5-19* vs. GP4 M2.5-14 |

*Because different 11.5-month intervals are compared, the control group GP1 is used in the analysis to correct for potential bias due to secular trends (e.g., caused by seasonality); Section 7.5 provides details on the analysis.

##### 5.1.5 Primary Endpoints Assessed over Shorter-term Follow-up

The primary endpoints are also analyzed only counting new malaria infections through 14 months post first vaccination (TVC analyses) or through 7 months post Dose 3 (ATP analyses), when immunity may be greatest and hence protection putatively greater. The short-term endpoints are specified for the ATP cohort as:

1. Time from 14 days post-Dose 3 to molecular detection of the first new malaria infection through to 7 months post-Dose 3

2. Number of new molecularly confirmed malaria infections during the same follow-up period specified immediately above

Similarly, the short-term endpoints are specified for the TVC as:

1. Time from first vaccination to molecular detection of the first new malaria infection through to the Month 14 study visit after first vaccination

2. Number of new molecularly confirmed malaria infections during the same follow-up period specified immediately above

Table 3 shows the group comparisons and time periods of follow-up for analysis of the primary outcomes over shorter term follow-up.

Table 3. Study Groups Compared to Assess Vaccine Efficacy or Relative Vaccine Efficacy Based on New Malaria Infection Endpoint Data Through Month 14 (Analyses of the Primary Endpoints Shorter Term)

| Analysis Type | Study Group and Follow-up Comparison |
|---|---|
| Assess Vaccine Efficacy (TVC) | GP2 + GP3 pooled M0-14 vs. GP1 M0-14 |
| | GP4 M0-14 vs. GP1 M0-14 |
| | GP5 M0-14 vs. GP1 M0-14 |
| Assess Vaccine Efficacy (ATP) | GP2 + GP3 pooled M2.5-9 vs. GP1 M2.5-9 |
| | GP4 M2.5-9 vs. GP1 M2.5-9 |
| | GP5 M7.5-14 vs. GP1 M7.5-14 |
| Assess Relative Vaccine Efficacy vs. Standard regimen R012 (TVC) | GP4 M0-14 vs. GP2 + GP3 pooled M0-14 |
| | GP5 M0-14 vs. GP2 + GP3 pooled M0-14 |
| Assess Relative Vaccine Efficacy vs. Standard regimen R012(ATP) | GP4 M2.5-9 vs. GP2 + GP3 pooled M2.5-9 |
| | GP5 M7.5-14 vs. GP2 + GP3 pooled M2.5-9* |
| Assess Relative Vaccine Efficacy (Other Comparisons) (TVC) | GP5 M0-14 vs. GP4 M0-14 |
| Assess Relative Vaccine Efficacy (Other Comparisons) (ATP) | GP5 M7.5-14* vs. GP4 M2.5-9 |

*Similar to Table 2, because different 6.5-month intervals are compared, the control group GP1 is used in the analysis to correct for potential bias due to secular trends (e.g., caused by seasonality); Section 7.5 provides details on the analysis.

##### 5.1.6 Primary Endpoints Assessed over Longer-term Follow-up through Month 32

Longitudinal samples taken at Month 23, 26, 29, 32 are subjected to haplotyping in the same way as monthly samples drawn at monthly visits before and including Month 20.

To assess durability of VE, the primary endpoints are also analyzed (i) counting new molecularly confirmed malaria infections in the TVC cohort through 32 months after the first vaccination, and (ii) counting new molecularly confirmed malaria infections in the ATP cohort through 24 months post-Dose 3. Note that membership in the ATP cohort does not depend on whether Dose 4 is received.

The two primary endpoints assessed for durability for the ATP cohort analyses are:

1. Time from 14 days post-Dose 3 to molecular detection of the first new malaria infection through to 24 months after Dose 3

2. Number of new molecularly confirmed malaria infections during the same follow-up period used for the endpoint defined immediately above

The two primary endpoints assessed for durability for the TVC analyses are:

1. Time from first vaccination to molecular detection of the first new malaria infection through to the Month 32 study visit after first vaccination

2. Number of new molecularly confirmed malaria infections during the same follow-up period used for the endpoint specified immediately above

Table 4 summarizes the group comparisons and time periods for including new malaria infections using data though Month 32. Note that there is no scheduled Month 31 visit. Therefore, ATP cohort analyses listed in Table 4 that involve Group 5 will include new malaria infections detected in this group in the time period between 14 days post-Dose 3 and no later than the minimum of exactly 24 months post-Dose 3 and the time of completion of the Month 32 visit.

Table 4. Study Groups Compared to Assess Vaccine Efficacy or Relative Vaccine Efficacy Based on New Malaria Infection Endpoint Data Through Month 32 (Analysis of Primary Endpoints Longer Term)

| Analysis Type | Study Group and Follow-up Comparison |
|---|---|
| Assess Vaccine Efficacy (TVC) | GP2 M0-32 vs. GP1 M0-32 |
| | GP3 M0-32 vs. GP1 M0-32 |
| | GP4 M0-32 vs. GP1 M0-32 |
| | GP5 M0-32 vs. GP1 M0-32 |
| Assess Vaccine Efficacy (ATP) | GP2 M2.5-26 vs. GP1 M2.5-26 |
| | GP3 M2.5-26 vs. GP1 M2.5-26 |
| | GP4 M2.5-26 vs. GP1 M2.5-26 |
| | GP5 M7.5-31 vs. GP1 M7.5-31 |
| Assess Relative Vaccine Efficacy vs. Standard regimen R012-20 (TVC) | GP3 M0-32 vs. GP2 M0-32 |
| | GP4 M0-32 vs. GP2 M0-32 |
| | GP5 M0-32 vs. GP2 M0-32 |
| Assess Relative Vaccine Efficacy vs. Standard regimen R012-20 (ATP) | GP3 M2.5-26 vs. GP2 M2.5-26 |
| | GP4 M2.5-26 vs. GP2 M2.5-26 |
| | GP5 M7.5-31 vs. GP2 M2.5-26* |
| Assess Relative Vaccine Efficacy (Other Comparisons) (TVC) | GP4 M0-32 vs. GP3 M0-32 |
| | GP5 M0-32 vs. GP3 M0-32 |
| | GP5 M0-32 vs. GP4 M0-32 |
| Assess Relative Vaccine Efficacy (Other Comparisons) (ATP) | GP4 M2.5-26 vs. GP3 M2.5-26 |
| | GP5 M7.5-31 vs. GP3 M2.5-26* |
| | GP5 M7.5-31 vs. GP4 M2.5-26* |

*Similar to Tables 2 and 3, because different 23.5-month intervals are compared, the control group GP1 is used in the analysis to correct for potential bias due to secular trends (e.g., caused by seasonality); Section 7.5 provides details on the analysis.

#### 5.2 Two Tiers of Sample Evaluation

This subsection summarizes the dried blood spot samples that are drawn and subjected to genotyping/haplotyping, providing the amplicon data needed to define occurrence of new malaria infections; the section STUDY ENDPOINTS describes periods of follow-up for counting new malaria infections for analyses of vaccine efficacy and relative vaccine efficacy. The following sequential approach is used for sample evaluation to investigate the study objectives, with Tier 1 sampling essential to addressing the Primary Objective, and Tier 2 sampling used for exploratory analyses, where Tier 1 sample evaluation is performed after the primary immunization series (Doses 1, 2, 3) and Tier 2 sampling performed after the booster immunization series, respectively. Two sets of samples will be analyzed:

1. Tier 1: All dried blood spot samples collected monthly from all subjects during the first 20 months (expect approximately 31,500 samples) and all clinical malaria cases during the first 20 months (expect approximately 14,400 samples) from all treatment arms of the study.

2. Tier 2: All dried blood spot samples collected quarterly after the first 20 months for one year following the first booster RTS,S dose (expect approximately 9,000 samples) and clinical malaria cases after the first 20 months through Month 32 (expect approximately 8,640 samples) from all treatment arms of the study.

### 6 VARIABLES OF INTEREST

#### 6.1 Independent Variable

The primary covariate of interest is treatment assignment.

#### 6.2 Candidate Adjustment Variables

Some of the statistical analyses described in the STATISTICAL ANALYSIS section make use of baseline and post-baseline participant-level variables that may be prognostic for subsequent new malaria infection. In particular, the following baseline variables are accounted for in development of a “baseline risk score” variable that is predictive of the rate of new malaria infection and in covariate-adjusted analyses such as conducted by targeted maximum likelihood estimation:

1. Study site

2. Age
3. Sex

In addition, the set of post-baseline time-dependent variables that may be prognostic for malaria infection and may be accounted for in some statistical analyses are:

1. mol[FOI]
2. Rainfall (typically the strongest prognostic factor)

The mol[FOI] by a follow-up time ***t*** is defined as the number of new infections molecularly detected between enrollment and time ***t***. For comparisons involving differential calendar periods (e.g., comparisons of Group 5 with any of Groups 2–4 in the ATP cohort), we may use monthly rainfall (in mm), separately for each site, spanning the calendar time during which participants were followed, as reported at http://worldweatheronline.com. Other information accompanying the history of previous malaria infections (timing of infections and clinical malaria cases, parasitemia, dates of malaria infection or clinical malaria, use of antimalarial treatments, and relevant clinical characteristics) may be utilized as available.

Antimalarial treatment is a time-dependent variable of special interest and has the potential to bias estimates of certain target parameters of interest, as described below in subsection 7.1.

### 7 STATISTICAL ANALYSIS FOR PRIMARY OBJECTIVES

#### 7.1 Structure of the Analysis Approach for Assessing Vaccine Efficacy and Relative Vaccine Efficacy

We first consider the following three issues that require attention across the primary and exploratory objectives:

1. Hierarchy of analyses and false-positive control/multiplicity adjustment
2. Adjustment for baseline covariates
3. Antimalarial treatment

For issue 1, as structured in Tables 2, 3, 4, the three sets of comparison types are (1) comparisons of active vaccine study groups vs. rabies control; (2) comparisons of novel active vaccine study groups vs. the standard RTS,S/AS01E regimen; and (3) comparisons of novel active vaccine study groups. Comparisons of Group 4 vs. Group 1 and of Group 5 vs. Group 1 are of primary interest and will not be adjusted for multiplicity in either cohort or for any time period of follow-up.

For all remaining comparisons, the multiplicity adjustment approach for primary objective analyses is as follows: For comparisons of type (1) (remaining comparisons minus the two primary ones), (2) and (3) separately, for each cohort (ATP and TVC) and time-period of follow-up separately, q-values controlling the false discovery rate (calculated using the ***p.adjust*** function in R) and Holm-adjusted p-values controlling the familywise type 1 error rate will be computed along with unadjusted p-values for the set of multiple comparisons. Results are expected to be correlated across cohorts and follow-up periods, such that full FWER adjustment is deemed overly stringent on type I error control at the expense of reduced power.

For issue 2, methods based on targeted maximum likelihood will use Super Learner as an intermediate step, both for predicting the probability of a missing measurement of primary endpoint 2 and for predicting the value of primary endpoint 2, allowing the entire set of pre-specified baseline covariates to potentially be used in predictions (see Section 7.6.1 for definition of an observed/known primary endpoint 2). In contrast, methods not using targeted maximum likelihood (e.g., Cox proportional hazards models) will instead stratify by study site in the study site-pooled analysis and possibly adjust for the “ATP Propensity Score” univariate baseline covariate defined below in Section 7.5 if this propensity score is found to be sufficiently predictive of the outcome under study.

For issue 3, the primary analyses will disregard antimalarial treatment because new malaria infections are observed in the trial within an arbitrary window following drug treatment onset. Nonetheless, because antimalarial treatment may still reduce the risk of a new infection for some time period after treatment onset and because the administration of treatment may be less frequent in vaccine arms if there is protective efficacy, there remains a potential for bias due to differential amount of post-treatment follow-up time characterized by a reduced risk of new infection across study groups. Therefore, we will also conduct a sensitivity analysis in which the primary analyses will be repeated after removing from the follow-up time the period of 14 days following the onset of each antimalarial treatment.

#### 7.2 Descriptive Analysis

Using graphical methods and descriptive statistics, assumptions required for validity of statistical methods and presence of outliers will be assessed. Continuous variables will be summarized by means, standard deviations, interquartile ranges, and ranges. When appropriate, variables will be transformed to improve inferences. Categorical variables will be summarized by proportions.

#### 7.3 Baseline Characteristics

Baseline characteristics of interest will be summarized by treatment groups to ensure balance due to randomization. The number of filter paper samples lost or not collected will also be tabulated by treatment groups and time points, as relevant, as well as PCR negative results or any other reason for excluding a sample from the analysis. In mixed infections, it is expected that sequencing will yield the genotype of the more prevalent strain(s), with sequencing error filters allowing detection of minor genotypes represented by as little as 1% of total reads and a minimum of 50 reads within a sample.

#### 7.4 Handling Missed Vaccinations and Loss to Follow-Up

Analyses in the TVC are intention-to-treat, including all follow-up information after first vaccination regardless of number of vaccinations received, whereas analyses in the ATP cohort only include individuals who qualify for ATP cohort membership. For primary endpoint 1, the event of right-censoring is defined in Section 7.5.1.

#### 7.5 Analyses of Primary Endpoint 1

The first primary endpoint is the time from a treatment-group-specific time origin until the time of the first molecular detection of a new malaria infection within a specified follow-up period.

VE will be assessed using a proportional hazards model, stratifying by study site in the study site-pooled analysis (i.e., allowing a separate baseline hazard rate of new malaria infection detection at each study site). For each vaccine group, VE (referred to as proportional hazards [PH] VE) is estimated as one minus the hazard ratio (vaccine/control vaccine) of the first molecularly confirmed infection detection with 95% Wald-based CI and a Wald test of the null hypothesis that VE equals zero. To minimize bias, it is helpful to control for predictors of molecular detection of new malaria infection (especially for the ATP analysis), and the Cox modeling analysis (implemented with the ***coxph*** function in the ***survival*** R package) will control for the baseline covariates listed in Section 6.2. The same proportional hazards modeling approach will be used for comparing the risk of molecular detection of new malaria infection between pairs of active vaccine arms, except hazard ratios will be assessed instead of VE parameters.

For the ATP analyses, differences in prognostic factors for molecular detection of new malaria infection could arise between the analyzed groups because the analyses are restricted to subgroups defined by the post-randomization variable of qualifying for the ATP cohort at Month 2.5 or Month 7.5 (Horne et al., 2001; Gilbert et al., 2013). In particular, a needed assumption for the ATP cohort analyses to assess causal effects of vaccination is that there are no unmeasured/unaccounted for baseline covariates that predict both ATP qualification status and the outcome type under study. Therefore, in preparation for the proportional hazard analyses, we will first study if there exist measured baseline covariates other than study site and ATP Propensity Score (described below) that are dual predictors of ATP status and either outcome type. If affirmative evidence is uncovered, such predictors will be adjusted for in every ATP cohort analysis, along with study site and ATP Propensity Score (if it is found to be predictive of the ATP status).

The following approach will be used to assess this evidence. Super Learner will be conducted to predict status of qualifying for the ATP from all available baseline covariates, using negative log-likelihood loss and 10-fold cross-validation. A simple parametric library of learners from the ***SuperLearner*** or ***sl3*** R packages will be used, for example, logistic regression that considers all possible two-way interactions from among the baseline covariates listed in Section 6.2 and that selects the best model based on stepwise model selection. If the classification accuracy is clearly greater than expected from random noise, based on the 95% confidence interval for the cross-validated area under the ROC curve exceeding 0.5, then an ATP Propensity Score will be defined as the logit transformed Super Learner model fitted probabilities of being ATP, and all ATP cohort analyses of primary endpoint 1 will adjust for the ATP propensity score. The statistical analyses will use the nonparametric bootstrap for producing confidence intervals and p-values, to account for the fact that the propensity score is estimated. Depending on the number of variables that may be predictive of ATP qualification, we may consider more flexible methods for modeling the ATP propensity score, such as the highly adaptive lasso.

The TVC analyses directly compare hazard rates between vaccine arms not including control group data, as do the ATP analyses that compare vaccine arms within the same vaccination schedule (Month 0, 1, 2 or Month 0, 1, 7). In contrast, ATP analyses comparing vaccine arms with a Month 0, 1, 2 schedule, versus the Group 5 vaccine arm with a Month 0, 1, 7 schedule, include the control vaccine group data in each distinct follow-up period between vaccinations, to control for potential bias due to secular trends, for example caused by seasonality of malaria infection. In particular, the ATP analyses that make an adjustment via the control group are listed in Tables 2, 3 and 4 in Section 5.1.

For example, as in Table 2, the analyses are done using a Month 2.5 to 14 follow-up period for Groups 3 and 4 versus a Month 7.5 to 19 follow-up period for Group 5, creating the same amount of follow-up for malaria post-Dose 3 in the groups being compared.

We implement these analyses using the Cox proportional hazards model as follows: To assess relative VE comparing GP5 with GP2+GP3 in the ATP cohort during 11.5 months following the third dose in each regimen, we will fit (1) a Cox model to GP5 and GP1 data during months 7.5−19 of follow-up, and (2) a separate Cox model to GP2+GP3 and GP1 data during months 2.5−14 of follow-up. We will estimate the hazard ratio comparing GP5 months 7.5−19 with GP2+GP3 months 2.5−14 as the ratio of the estimated GP5-vs-GP1 hazard ratio in model (1) and the estimated GP2+GP3-vs-GP1 hazard ratio in model (2). We will use a nonparametric bootstrap procedure to construct a 95% CI for the GP5-vs-GP2+GP3 hazard ratio, as well as for obtaining a 2-sided p-value for the test of the null hypothesis that the hazard ratio equals one.

A secondary analysis will estimate instantaneous VE over time for each vaccine arm versus the control vaccine using nonparametric kernel smoothing (Gilbert et al., 2002). This analysis will provide pointwise and simultaneous 95% CIs about instantaneous VE over time, together with a p-value for whether instantaneous VE varies with time. Similarly, we will apply this method to estimate hazard ratios over time comparing pairs of vaccine arms. Nonparametric kernel smoothing will also be used to estimate the instantaneous risk of new malaria infection over time for each individual study arm, where this plot for the control vaccine arm is useful for displaying background malaria exposure and infection intensity over time.

For analyses of the TVC, the nonparametric kernel smoothing analysis described above will be applied to each of the pairwise study group contrasts described in Table 4, as well as for each individual study group in Table 4, all for follow-up between Month 0 and 20. For analyses of the ATP cohort, comparisons are done post-Dose 3 through 20 months, not including comparisons of Group 5 with Groups 3 and 4 because of the different times of dose 3. Specifically, Table 5 describes the ATP comparisons that are made, and the individual study group analyses will also be done. If for some of the study groups a substantial number of participants did not experience primary endpoint 1 by 20 months (operationally defined as at least 25% for at least one study group), then the nonparametric kernel smoothing analyses are repeated for follow-up through Month 32 (exactly as in Table 5 with M20 replaced with M32).

Table 5. Study Groups Compared and Individual Groups Analyzed by Nonparametric Kernel Smoothing of the Instantaneous Hazard Ratio (Primary Endpoint 1)

| Analysis Type | Study Group and Follow-up Comparison |
|---|---|
| Assess Vaccine Efficacy (ATP) | GP2 M2.5-20 vs. GP1 M2.5-20 |
| | GP3 M2.5-20 vs. GP1 M2.5-20 |
| | GP4 M2.5-20 vs. GP1 M2.5-20 |
| | GP5 M7.5-20 vs. GP1 M7.5-20 |
| Assess Relative Vaccine Efficacy vs. Standard regimen R012-20 (ATP) | GP3 M2.5-20 vs. GP2 2.5-20 |
| | GP4 M2.5-20 vs. GP2 2.5-20 |
| Assess Relative Vaccine Efficacy (Other Comparisons) (ATP) | GP4 M2.5-20 vs. GP3 M2.5-20 |

An additional secondary analysis will estimate cumulative distributions of the time to the first molecular detection of a new malaria infection by study group. The cumulative distribution functions will be estimated in two ways: first, by using the Nelson-Aalen estimator for the cumulative hazard function, and second, by adjusting for other baseline participant characteristics via targeted maximum likelihood estimation (TMLE, Moore and van der Laan, 2009). The latter fully covariate-adjusted method is especially relevant for the ATP analysis, as described above. This analysis uses as an intermediate step an estimate of the distribution of the time to the first molecular detection of a new malaria infection conditional on baseline covariates (listed in Section 6.2), which is produced using the Super Learner method (as implemented in R packages *SuperLearner* or *sl3*). Given the possibility of our requiring resampling methods (e.g., nonparametric bootstrapping) to obtain inference, we may restrict the Super Learner library of candidate regression methods to those that are computationally stable and run relatively quickly. For example, these candidate regressions will include regressions based on several different generalized linear models that include up to two-way interactions between covariates and treatment.

The estimated cumulative distribution functions of the time to the first molecular detection of a new malaria infection will be plotted with 95% pointwise and simultaneous confidence intervals, through to the last time point in the specified time period. Similarly, point estimates and 95% confidence intervals will be produced for additive differences and cumulative incidence-based vaccine efficacy or cumulative relative risk contrasts between the two study groups being compared. These analyses will be done for each individual and pair of study groups specified in Table 5, analogously to the analysis of instantaneous kernel-smoothed hazard functions and their contrasts.

##### 7.5.1 Handling of Loss to Follow-up for Primary Endpoint 1

*Cox proportional hazards models and nonparametric inference on instantaneous hazard ratios over time*

Regarding loss to follow-up, for both TVC and ATP cohort analyses, for each participant included in the analysis, the right-censoring time is defined as the last parasite negative sample collection date recorded during the follow-up period under study. The assumption of random censoring is needed for valid inferences.

The time to the first molecularly confirmed new malaria infection detection is right-censored. The analyses will be conducted using a standard right-censored failure time set-up, with failure time defined as the date of the first molecular detection of a new malaria infection.

*TMLE inference on cumulative incidence curves and study group contrasts in cumulative incidence curves*

Right-censoring/loss to follow-up is defined as for the proportional hazards analyses. The assumptions of random censoring after conditioning on measured baseline covariates and consistent estimation of the conditional distribution of the censoring time given these covariates is needed to assure valid inferences. Because the TMLE method is doubly-robust, it is valid even if the conditional censoring distribution is modeled inconsistently, as long as the conditional outcome regression is estimated consistently. The analysis will be done using the same time-to-event variable used for the Cox proportional hazards analysis.

#### 7.6 Analyses of Primary Endpoint 2

Primary endpoint 2 is the number of new molecularly confirmed malaria infections during a specified follow-up period (see Section 7.6.1 for definition of an observed/known primary endpoint 2).

Plots of estimates of reverse cumulative distribution functions (rcdfs) of the number of new molecularly confirmed malaria infections by treatment arm occurring within each specified follow-up period will be used to summarize the distributions of primary endpoint 2. These plots show the proportion of participants with more than $n$ new malaria infections on the y-axis versus $n$ on the x-axis, and allow visual assessment of how specified quantiles differ over study groups (e.g., medians, 25th percentiles, 75th percentiles). The rcdfs will be estimated by targeted maximum likelihood estimation that adjusts for the baseline covariates described in Section 6.2. Corresponding to these plots, the distribution of the total number of new malaria infections over specified time periods will be tabulated by study group. These analyses are done for each study group and follow-up period specified in Tables 2–4.

Vaccine efficacy against primary endpoint 2 for a given vaccine arm vs. the control arm will be assessed by the additive difference in the mean number of new molecularly confirmed malaria infections in each vaccine arm vs. the mean number of new molecularly confirmed malaria infections in the control vaccine arm. The targeted maximum likelihood estimator of Porter et al. (2011) will be used, which incorporates information in baseline study characteristics predictive of whether data are missing or predictive of the number of new molecularly confirmed malaria infections. This method also provides a way to estimate the variance of the estimated mean number of new molecularly confirmed malaria infections in each study arm. Based on these variance estimates, Wald tests will be used to test for the additive difference VE differing from zero and to obtain a 95% CI. These analyses are done for each comparison specified in Tables 2–4 with the control group as the comparison arm.

Similarly, the same methods will be used to test for differences in the mean numbers of new molecularly confirmed malaria infections between vaccine arms, and to compute 95% CI for these differences. Again Wald 95% CIs and Wald 2-sided p-values are used. These analyses are done for each comparison specified in Tables 2–4.

##### 7.6.1 Handling of Loss to Follow-up and Missing Visit Data for Primary Endpoint 2

The number of new molecularly confirmed malaria infections must be estimated based on the genotyping/haplotyping data, and we use an operational definition for what data availability constitutes enough information for the study endpoint to be considered observed/known for an individual. A participant's endpoint is considered to be observed/known if both (a) s/he had at least one visit with genotyping data in the 75-day period leading up to the last scheduled visit in the given follow-up period (e.g., Month 0 to 20, Month 2.5 to 14, Month 7.5 to 19), and (b) s/he had genotyping data from at least 50% of scheduled visits during the given follow-up period. If the last scheduled visit in the given follow-up period was missed, then the 75-day period is keyed off of the target date of the missed visit. The premise for this estimator is that, with these available data, it will be possible to accurately estimate the number of new molecularly confirmed malaria infections during the follow-up period.

*TMLE inference about reverse cumulative distribution functions (RCDFs)*
The TMLE of the RCDF incorporates both a Super Learner estimate of the probability of missing the outcome conditional on baseline covariates and a Super Learner estimate of the outcome itself conditional on baseline covariates (again done with the *SuperLearner* or *sl3* R packages with default learners and baseline covariates described in Section 6.2). The method is well-suited to the application, because some participants who do not have an observed primary endpoint 2 due to not meeting condition (a) or (b) in the paragraph above will still have useful data (e.g., genotyping

results on, say, 40% of samples) for predicting primary endpoint 2, and any prediction ability even if quite imperfect improves efficiency of the analysis. More specifically, we view the objective of inference about the mean of primary endpoint 2 as a problem to address using two phases of data—phase one data measured in all participants and the phase two data the primary endpoint 2 ($Y$) that is missing for some participants. The phase one data include baseline characteristics of participants as well as auxiliary outcomes that may be predictive of the outcome $Y$ of interest. Accounting for these phase one data, including information from participants with $Y$ missing, can improve robustness and efficiency. We select as a phase one auxiliary outcome of interest (call it $Y*$) the number of molecularly detected new infections divided by the number of available molecular detection tests during the time period of study for a given analysis. If a participant has zero in the denominator, then $Y*$ is set to 0. Moreover, the method is doubly-robust such that it gives valid inferences even if the Super Learner model for this prediction is inconsistent, as long as the probability of missing the outcome conditional on baseline covariates is consistently estimated. Thus, importantly, a completely accurate prediction is not required for valid inference, a property that has been shown theoretically and in simulation studies in many TMLE papers.

TMLE inference on mean number of new malaria infections and study group contrasts will be reported. Missing data for this TMLE are handled in the same way as described above for the TMLE of the RCDF.

#### 7.7 Assessment and Comparison of Vaccine Efficacy after the Month 14 or 20 Boost through to Month 32

A booster dose is given at Month 14 for two of the vaccine arms, Groups 3 and 4 (with later boosts at Month 26 and Month 38), and at Month 20 for two of the vaccine arms, Groups 2 and 5 (one with no later boost and one with a later boost at Month 32).

All analyses using follow-up through Month 32 will use the modified post-hoc definition of molecular parasite positivity as described in Section 5.1.1.1.

##### 7.7.1 Analysis of Primary Endpoint 1

The TVC analysis of primary endpoint 1 assesses time from first vaccination until the first new malaria infection through to Month 32, using the same methods as described above (see Table 4). This analysis may provide little information beyond the analyses described above based on data collected through Month 20 given that most participants are expected to be infected with malaria by Month 20. The ATP cohort analysis of primary endpoint 1 through 24 months post-Dose 3 will likely be more informative. This ATP cohort analysis uses the methods described above, analyzing the study groups and follow-up periods described in Table 4, and with baseline covariates to adjust for described above in Section 6.2.

##### 7.7.2 Analysis of Primary Endpoint 2

The statistical methods are those described above for primary endpoint 2, except that different follow-up intervals are used for counting the numbers of new infections. ATP analyses will be conducted using two follow-up periods: 7 and 24 months of follow-up post-Dose 3. Additionally, TVC analyses will be conducted also using two follow-up periods: 0-14 and 0-32 months. Obtaining results over the three segments—7, 14, and 24 months post-Dose 3 in the ATP analyses, and 7, 20, and 32 months post-first-vaccination in the TVC analyses—will inform about how vaccine efficacy on primary endpoint 2 varies with follow-up segment. The study group comparisons and follow-up periods are described in Tables 2–4.

#### 7.8 Significance Level

In primary analyses comparing Group 4 vs. Group 1 and Group 5 vs. Group 1 in either study cohort and for any time period of follow-up, statistical significance is defined as an unadjusted p-value ≤ 0.05. In all remaining primary analyses, multiple hypothesis testing adjustment is used, as described in Section 7.1., to evaluate statistical significance. Within each multiple comparison procedure, statistical significance is defined as a q-value ≤ 0.2 or a Holm-Bonferroni-adjusted p-value ≤ 0.05, together with an unadjusted p-value ≤ 0.05. All statistical tests in support of the primary objectives are two-sided.

#### 7.9 Interim Analyses

An interim genetic analysis will occur on data collected up to Month 21 as per the main MALARIA-094 protocol. A second analysis will be performed on data collected up to Month 32. These analyses will be triggered by the last enrolled participant reaching Month 21 or Month 32, respectively. A study report presenting all results accounting for study follow-up through to Month 32 will be produced.

All genetic material and sequence analyses will be performed through the study periods following a schedule agreed-upon by GSK and as specified in this SAP. At the interim analyses, the TVC and ATP cohort analyses will be performed using data through Month 20, with comparisons and follow-up periods defined in Tables 1, 2, and 3. At the final analysis, the TVC and ATP cohort analyses will also analyze new malaria infection data through Month 32, with comparisons and follow-up periods defined in Table 4.

#### 7.10 Procedures for Preventing Biases and Accounting for Missing, Unused, or Spurious Data

Bias in genotyping analyses will be prevented by the sample collection protocol previously described and procedures specified in the main trial protocol MALARIA-094 that will de-identify subject samples. Comparisons of relative diversity between vaccinated and control vaccinated arms will be made at the level of subject study sites when possible. In all cases, adjustments by study sites will be performed. To prevent differential misclassification bias, sequence analysis is performed blinded with respect to knowledge about the vaccination status connected to an isolate and to the specific malaria outcome (i.e., asymptomatic infection or clinical malaria).

The genotyping and re-sequencing platforms at the Broad Institute implement strict quality control guidelines to reduce the likelihood of sample contamination. Appropriate and rigorous data cleaning for all single nucleotide polymorphisms (SNPs) will be performed. Unreliable sequencing or genotyping data will be excluded from the analysis, and reasons for excluding data included in the data base when available. All unused or discarded samples will be noted and reported as described in the main trial protocol MALARIA-094.

### 8 STATISTICAL ANALYSIS FOR EXPLORATORY OBJECTIVES

#### 8.1 Overview of Exploratory Objectives

The exploratory objectives are as follows:

> **Exploratory objective 1:** To assess whether VE against primary endpoint 1 and 2 depends on genotypic characteristics of the malaria parasites, based on longitudinal monthly and other unscheduled sampling.
>
> **Exploratory objective 2:** To study whether prior infection or vaccination has a relationship to subsequent infection by measuring the molecular Force of Infection (mol[FOI]) determining the relationship between (mol)FOI and subsequent malaria infection risk.
>
> **Exploratory objective 3:** To determine whether prior infection by a particular parasite genotype, not necessarily the vaccine strain, reduces the likelihood of re-infection by a parasite with the same genotype.

Given that the exploratory analyses must deal with the diversity of malaria infections, we first describe a section of how we plan to characterize AA sequence features and within-infection malaria diversity. Subsequent sections describe how the analyses to meet the exploratory objectives account for that characterization.

#### 8.2 Structuring AA Sequence Features for Sieve Analysis (Exploratory Objective 1)

The exploratory objective 1 involves evaluating whether and how the various efficacy and relative efficacy comparisons, described in Table 2 plus the additional comparison of active arms GP2–GP4 pooled vs. the control arm, for primary endpoints 1 and 2 depend on different AA sequence features pre-specified in a treatment-blinded manner. All features will be calculated using aligned translated AA sequences of the CS C-terminus and SERA2 amplicons, obtained after application of the standardized error filtration process (Early et al., 2019) to the underlying nucleotide sequence data measured on an Illumina platform. We will consider two categories of features:

(i) features that are characteristics at the haplotype level, and
(ii) features that are characteristics at the sample level.

Category (i) constitutes the same set of features that were analyzed in the Phase 3 trial sieve analysis and published in Neafsey et al. (2015) and Benkeser, Juraska and Gilbert (2020). These features are as follows:

a) AA residue match vs. mismatch to the 3D7 vaccine strain in the full CS C-terminus amplicon;
b) AA residue match vs. mismatch to the 3D7 vaccine strain in each of the following four haplotypic regions in the CS C-terminus: Th2R, Th3R, DV10, and the LD haplotype (a union of specified positions in Th2R and Th3R in linkage disequilibrium);
c) AA residue match vs. mismatch to the 3D7 vaccine strain at individual polymorphic AA positions in the CS C-terminus;
d) Hamming distance to the 3D7 vaccine strain based on the full CS C-terminus amplicon as well as in each of the haplotypic regions in (b).

In addition, sieve analyses for (a), (c), and the full amplicon in (d) will be repeated for the SERA2 amplicon. This analysis serves as a control for the analysis of the CS C-terminus amplicon, given that SERA2 is not included in the vaccine, and hence we assume that there should be no sieve effects in SERA2.

The novelty in the analysis of the same features as in the Phase 3 trial sieve analysis is twofold: it will characterize sieve effects pertaining to the fractional dose regimens, and it will characterize feature-specific efficacy and relative efficacy against primary endpoints 1 and 2.

Features in category (ii) aim to describe within-host ***P. falciparum*** population complexity using multiple different sample-level complexity indices to capture different aspects of the proteomic diversity and to subsequently study variations in vaccine efficacy and relative efficacy with the selected complexity measures. With the exception of COI, all other complexity indices disregarding the vaccine strain will be calculated separately for the CS C-terminus and SERA2 loci as the sequencing technology does not provide requisite data to concatenate sequences from the two amplicons. The indices, specified below, were down-selected from a large pool of indices described in Gregori et al. (2016). The down-selection was based on a combination of the authors' recommendations and results from an exploratory study, performed by the Fred Hutch statistics group, of statistical properties of all Gregori et al. indices and their inter-relatedness using CS C-terminus amplicon AA sequence data from clinical malaria and Month 20 cross-sectional parasite positivity cases among participants aged 5-17 months in the MAL-066 trial that were also analyzed in Neafsey et al. (2015).

The following complexity indices (i.e., estimators) in category (ii) will be calculated in each sample, separately for the CS C-terminus and SERA2 amplicon:

e) Hill numbers (Hill, 1973), H(q), of orders q = 0, 1, and 2, where H(0) is the number of distinct haplotypes observed in the sample, H(1) is undefined but its limit as q tends to 1 is the exponential of Shannon entropy, and H(2) is the inverse of the Simpson index defined as the probability that two sequence reads drawn at random in a given sample belong to the same haplotype;

f) the standardized number of segregating AA positions, $S/a_1$, where $S$ is the number of segregating positions, and $a_1 = \sum_{i=1}^{n-1} 1/i$, where $n$ is the number of reads in a given sample; and

g) the mean Hamming distance to the 3D7 strain with equally weighted haplotypes.

In addition to (e)-(g), we will analyze variation in efficacy and relative efficacy by

h) COI,

which pools information from both amplicons and equals the maximum of the Hill numbers H(0) for CS C-terminus and SERA2.

Indices (e), (f) and (h) disregard the 3D7 reference sequence, whereas index (g) accounts for the 3D7 sequence (which, for CS C-terminus, was the vaccine insert). The mean pairwise Hamming distance is not considered for analysis because we anticipate that a large portion of the first new infections will have COI = 1, and therefore zero mean pairwise Hamming distance. Indices (e)-(g) will be calculated using the Fred Hutch's internal R package ***divIndex***.

For analyses in pursuit of the exploratory objective 1, for each AA sequence feature in both categories (i) and (ii), we will calculate the value(s) of the feature in the sample associated with the first molecular detection of a new infection (i.e., primary endpoint 1) as well as pooling all samples associated with all molecular detections of a new infection in a given participant within the studied follow-up period, unless data are missing for these calculations. For category (i) features, a value will be calculated, if possible, for each haplotype. For each category (ii) feature, we will generate scatter-box-violin plots of the observed values of the feature by study group. Furthermore, we will generate Hill number profiles for q in [0, ∞] by study group, where, for a given q, each study group-specific curve will show the mean Hill number across all samples in this group associated with the first new infection.

We expect that 20% or more of primary endpoint 1 cases will have missing haplotype data for one amplicon. This is a result of the fact that measuring at least 1 haplotype (with ≥50 reads) for one amplicon and none for the other is sufficient to register the event as primary endpoint 1. We will use missing-data versions of sieve analysis methods to account for the missing haplotype data.

#### 8.3 Pre-screening of CS C-terminus and SERA2 Sequence Features

To maximize statistical power to detect sieve effects, a pre-specified screening process is used to select a subset of the features listed in Section 8.2 to include in sieve analyses addressing exploratory objective 1. The screening procedure, described below, is performed separately for each comparison, pooling over all treatment arms included in the comparison (i.e., blinded to individual treatments included in the comparison). However, to identify the treatment arms included in the comparison, the treatment assignments must be available. The treatment comparisons considered for exploratory objective 1 are listed in Table 2, in addition to the comparison of active arms GP2–GP4 pooled vs. the control arm. The screening procedure will be conducted separately for each study site-specific analysis and for the study site-pooled analysis.

For the haplotype-specific AA sequence features (a)–(c) that are categorical, a screening process is needed to determine which features have a large enough representation in the data set, and enough variability, to make possible sieve analysis. A subset of the features in (a)–(c) will be selected for sieve analysis, defined as follows: Two screened-in feature sets are defined, one for the analysis of the first new infection, and one for longitudinal data analysis including new haplotypes (i.e., not observed in the participant's previous new infections) from all new infections in the ATP cohort with follow-up through 12 months post-Dose 3 (see Section 8.4.1 for the sieve analysis cohort and follow-up period), relevant for primary endpoint 2 of this objective and for exploratory objective 3. We first describe the former screen. For each feature (i.e., a 3D7 match/mismatch in a specific haplotypic region, or an AA residue 3D7 match/mismatch at a specific AA position), the frequencies of the possible values of each feature (0 or 1 for a match/mismatch feature) among all first new infection endpoint cases (pooling over all treatment arms included in a comparison) will be tabulated (number of primary endpoint 1 cases with a given feature value / number of primary endpoint 1 cases). Feature values with representation from at least 30 participants, pooling over all treatment arms included in a comparison, will be screened-in for data analysis of vaccine efficacy or relative vaccine efficacy against primary endpoint 1 with the given feature value. In addition, only features for which there is enough representation from at least 30 participants for each level of the feature (e.g., for both 3D7 match and mismatch) will be screened-in for sieve analysis of differential vaccine efficacy or relative vaccine efficacy across different feature levels associated with the primary endpoint 1 sample. The number 30 may be altered based on this descriptive analysis, which is valid because the screening procedure itself is blinded to randomization arm (albeit the selection of arms in a comparison is unblinded).

Similarly, for longitudinal data analysis including all new infections during M0-20, the following screen is used: For each feature in (a)–(c), the frequencies of the possible values of each feature among all new infection endpoint cases (pooling over all treatment arms included in a comparison) will be tabulated (number of primary endpoint 2 cases with a given feature value / number of primary endpoint 2 cases). Parallel to the above, feature values with representation from at least 50 participants, pooling over all treatment arms included in a comparison, will be screened-in for data analysis of vaccine efficacy against primary endpoint 2 with the given feature value, and only features for which there is enough representation from at least 50 participants for each level of the feature will be screened-in for primary endpoint 2 exploratory objective 1 and 3 sieve analyses. As above, the number 50 may be altered based on this descriptive analysis, which is valid because the screening procedure itself is blinded to randomization arm (albeit the selection of arms in a comparison is unblinded).

#### 8.4 Analysis of Exploratory Objective 1

Exploratory objective 1 is approached in two ways: first, for primary endpoint 1, and, second, for primary endpoint 2, for which the issue of COI is treated differently. For primary endpoint 1, it is expected that the first new malaria infection will usually have COI = 1 (under the premise that most new infections are with a single haplotype), or occasionally COI = 2 or 3 if co-infection from the same mosquito sometimes occurs or multiple mosquito transmission events occur between longitudinal monthly blood samples. Because COI is expected to be low for primary endpoint 1, for haplotype-specific AA sequence features (a)–(d), sieve analysis methods that assume only one haplotype will be used, and exhaustive or Monte Carlo multiple outputation applied, similar to what was done in Neafsey et al. (2015). For sample-specific sequence features (e)–(h), the employed sieve analysis methods will not require to be augmented with multiple outputation.

In contrast, for primary endpoint 2, COI by the end of a given follow-up period may be quite large and is essentially the same concept as primary endpoint 2 itself in being closely related to the number of new malaria infections by a given time point. Therefore sieve analyses of primary endpoint 2 treat COI as a key issue, and the "active surveillance" versions of the Follmann and Huang (2018) methods are one set of methods that apply, where the "terminal event" is defined as the last new malaria infection event during the specified follow-up period.

The following paragraphs describe the exploratory statistical analyses that are planned of the AA sequence features described above.

##### 8.4.1 Cohort and Follow-up Periods for Sieve Analysis

All sieve analyses will be conducted in the ATP cohort with (i) follow-up through 12 months post-Dose 3, and (ii) follow-up through 24 months post-Dose 3 (i.e., the same cohort and follow-up periods as in the primary analyses). Both study site-specific and study site-pooled analyses will be performed.

##### 8.4.2 Sieve Analysis for Primary Endpoint 1

The methods assess genotype-specific (relative) vaccine efficacy and evaluate differential (relative) vaccine efficacy (i.e., the so-called sieve effect) against primary endpoint 1.

The following sieve analysis methods will be applied that assume a single haplotype, observed or missing, per case (with multiple outputation added for haplotype-specific sequence features (a)–(d)):

For the binary haplotype-level features in (a)–(c), we will estimate genotype-specific hazard-ratio (relative) VE with 95% CIs and a two-sided test for differential genotype-specific (relative) VE using the methods proposed in the yet unpublished manuscript Heng, Sun, and Gilbert, “Estimation and hypothesis testing of strain-specific vaccine efficacy with missing strain types with applications to a COVID-19 vaccine trial” and implemented in the R package ***cmprskPH***. This method accommodates missing sequences by augmented IPW.

For the Hamming distance feature in (d), the genotype-specific hazard-ratio (relative) VE, defined as one minus the genotype-specific hazard ratio (vaccine/control), will be assessed in the ATP cohort using the genotype-specific hazard-ratio model for either fully observed genotypes (Juraska and Gilbert, 2013; implemented in the R package *sievePH*) or for partially missing genotypes using inverse-probability weighting (IPW) or augmented IPW (AIPW) (Juraska and Gilbert, 2016; implemented in *sievePH*), stratified by study site (for the study site-pooled analysis only) and adjusted for predictors of new malaria infection. We will use the IPW or AIPW version of the analysis method for amplicon-specific genotypes that are continuous or count variables and the standard version for COI which is an aggregate measure across both amplicons. For each ATP cohort analysis in Table 2 comparing vaccine to control, plus comparing vaccine arms GP2–GP4 pooled to control, inference will include 95% pointwise Wald CIs for genotype-specific VE and a Wald test of the null hypothesis that VE is invariant with respect to the AA sequence feature. For each ATP cohort analysis in Table 2 comparing two vaccine regimens head-to-head, the same inference will be reported except on the hazard ratio scale.

One-sided testing will be performed for comparisons to the control GP1 regimen of 3D7 Hamming distances, and two-sided testing will be performed for head-to-head RTS,S regimen comparisons of 3D7 Hamming distances as well as for all comparisons of all other quantitative sequence features. Statistical significance is defined as in Section 7.8 except, for multiplicity sets in which one-sided testing is performed, the same definition of FWER and FDR statistical significance is applied to doubled one-sided p-values.

We will also apply the cumulative incidence-based sieve analysis methodology of Benkeser, Juraska, and Gilbert (2020), extended to accommodate missing sequence data under a missing at random assumption. These methods treat the Hamming distance of an infecting parasite to the 3D7 vaccine strain as a count variable. For applications of the method where the distance is continuous (or nearly so) rather that a count variable, the Benkeser, Juraska, Gilbert et al. methodology will be applied binning the continuous values onto a grid.

##### 8.4.3 Multiple Hypothesis Testing Adjustment for Sieve Analysis for Primary Endpoint 1

To minimize the occurrence of false positive sieve effect results given the large number of analyzed sequence features and group comparisons and thereby increase the credibility of the sieve analysis, it is critical to use a stringent multiplicity adjustment for hazard-based sieve tests. To this end, q-values controlling the false discovery rate and

Holm-Bonferroni-adjusted p-values controlling the familywise type 1 error rate will be used, applied in the analyses of follow-up data through the Month 20 visit separately to the following multiplicity sets defined by combinations of a group comparison with a set of features:

- GP2–GP4 vs. GP1 by 3D7 residue match/mismatch features in all screened-in CS C-terminus haplotypic regions and CS C-terminus AA positions
- GP2+GP3 vs. GP1 by 3D7 residue match/mismatch features in all screened-in CS C-terminus haplotypic regions and CS C-terminus AA positions
- GP4 vs. GP1 by 3D7 residue match/mismatch features in all screened-in CS C-terminus haplotypic regions and CS C-terminus AA positions
- GP5 vs. GP1 by 3D7 residue match/mismatch features in all screened-in CS C-terminus haplotypic regions and CS C-terminus AA positions
- GP4 vs. GP2+GP3 by 3D7 residue match/mismatch features in all screened-in CS C-terminus haplotypic regions and CS C-terminus AA positions

- GP2–GP4 vs. GP1 by 3D7 Hamming distance in all CS C-terminus haplotypic regions
- GP2+GP3 vs. GP1 by 3D7 Hamming distance in all CS C-terminus haplotypic regions
- GP4 vs. GP1 by 3D7 Hamming distance in all CS C-terminus haplotypic regions
- GP5 vs. GP1 by 3D7 Hamming distance in all CS C-terminus haplotypic regions
- GP4 vs. GP2+GP3 by 3D7 Hamming distance in all CS C-terminus haplotypic regions

- GP2–GP4 vs. GP1 by 3D7 residue match/mismatch features in all screened-in SERA2 AA positions
- GP2+GP3 vs. GP1 by 3D7 residue match/mismatch features in all screened-in SERA2 AA positions
- GP4 vs. GP1 by 3D7 residue match/mismatch features in all screened-in SERA2 AA positions
- GP5 vs. GP1 by 3D7 residue match/mismatch features in all screened-in SERA2 AA positions
- GP4 vs. GP2+GP3 by 3D7 residue match/mismatch features in all screened-in SERA2 AA positions

Multiplicity adjustment will be applied separately in the study site-pooled and each study site-specific analysis. In the analyses of data through the Month 32 visit, comparisons involving GP2 and GP3 will be performed separately, i.e., the full set of group comparisons will be: GP2–GP4 vs. GP1, GP2 vs. GP1, GP3 vs. GP1, GP4 vs. GP1, GP5 vs. GP1, GP3 vs. GP2, GP4 vs. GP2, GP4 vs. GP3.

No multiplicity adjustment will be applied to COI and the other complexity indices.

Statistical significance is defined the same as in Section 7.8 with one-sided p-values doubled before multiplicity adjustment is performed.

##### 8.4.4 Sieve Analysis for Primary Endpoint 2

Accounting for all data on new malaria infections collected during a given specified follow-up period, the three "active surveillance" versions of the Follmann and Huang (2018) methods specified in the last two columns of Table 3 of their paper will be applied to address different questions, as explained in Follmann and Huang. Here we specify the details of how the variables and models are defined to assess genotype-specific vaccine efficacy and genotype-specific relative vaccine efficacy, for each of the genotype features defined above.

The two Follmann and Huang methods that are applied are the "product method on $X_f$" and the "product method on $I(X_f > 0)$" method. Here $X_f$ for a given participant denotes $X_f = X_{1f} + \ldots + X_{Cf}$, where C is the number of new malaria infections (i.e., primary endpoint 2) and $X_{cf}$ is the number of sequences from the $c^{th}$ new malaria infection that have haplotype f. Both methods are applied assuming model (5) in Follmann and Huang, where the regression coefficient β is estimated using standard Cox model software, which is fit adjusting for the same baseline covariates adjusted for in the primary analyses of primary endpoint 2, and also adjusting for study site for the analyses that pool over study sites. In addition to assuming model (5), the product method on $X_f$ also assumes model (6) of Follmann and Huang, and yields point and 95% CI estimates of $VE_{Mf}$ defined in Follmann and Huang. The product method on $I(X_f > 0)$ assumes model (7) of Follmann and Huang (in addition to model (5)) and yields point and 95% CI estimates of $VE_{If}$ defined in Follmann and Huang. For inferences about differential VE by two haplotypes f=1 vs. f=2, the methods also provide point and 95% CI estimates for Follmann and Huang's differential VE parameters: $\exp(\alpha_{2M}) = (1 - VE_{M1})/(1-VE_{M2})$ and $\exp(\alpha_{2I}) = (1 - VE_{I1})/(1-VE_{I2})$, as well as providing Wald 2-sided p-values for sieve effects, where the product method on $X_f$ tests H0: $\alpha_{2M}=0$ and the product method on $I(X_f > 0)$ testing H0: $\alpha_{2I}=0$.

As for other analyses, the analyses are done for each study site separately as well as pooling the two study sites.

#### 8.5 Analytic Approach for Exploratory Objective 2

##### 8.5.1 Summary of Modeling Approach for Exploratory Objectives 2 and 3

A flexible dynamic model of multiple types of recurrent events will be applied to address exploratory objectives 2 and 3. To express the model, let $T_{i,k,j}$ be the $j$th type-$k$ event of participant $i$, $0 \leq T_{i,k,1} < T_{i,k,2} < \cdots < T_{i,k,n_{ik}} \leq \tau$, where $n_{ik}$ is the total number of type-$k$ events for participant $i$. Let $N_{ik}^*(t)$ be the counting process registering the number of type-$k$ recurrent events for individual $i$ in the time interval $[0, t]$, for $k = 1, \ldots, K$ and $i = 1, \ldots, n$. Let $Y_i(t) = I(C_i \geq t)$ be the at-risk process and $N_{ik}(t) = \int_0^t Y_i(s) dN_{ik}^*(s)$ be the process registering numbers of observed type-$k$ events, where $C_i$ is the right censoring time. Let $\mathcal{F}_t$ represent the event history for all participants up to time $t$. The intensity of $N_{ik}(t)$ is given by $P$.

We consider the following dynamic semiparametric model for the intensity:

$$\lambda_{ik}(t) = \lambda_{k0}(t)e^{\beta_k^T(t)X_i(t)+\gamma_k^T(U_i(t))Z_i(t)}$$

for $k = 1, \ldots, K$, where $\lambda_{k0}(t)$ is an unspecified baseline function, and $\beta_k(\cdot)$ is a vector of unspecified functions that represents possibly time-varying effects of covariates $X_i(t)$, and $\gamma_k(\cdot)$ is a vector of unspecified functions that represents effects of covariates $Z_i(t)$ that can depend dynamically on exposure variables $U_i(t)$ over time such as past malaria infections. The covariates $X_i(t)$ of interest include baseline covariates to adjust for and treatment assignment to RTS,S vaccination vs. control.

The coefficient function $\beta_k(t)$ is the log relative risk corresponding to the increase in the intensity of having a new malaria infection of type k at time t for every one unit increase in the covariate $X_i(t)$. The coefficient function and $\gamma_k(u)$ is the log relative risk corresponding to the increase in the intensity of having a new malaria infection of type k at time t and u unit time since the last new infection or vaccination for every one unit increase in the covariate $Z_i(t)$.

Setting $Z_i(t) = 1$ specifies an analysis of interest, in which case and $\gamma_k(u)$ is the log-scale increase in the intensity of having a new malaria infection of type k at time t and u unit time since the last new infection or vaccination. When and $\gamma_k(u)$ is positive, exp($\gamma_k(u)$) exceeds one and corresponds to a relative intensity (instantaneous relative risk) increasing with time since the last new infection or vaccination.

We can consider $U_i(t) = (U_{i1}^T(t), \ldots, U_{iK}^T(t))^T$ as the part of the multi-type event history that could affect the intensity $\lambda_{ik}(t)$, where $U_{ik}(t)$ is related to the type-k event history of subject $i$. For example, choosing $U_{ik}(t) = (N_{ik}(t^-), t - T_{i,k,N_{ik}(t^-)})^T$ specifies the number of new infections molecularly detected between enrollment and time $t$ (i.e., $N_{ik}(t^-)$ is the molecular force of infection mol[FOI] of strain k by time t for subject i) and the time since the last occurrence of molecularly detected strain $k$ malaria infection, respectively. Specifying $\gamma_k(U_i(t)) = \eta_{kk}(t - T_{i,k,N_{ik}(t^-)})$ models time-varying effects since the last strain $k$ infection, and specifying $\gamma_k(U_i(t)) = \eta_{kj}\left(t - T_{i,k,N_{ij}(t^-)}\right)$ models how the time since the last strain $j$ infection impacts time-varying effects on the intensity of strain $k$ infection. In this notation $\eta_{kj}\left(t - T_{i,k,N_{ij}(t^-)}\right)$ is a nonparametric function of $\left(t - T_{i,k,N_{ij}(t^-)}\right)$.

The new infection event time variable is the same as used for the primary analyses, and the right-censoring time variable is modified. Specifically, the right-censoring time is defined as the last observed visit time (scheduled or unscheduled) before three consecutive missed scheduled visits with no intervening unscheduled visits. We selected three consecutive visits to define censoring based on a treatment-arm pooled descriptive analysis comparing several potential right-censoring definitions based on 2, 3, 4, 5, or 6 consecutive missed visits, where the percentage right-censoring was

45.3%, 29.5%, and 25.4% under the 2, 3, and 4 consecutive missed visits definitions of right-censoring, respectively.

To handle intermittent missed visits prior to right-censoring, we take advantage of the fact that the method flexibly allows subject-specific visit schedules.

Sensitivity analyses may be done that consider a new definition for the new infection event time variable (in terms of the sequence read-count criterion for defining a new malaria infection); if this is done the same new variable studied as a sensitivity analysis to the primary analysis will be studied.

##### 8.5.2 Application of the modeling approach to address Exploratory Objective 2

Analyses for exploratory objective 2 are done in the TVC cohort only and studying new molecularly detected infection events during the M0-20 and M0-32 follow-up periods. Exploratory objective 2 applies the above-specified model counting all new malaria infection endpoints regardless of type k: thus the model is applied with a single k=1. We will study how does all-strain malaria infection risk depend on the molecular force of infection $N_i(t^-)$ and on time since most recent infection, as well as studying whether the effect of the molecular force of infection is modified by prior RTS,S vaccination.

Analysis 1. The model is first applied using the control arm participants only and with $X_i(t)$ consisting of (1) the baseline covariates study site (Siaya or Agogo), age at enrollment, and sex, and (2) the time-dependent covariate $N_i(t^-)$, with $\beta_1(t)$ and $\beta_2(t)$ time-dependent effects. In addition, the model specifies $\gamma(U_i(t)) = h(t - T_{i,N_i(t^-)})$. Estimation of the coefficient $\beta_2(t)$ in front of the $N_i(t^-)$ covariate assesses whether and how much the molecular force of infection up until time t impacts the intensity of new infection, after covariate adjustment. Estimation of the coefficient function $h(t - T_{i,N_i(t^-)})$ assesses how the time since the last new malaria infection impacts the intensity of new malaria infection, after covariate adjustment.

Analysis 2. The above analysis is repeated for all of the RTS,S vaccine arms pooled together.

Analysis 3. Analysis 3 repeats Analysis 2, again restricting to the RTS,S vaccine arms pooled, except now using $\gamma(U_i(t)) = h(t - S_{i,V_i(t^-)})$ where $V_i(t)$ is the counting process of vaccination times $\{S_{ij}\}$ and $t - S_{i,V_i(t^-)}$ is the time since the most recent vaccination. Estimation of the function $h(t - S_{i,V_i(t^-)})$ in this model assesses how the recency of a vaccine dose impacts the intensity of malaria infection.

Analysis 4. Analysis 4. applies the model to study whether and how time-varying VE against all-strain infection depends on the time since the last occurrence of infection. Similar to Analysis 2., all randomization arms are included in the analysis, and $Z_i$ is the indicator that participant i was randomly assigned to an RTS,S vaccination arm.

#### 8.6 Analytic Approach for Exploratory Objective 3

As for exploratory objective 2, analyses for exploratory objective 3 are done in the TVC cohort only and studying new molecularly detected infection events during the M0-20 and M0-32 follow-up periods.

The set of haplotypes k that are studied for exploratory objective 3 is the same set of haplotypes that are screened-in for the sieve analysis exploratory objective 1. In addition, sufficiently prevalent SERA2 haplotypes k may also be studied with exploratory objective 3.

Each of the four analyses described for exploratory objective 2 are repeated for each haplotype k. As always, the genotype categories are defined based on treatment-blinded analysis. For each defined genotype category, the four specified analyses are conducted.

### 9 POST-HOC EXPLORATORY ANALYSES

#### 9.1 Sensitivity Analysis of Vaccine Efficacy by Read Count Threshold for Parasite Positivity

We will estimate VE of each RTS,S vs. the control regimen in both the TVC and ATP cohorts using different parasite positivity sequence read count thresholds ranging between 50 and 1000. Point estimates and 95% CIs will be reported to evaluate sensitivity of the VE estimates to the threshold value.

#### 9.2 First New Asymptomatic Infection

It is of interest to assess VE of each RTS,S vaccine regimen to prevent the first new asymptomatic molecularly confirmed infection prior to any clinical malaria. To this end, we define the event of interest as the first new molecularly confirmed infection that does not meet the MAL-094 secondary case definition of clinical malaria (parasitemia > 0 parasites/μl). The rationale for this event definition is that all molecularly confirmed infections with any ***available*** evidence of clinical symptoms are disqualified, albeit some symptomatic infections may still be included such as mild symptomatic cases of clinical disease when either the child did not attend a clinic visit or the secondary case definition of clinical malaria was not met (e.g., if the axillary temperature was < 37.5°C).

The occurrence of the secondary case definition of clinical malaria is a competing event in this setting. Therefore, the following analyses might be performed to assess VE of each RTS,S regimen (i.e., comparisons with GP1) in both the TVC and ATP cohorts during primary follow-up:

(a) the competing risks Cox regression model, and
(b) the Aalen-Johansen estimation of cumulative incidence.

The output of the analysis in (a) will be a hazard-based VE estimate, with a 95% CI, for each RTS,S regimen and each cohort. The output of the analysis in (b) will be a cumulative incidence curve over time for each of GP1–GP5 and a cumulative VE curve over time for each of GP2–GP5, separately for each cohort.

#### 9.3 Vaccine Effects on Post-infection Outcomes (Analysis of Parasite Density)

Vaccine efficacy against clinical malaria measures a combination of vaccine effects on sterilizing immunity and other effects on post-infection processes such as parasitemia. Since the estimated overall VE against first clinical malaria is greater than the estimated overall VE against first molecular infection, it is of interest to investigate in a post-hoc analysis any vaccine effects on post-infection processes that would elucidate the higher VE against clinical disease. In particular, it is of interest to assess the marginalized mean difference (GP1 minus pooled GP2–5) in parasite density across all new molecularly confirmed infections in the TVC, controlling for baseline covariates that might predict both parasite density and the infection endpoint. Due to "differential cluster sizes" (different numbers of new infections per participant), multiple outputation will be used for valid inference. The analysis will be repeated for the first new molecularly confirmed infection in the TVC, without the need to use multiple outputation. For each of the two endpoints, we will show a histogram of parasite density, separately for GP1 and GP2–5, with zero as a separate category, and report a point estimate of the marginalized mean difference in parasite density with a 95% CI.

Given the finding that selective blockage of 3D7-matched infection acquisition does not explain sieve effects by 3D7 match vs. mismatch on clinical malaria, it is further desirable to evaluate in a post-hoc analysis whether a potential vaccine effect on parasitemia might explain these sieve effects. To this end, we will study whether parasitemia modifies VE against clinical malaria in the ATP cohort by fitting a mark-specific hazard-ratio model (Juraska and Gilbert, 2013) using parasitemia as a quantitative mark. Additionally, we will investigate whether 3D7 match modifies the vaccine effect on parasitemia in the ATP cohort by fitting a linear regression model for the mean parasite density with predictors treatment, indicator of 3D7 match, and an interaction of treatment and 3D7 match.

#### 9.4 Modification of RTS,S Vaccine Efficacy by Baseline Parasite Positivity and Month 2 Force of Infection

Herein, we define baseline parasite positivity as either molecular (≥ 325 read threshold) or microscopic detection of parasite positivity in the sample collected on the day of the first vaccination. We define month 2 force of infection (M2-FOI) as an individual's number of new molecular infections detected *after* the first vaccination visit and no later than at the month 2 scheduled visit (i.e., M2-FOI measures the number of intercurrent new infections post-first-vaccination by the month 2 visit).

Post-hoc analyses presented in this section aim to address four separate exploratory scientific questions:

1. Did baseline parasite positivity modify VE against the first new molecular infection?
2. Did baseline parasite positivity modify VE against the first new clinical malaria episode?
3. Did M2-FOI modify VE against the subsequent first new molecular infection?
4. Did M2-FOI modify VE against the subsequent first new clinical malaria episode?

We will investigate whether and how each of baseline parasite positivity and M2-FOI variables modified the effect of RTS,S vaccination on (i) the time to the first new molecular infection, and (ii) the time to the first new clinical malaria episode meeting the MAL094 protocol's primary case definition.

Questions 1. and 2. will be addressed in both the TVC and ATP cohorts, while questions 3. and 4. will be addressed in the ATP cohort only, given that M2-FOI is measured at the M2 visit (after randomization).

##### 9.4.1 Descriptive Analysis of Potential VE Modifiers of Interest

All descriptive analysis outputs (summary measures, figures, tables) will be generated for the individual as well as combined study sites. To characterize baseline positivity, the following descriptive outputs will be produced in both the TVC and ATP cohorts:

- prevalence of baseline parasite positivity pooled over GP1–4 and by treatment;
- a scatter/box plot of the date of the first vaccination stratified by baseline parasite positivity status pooled over GP1–4;
- a scatter/box plot of age at enrollment stratified by baseline parasite positivity status pooled over GP1–4.

To characterize M2-FOI, the following descriptive outputs will be produced in the ATP cohort only:

- frequency distribution of M2-FOI by GP2–4 vs. GP1 and by individual treatment arms;
- a bar plot of the percentage of GP1 vs. GP2–4 ATP participants (and of ATP participants by individual treatment arms) for each value of M2-FOI (to assess the association of treatment and M2-FOI);
- a bar plot of the percentage of baseline positive vs. baseline negative ATP participants for each value of M2-FOI, with a participant count on top of each bar (to assess the association of baseline positivity and M2-FOI) pooled over GP1–4;
- a scatter/box plot of the date of the first vaccination stratified by M2-FOI pooled over GP1–4;

- a scatter/box plot of age at enrollment stratified by M2-FOI pooled over GP1–4.

We will summarize correlation in the ATP cohort between baseline positivity and ordered categorical M2-FOI using Spearman's rho. Correlation in the ATP cohort between baseline positivity and the indicator that M2-FOI exceeds zero, I(M2-FOI > 0), will be assessed using Kendall's tau.

##### 9.4.2 Inferential Analysis of Potential VE Modifiers of Interest

Cox proportional hazards models, adjusted for baseline covariates, will be used for analysis. Assessments of vaccine effects within the baseline positive subgroup and within the baseline negative subgroup in the TVC or ATP cohort are valid based on randomization. If M2-FOI is uncorrelated or negligibly correlated with treatment assignment in the ATP cohort, assessments of vaccine effects separately within categories of M2-FOI = 0 and M2-FOI > 0 in the ATP cohort are approximately valid based on randomization, and there is minimal risk of post-randomization selection bias induced by subsetting on M2-FOI = 0 vs. M2-FOI > 0. Given that few participants with M2-FOI > 1 had first new malaria infection endpoints, statistical inferences studying M2-FOI analyze M2-FOI as a dichotomous variable I(M2-FOI > 0), interpreted as whether an intercurrent new malaria infection occurred after the first vaccination and by the third vaccination.

###### 9.4.2.1 Cox Proportional Hazards Analysis for Assessing Baseline Positivity as a VE Modifier

To address scientific questions 1. and 2. stated above, Cox models studying VE modification by baseline positivity in the TVC cohort will be adjusted for treatment group, baseline positivity, treatment × baseline positivity interaction, and age at enrollment (model M1-TVC). Cox models studying VE modification by baseline positivity in the ATP cohort will include the following independent variables:

- Model M1-ATP: Adjust for treatment group, baseline positivity, treatment × baseline positivity interaction, indicator of the onset of antimalarial drug treatment between the first vaccination and the month 2 scheduled visit (M2-mal-tx), age at enrollment, sex, and baseline levels of BMI and hemoglobin.

The baseline positivity status was not randomized such that the assessment of effect modification by baseline positivity is susceptible to potential confounding bias by any other covariate associated with baseline positivity and, at the same time, a VE modifier itself. Since M2-FOI may plausibly serve in this role, for the comparison of pooled RTS,S regimens GP2–4 vs. the control regimen GP1 for both individual and combined sites, we will also fit the following nested Cox models to ATP cohort data while controlling for potential confounding by M2-FOI (in addition to controlling for the main effects of M2-mal-tx, age at enrollment, sex, and baseline levels of BMI and hemoglobin):

- Model M2-ATP: Adjust for treatment group, baseline positivity, ordinal M2-FOI, M2-mal-tx, treatment × baseline positivity interaction, age at enrollment, sex, and baseline levels of BMI and hemoglobin
- Model M3-ATP: Adjust for treatment group, baseline positivity, ordinal M2-FOI, M2-mal-tx, treatment × baseline positivity interaction, treatment × ordinal M2-FOI interaction, age at enrollment, sex, and baseline levels of BMI and hemoglobin
- Model M4-ATP: Adjust for treatment group, baseline positivity, ordinal M2-FOI, M2-mal-tx, treatment × baseline positivity interaction, treatment × ordinal M2-FOI interaction, baseline positivity × ordinal M2-FOI interaction, age at enrollment, sex, and baseline levels of BMI and hemoglobin

We will report AIC values of models M1-ATP, M2-ATP, M3-ATP, and M4-ATP for the treatment comparison of GP2–4 vs. GP1 for the individual and combined sites as a way of characterizing the relative quality of the nested models.

All models fit to data pooling over both study sites will have a separate baseline hazard function for each site. All models will be fit in parallel for the first new molecular infection endpoint and the first new clinical malaria endpoint.

Cox models will be fit to assess hazard ratio-based VE (RTS,S/control) separately within the baseline positive and baseline negative subgroup, with 95% Wald CIs, and to generate treatment × baseline positivity two-sided interaction Wald test p-values for the following comparisons:

- **[TVC analyses]** RTS,S regimens GP2–4 pooled (i.e., excluding GP5) vs. the control regimen (GP1) for both combined and individual study sites in the TVC cohort.
- **[ATP analyses]** RTS,S regimens GP2–4 pooled (i.e., excluding GP5) vs. the control regimen (GP1) for both combined and individual study sites in the ATP cohort. GP5 is excluded from the RTS,S pooled group in both the TVC and ATP analyses because the post-dose-3 ATP follow-up period in GP5 is delayed until M7.5–19, whereas, for all other groups including the control group, the follow-up period is M2.5–14. The exclusion of GP5 in the analyses of pooled RTS,S vs. control is of little concern because we hypothesize that neither baseline positivity nor M2-FOI are likely to impact VE during a follow-up period starting 7.5 months after baseline. In addition, GP5 vs. control VE modification analyses in the TVC cohort during the M0–20 period and in the ATP cohort during the M7.5–19 period are evaluated separately as specified in the next bullet.
- Each individual RTS,S regimen (except ATP GP2–3 pooled) vs. the control regimen pooling over the study sites as summarized in the following table:

| Analysis Cohort | Study Group and Follow-up Comparison |
|---|---|
| TVC | GP2 M0-20 vs. GP1 M0-20 |
| | GP3 M0-20 vs. GP1 M0-20 |
| | GP4 M0-20 vs. GP1 M0-20 |
| | GP5 M0-20 vs. GP1 M0-20 |

| | |
|---|---|
| ATP | GP2 + GP3 pooled M2.5-14 vs. GP1 M2.5-14 |
| | GP4 M2.5-14 vs. GP1 M2.5-14 |
| | GP5 M7.5-19 vs. GP1 M7.5-19 |

The ATP cohort analyses will consider the follow-up period from 14 days after the 3rd dose through the scheduled visit at 12 months after the 3rd visit for each of GP1–5.

In addition to the models described above, we will fit model M1-ATP for the GP2–4 vs. GP1 treatment comparison for the follow-up period restricted to 14 days to 4.5 months after the 3rd dose for assessments of VE modification during a period of potentially limited waning of overall VE against the first new infection (or the first clinical malaria).

###### 9.4.2.1.1 Sensitivity 'Matching' Cox Proportional Hazards Analysis Assessing Baseline Parasite Positivity as a Modifier of VE

For further evaluating baseline positivity as a potential VE modifier of interest, we will conduct a sensitivity 'matching' ATP cohort Cox analysis which involves stratified sampling, wherein baseline negative participants will be randomly sampled from the same randomization group and study site by matching baseline positive participants on the date of the third dose administration. All models assessing VE of pooled RTS,S (GP2–4) vs. control regimens in the ATP cohort as described in Section 9.4.2.1. will be fit to data from matched random samples, with results averaged across 1000 resampled data sets. The purpose of the matching analysis is to eliminate the possibility that the modifying impact of baseline parasite positivity on VE is confounded by well-timed vaccination at the start of a high-transmission season. This confounding could arise if the baseline positivity status was correlated with calendar time of the third dose administration just prior to the start of a high-transmission season and, at the same time, the effect of RTS,S vaccination on the study endpoint was differential in low- vs. high-transmission seasons.

The matching analysis will use the following stratified sampling design: For each baseline positive participant, $m$ baseline negative participants will be randomly sampled from the same randomization group and study site by matching on the date of the third dose administration, with $m = 1, 2, 3$. The rationale for $m > 1$ is increased precision in estimation of Cox model coefficients by adding hundreds of additional baseline negative matches (the second and third matches). Matching will be performed by prioritizing matches on the exact date of the third dose administration, and only if such matches are unavailable, then the search will be expanded sequentially to ∓1 day, ∓2 days,…, ∓7 days of the date of the third dose administration. No matching will be considered beyond the interval of ∓7 days of the date of the third dose administration. Sampling of $m$ baseline negative matches will be performed without replacement for each baseline positive participant. Across all baseline positive participants, sampling of baseline negative matches will be performed without replacement of sampled matches to other baseline positive participants. A random permutation of baseline positive participants will be used in each iteration to avoid 'more accurate matching' of baseline positive participants on the top of the sampling list. To describe the matching accuracy of the sampling algorithm, a stacked bar plot will be generated for each $m$ showing the proportions of baseline positive participants with matches within intervals of $\mp 0$, $\mp 1$,..., $\mp 7$ days of the date of the baseline positive participants' 3rd dose administration. The proportions will be reported as sample medians across 1000 replications. For $m > 1$, each baseline positive participant will be assigned to a color-coded category according to the match (among the $m$ matches) sampled from the widest time interval.

Sample medians of estimated hazard ratio-based VEs and confidence interval limits for the baseline positive and baseline negative subgroups as well as treatment × baseline positivity interaction test p-values will be reported. Besides the sample medians, the whole distributions across the 1000 replications will be plotted.

###### 9.4.2.2 Cox Proportional Hazards Analysis for Assessing M2-FOI as a VE Modifier

To address scientific questions 3. and 4. stated above, we will study the indicator I(M2-FOI > 0) rather than ordinal M2-FOI as a VE modifier because it eliminates the need to model the dose-response association parametrically and pools presumably smaller categories in the right tail of M2-FOI support.

The first Cox model studying VE modification by I(M2-FOI > 0) in the ATP cohort will include the following independent variables:

- Model M5-ATP: Adjust for treatment group, I(M2-FOI > 0), M2-mal-tx, treatment × I(M2-FOI > 0) interaction, age at enrollment, sex, and baseline levels of BMI and hemoglobin

I(M2-FOI > 0) was not randomized such that, analogous to Section 9.4.2.1, the assessment of effect modification by I(M2-FOI > 0) is susceptible to potential confounding bias by any other covariate associated with I(M2-FOI > 0) and, at the same time, a VE modifier itself. Since baseline positivity may plausibly serve in this role, for the comparison of pooled RTS,S regimens GP2–4 vs. the control regimen GP1 for both individual and combined sites, we will also fit the following nested Cox models to ATP cohort data while controlling for potential confounding by baseline positivity:

- Model M6-ATP: Adjust for treatment group, I(M2-FOI > 0), baseline positivity, M2-mal-tx, treatment × I(M2-FOI > 0) interaction, age at enrollment, sex, and baseline levels of BMI and hemoglobin
- Model M7-ATP: Adjust for treatment group, I(M2-FOI > 0), baseline positivity, M2-mal-tx, treatment × I(M2-FOI > 0) interaction, treatment × baseline positivity interaction, age at enrollment, sex, and baseline levels of BMI and hemoglobin
- Model M8-ATP: Adjust for treatment group, I(M2-FOI > 0), baseline positivity, M2-mal-tx, treatment × I(M2-FOI > 0) interaction, treatment × baseline positivity interaction, baseline positivity × I(M2-FOI > 0) interaction, age at enrollment, sex, and baseline levels of BMI and hemoglobin

We will report AIC values of models M5-ATP, M6-ATP, M7-ATP, and M8-ATP for the treatment comparison of GP2–4 vs. GP1 for the individual and combined sites as a way of characterizing the relative quality of the nested models.

All models fit to data pooling over both sites will have a separate baseline hazard function for each site. All models will be fit in parallel for the first new molecular infection endpoint and the first new clinical malaria endpoint.

Cox models will be fit to assess hazard ratio-based VE (RTS,S/control) separately within subgroups defined by M2-FOI = 0 and M2-FOI > 0, with 95% Wald CIs, and to generate treatment × I(M2-FOI > 0) two-sided interaction Wald test p-values for the following comparisons in the ATP cohort:

- RTS,S regimens GP2–4 pooled (i.e., excluding GP5) vs. the control regimen (GP1) for both combined and individual study sites in the ATP cohort. GP5 is excluded from the RTS,S pooled group in the ATP analysis because the post-dose-3 ATP follow-up period in GP5 is delayed until M7.5–19, whereas, for all other groups including the control group, the follow-up period is M2.5–14. The exclusion of GP5 in the ATP analysis of pooled RTS,S vs. control is of little concern because we hypothesize that neither baseline positivity nor M2-FOI are likely to impact VE during a follow-up period starting 7.5 months after baseline. In addition, GP5 vs. control VE modification during the M7.5–19 period is evaluated separately as specified in the next bullet.
- Each individual RTS,S regimen (except ATP GP2–3 pooled) vs. the control regimen pooling over the study sites as summarized in the following table:

| Analysis Cohort | Study Group and Follow-up Comparison |
|---|---|
| ATP | GP2 + GP3 pooled M2.5-14 vs. GP1 M2.5-14 |
| | GP4 M2.5-14 vs. GP1 M2.5-14 |
| | GP5 M7.5-19 vs. GP1 M7.5-19 |

The ATP cohort analyses will consider the follow-up period from 14 days after the 3$^{rd}$ dose through the scheduled visit at 12 months after the 3$^{rd}$ visit for each of GP1–5.

In addition to the models described above, we will fit model M5-ATP for the GP2–4 vs. GP1 treatment comparison for the follow-up period restricted to 14 days to 4.5 months after the 3$^{rd}$ dose for assessments of VE modification during a period of potentially limited waning of overall VE against the first new infection.

###### 9.4.2.3 Sensitivity Analysis to Unmeasured Confounding

For selected Cox model analyses specified above, for studying each of baseline parasite positivity and I(M2-FOI > 0) as modifiers of vaccine efficacy, the method of Mathur et al. (2022) will be used to quantify the amount of unmeasured confounding that would need to exist, in order to explain away that modification of vaccine efficacy truly exists, after accounting for all of the measured potential confounders included in the model. This method will be applied both for the point estimate of the ratio of hazard ratios (one hazard ratio for each positive or negative subgroup), as well as for the confidence limit of the ratio of hazard ratios.

###### 9.4.2.4 Cumulative Incidence and Instantaneous Hazard Estimation

To assess potential differences in timing of the occurrence of endpoint events, Kaplan-Meier cumulative incidence estimates over time and nonparametric kernel-smoothed instantaneous hazard rate estimates over time will be plotted for the following subgroups defined by the potential VE modifiers of interest in the ATP cohort:

- baseline parasite positive subgroups
    - baseline parasite positive control regimen recipients
    - baseline parasite positive pooled RTS,S regimen recipients (GP2–4 excluding GP5)
    - baseline parasite negative control regimen recipients
    - baseline pasrasite negative pooled RTS,S regimen recipients (GP2–4 excluding GP5)
- M2-FOI subgroups
    - control regimen recipients with M2-FOI = 0
    - pooled RTS,S regimen recipients (GP2–4 excluding GP5) with M2-FOI = 0
    - control regimen recipients with M2-FOI > 0
    - pooled RTS,S regimen recipients (GP2–4 excluding GP5) with M2-FOI > 0.

### 11 LIST OF ABBREVIATIONS

1. AS01E GlaxoSmithKlines proprietary Adjuvant System containing MPL, QS-21 and liposome (25 g MPL and 25 g QS-21)
2. ATP according to protocol
3. CDC Center for Disease Control
4. CI confidence interval
5. COI complexity of infection
6. CRF case report forms
7. CRO clinical research organization
8. CS circumsporozoite protein of Plasmodium falciparum
9. csp DNA seq for circumsporozoite gene
10. C-terminus C-terminal domain from CSP protein
11. DNA deoxyribonucleic acid
12. EMA European Medicines Agency
13. FOI Force of Infection
14. Fx fractionated / fractional
15. GP Genomics Platform
16. GSK GlaxoSmithKline
17. LIMS laboratory information management system
18. LL lower limit

19. mol(FOI) molecular Force of Infection

20. msp merozoite surface protein gene from Pf

21. MVI Malaria Vaccine Initiative

22. NCBI National Center for Biotechnology Information

23. PE proportional hazards

24. PCR polymerase chain reaction

25. P. falciparum Plasmodium falciparum

26. POC proof of concept

27. PI principal investigator

28. pyr per year

29. RCDFs reverse cumulative distribution functions

30. RTS,S Particulate antigen, containing both RTS and S (hepatitis B surface antigen) proteins

31. SERA2 serine repeat antigen 2 from Pf

32. SNP single nucleotide polymorphisms

33. TBS thick blood smear

34. TVC total vaccinated cohorts

35. VE vaccine efficacy

TRADEMARKS

WHATMAN is a trademark of GLOBAL LIFE SCIENCES SOLUTIONS OPERATIONS UK LTD.

QIAamp and HotStarTaq are trademarks of the QIAGEN Group.

PicoGreen is a trademark of Invitrogen, Inc.

Illumina is a trademark of Illumina, Inc.

LabChip and Caliper are trademarks of Caliper Life Sciences, Inc.

AGENCOURT AMPURE XP is a trademark of Agencourt Bioscience.

BioAnalyzer is a trademark of Agilent Technologies, Inc.

DNA ZAP is a trademark from Ambion company.
